# Working memory in chronic pain: evidence for task-specific rather than global differences

**DOI:** 10.64898/2026.03.20.26348805

**Authors:** Monika Halicka, Meike Scheller, Christopher Brown

## Abstract

**Background:** Chronic pain is often accompanied by cognitive complaints, but evidence for global working-memory problems is mixed. We tested whether working-memory performance differences in chronic pain are global or task-specific and whether model-based analysis could help distinguish differences in processing efficiency, response caution or sensory/motor speed.

**Methods:** In a preregistered online case-control study, 99 adults with mixed chronic pain conditions and 87 pain-free controls completed visuo-verbal, visuo-spatial and auditory–temporal n-back tasks at low (0/1-back) and high (2-back) load. Accuracy and reaction times were analysed with mixed effects models. Drift diffusion modelling decomposed performance into processing efficiency (drift rate), response caution (threshold separation) and non-decision (sensory/motor) time.

**Results:** Higher load reduced accuracy and slowed responses in both groups. There was no evidence of a global working-memory deficit in the chronic pain group. The clearest group difference was a larger load-related drop in accuracy in the auditory-temporal task (odds ratio 0.64, 95% CI 0.56 to 0.73), persisting after adjustment for mood, sleep and fatigue. Load-related slowing in visuo-verbal (6.7% slower, 5.1% to 8.2%) and auditory–temporal tasks (3.6% slower, 1.7% to 5.4%) were attenuated after adjustment. Diffusion modelling showed no evidence for sensory/motor slowing, but rather greater response caution in the auditory–temporal task and small efficiency (drift rate) reductions in low-load visual conditions.

**Conclusions:** The results do not support a global working-memory capacity loss account in this mixed chronic pain sample. Rather, they suggest task-specific performance differences, most evident in auditory–temporal processing, with response caution as a plausible contributor.

**Significance Statement:** This preregistered multimodal online study found no evidence of a global working-memory deficit in a mixed chronic pain sample. Instead, the clearest difference emerged in the auditory-temporal n-back task under higher load, with diffusion modelling suggesting a contribution of greater response caution rather than reduced processing efficiency or sensory-motor slowing.

## 2 Introduction

Chronic pain is common, disabling and costly (Rice et al., 2016). Alongside sensory symptoms, many patients report attention and memory difficulties affecting daily functioning (Timm et al., 2025). While pain may influence cognitive function, prospective studies suggest cognitive function may also influence pain (Ng & Hartanto, 2022). Neuroimaging work also implicates fronto-parietal and subcortical networks relevant to cognitive control in chronic pain (Baliki et al., 2011, 2012; Henry et al., 2011; Pinheiro et al., 2016; Veinot et al., 2025). Clarifying the nature of cognitive differences in chronic pain may therefore help explain functional difficulties and identify targets for assessment and intervention.

Meta-analyses suggest small-to-moderate differences in executive function (Berryman et al., 2014) and working memory (WM) (Berryman et al., 2013) in chronic pain vs controls, although the pattern and magnitude of these differences are mixed. While many studies imply global cognitive deficits, task-specific differences have also been reported including spatial attention, mental rotation and temporal discrimination (Breckenridge et al., 2019; Bultitude et al., 2017; Cohen et al., 2013; Gunendi et al., 2019; Halicka et al., 2020; Kuttikat et al., 2016). Some evidence also points to selective vulnerability of WM updating rather than a broad executive deficit (Rader et al., 2025).

Most prior WM studies in chronic pain relied on simple metrics such as mean accuracy and reaction time, which conflate multiple latent processes. Model-based approaches such as hierarchical drift diffusion modelling (HDDM) can separate observed performance into components such as evidence accumulation, response caution and non-decision time. This is useful here because slower performance in chronic pain could reflect reduced working-memory efficiency, a more cautious response strategy, or slower stimulus encoding and response execution. Such approaches have also linked sensory evidence accumulation to parietal lobe function (Huk & Meister, 2012), which may be particularly informative when testing whether any group differences are more evident in tasks with greater spatial demands. Because many previous studies have relied on broad behavioural summaries, it remains unclear whether reported WM differences in chronic pain reflect global problems or more task-specific effects. In addition, mood disturbance, sleep disturbance and fatigue commonly accompany chronic pain and may themselves influence cognitive performance, but are not always measured or accounted for.

Our main aim was to test whether WM performance differences between people with and without chronic pain were global or task-specific across modalities and cognitive load, and whether any differences were sensitive to mood, sleep disturbance and fatigue. Given prior evidence for WM differences in chronic pain, we expected poorer performance in chronic pain under higher load and in task modalities placing greater spatial, and possibly temporal-sequential, demands. Using diffusion modelling, we also asked whether observed group differences were more consistent with reduced evidence accumulation, altered response caution, or changes in non-decision time. To address this, we compared visuo-verbal, visuo-spatial and auditory-temporal sequence-matching n-back tasks under lower and higher load. Online testing provided efficient sampling across multiple tasks.

## 3 Methods

This was a preregistered online unmatched case-control study in a mixed community sample of adults with chronic pain and pain-free controls. Full pre-registered hypotheses are in Supplementary Materials.

### 3.1 Participants

#### 3.1.1 Eligibility

All participants had to be 18 - 65 years old, live in the UK, have (corrected-to-) normal vision and hearing, and have no history of neurological disorders (such as a stroke, traumatic brain injury, neurodegenerative disease, epilepsy). Participants with chronic pain should have been experiencing pain that persisted (was present on most days) or recurred (as repeating episodes) for the past three months or more. Pain-free control participants should not have been diagnosed with any chronic pain condition, or experience any pain during the study. Participants required access to an internet-connected desktop or laptop computer with a physical keyboard, and headphones, to complete the online tasks. Participants also required sufficient English comprehension to understand task instructions and questionnaire items.

#### 3.1.2 Recruitment

Participants were recruited in March 2021 to October 2021 through word of mouth, online advertisements on University and community websites, chronic pain support groups and charities, and through online recruitment platform Prolific. Participants were reimbursed for their time. The Institutional Research Ethics Committee approved this research and all participants provided informed consent. Patients and the public were not involved in the design, conduct, reporting, or dissemination plans of this research. Recruitment was monitored by age band and gender identity to improve comparability between groups, with an approximately 1:1 case-control ratio targeted during accrual.

#### 3.1.3 Sample size

The sample size estimation was based primarily on the main case-control comparison and details are available in the preregistration record (https://doi.org/10.17605/OSF.IO/TH7DV). Drawing on prior literature showing variable but generally modest group differences in attention and working-memory performance in chronic pain (Berryman et al., 2013, 2014; Coppieters et al., 2015; Oosterman et al., 2012; Sjøgren et al., 2005), we planned for 80% power to detect a standardised between-group difference of Cohen’s d = 0.4 between participants with and without chronic pain. This suggested a sample of 78 participants per group (156 in total) under the preregistered assumptions. Allowing for approximately 20% loss of data, we aimed to recruit up to 97 participants per group (194 in total). These calculations should be interpreted as supporting the primary comparison rather than every secondary or exploratory analysis. Because the main behavioural analyses used trial-level mixed-effects models for accuracy and reaction time, they may be more statistically efficient than analyses based only on participant-level summary scores; however, usable sample size still varied across tasks after quality control, so precision was lower for some task-specific analyses.

Secondary within-group regression analyses were considered separately. For linear multiple regression with six predictors on neuropsychological tasks outcomes (e.g. to test pain chronicity as a predictor), a sample of 55 participants with chronic pain would be required to detect a medium effect (Cohen’s *f^2^* = 0.15) (Jongsma et al., 2011) with 80% power and alpha of 0.05.

### 3.2 Procedures

Participants accessed the study via a link to the Gorilla platform (www.gorilla.sc) (Anwyl-Irvine et al., 2020) and completed each session online. Online administration increased reach but introduced uncontrolled hardware and environmental variation; this was partly mitigated by restricting participation to desktop/laptop devices and by requiring sound-volume adjustment and a headphones check before the auditory conditions. The study flow is illustrated in Figure 1. Following familiarisation with a participant information sheet and signing an electronic consent form, participants completed a brief real effort task and a bot check, allowing to eliminate automatic or fraudulent responses. Next, participants completed self-report questionnaires regarding their demographics, mood, sleep, fatigue, and if they indicated experiencing chronic pain, also a chronic pain symptoms and treatments survey. Then they were randomised (without replacement, with equal ratio) to one of two response mappings, where either ‘F’ or ‘J’ key corresponded to a ‘target’ or ‘non-target’ response. Next, participants completed three types of cognitive tasks (visuo-verbal [VV], visuo-spatial [VS], and auditory-temporal [AU]) in a randomised order. Each included a low (0/1-back) and a high (2-back) cognitive load condition, order of which was also randomised within each task type. Each task condition was preceded by 10 trials of training and followed by a post-experiment survey regarding current levels of fatigue, pain, engagement, and perceived task difficulty, and whether participants were interrupted during the task and for how long. The AU tasks were additionally preceded by sound volume adjustment and headphones check (Milne et al., 2021; Woods et al., 2015) to reduce background noise and ensure adequate perception of the auditory stimuli. After one week from completing the first session, participants received a link to the second session (to be completed within a week), which followed the same procedure, except that the self-report questionnaires only measured mood, fatigue, and pain severity (for participants with chronic pain). Each session lasted up to 50 minutes.

**Figure 1.**
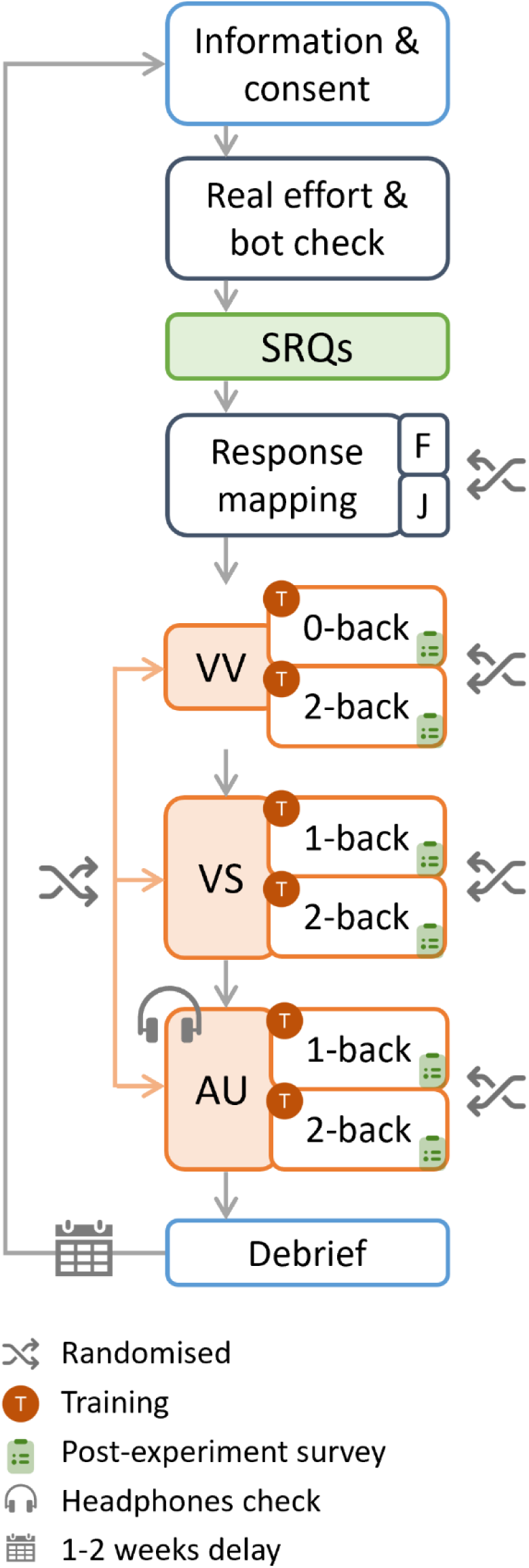
Study flow. SRQs, self-report questionnaires; F, J, response keys; T, training; VV, visuo-verbal task; VS, visuo-spatial task; AU, auditory-temporal task.

### 3.3 Measures

#### 3.3.1 Self-report questionnaires

In the *demographic survey*, participants reported their age and gender identity. Participants also indicated whether they were experiencing chronic pain, or any current pain. We did not collect race/ethnicity data, and education was not assessed.

The *mood, sleep, and fatigue survey* included a short form of Profile of Mood States (McNair et al., 1971; Shacham, 1983), a recommended measure of emotional functioning in chronic pain research (Dworkin et al., 2005). Participants rated how they felt at the moment in relation to 37 adjectives describing mood states using a 5-point scale ranging from ‘not at all’ to ‘extremely’. The responses were scored to obtain a total mood disturbance score, as well as tension-anxiety, depression, anger-hostility, vigour, fatigue, and confusion subscale scores. Considering the effects of fatigue on cognitive performance (Suhr, 2003), in particular sustained attention, we used a single-item Rhoten Fatigue Scale (Rhoten, 1982), where participants rated their current level of fatigue on a numerical rating scale, ranging from 0 (‘not tired, full of energy’) to 10 (‘total exhaustion’). Considering emerging evidence of bidirectional relationships between sleep disturbance and severity of chronic pain (Alsaadi et al., 2014; Husak & Bair, 2020; Mathias et al., 2018; Miettinen et al., 2021) and the effects of sleep deprivation on cognitive performance (Wardle-Pinkston et al., 2019), we included a 4-item standardised Sleep Disturbance Scale (Jenkins et al., 1988), where participants rated how often they experienced various sleep disturbances over the past month on a 0 (‘not at all’) to 5 (’22-31 days’) scale. They additionally reported how many hours they slept during the night before the study.

Participants with chronic pain additionally completed a c*hronic pain symptoms survey.* They indicated for how long they have been experiencing pain, in which areas of the body they usually experienced pain and where the pain was most severe, whether they received any medical diagnosis for their pain condition, if yes, what diagnosis and how long ago. Participants further rated their worst and least pain in the last 24 hours, average pain, and current pain intensity on numerical rating scales ranging from 0 (‘no pain’) to 10 (‘pain as bad as you can imagine’). An average of these four items formed a Brief Pain Inventory (Cleeland, 1996) severity score. Participants also indicated what type of treatments and/or medications they used for their pain regularly and as needed, and rated how much relief these have provided in the past 24 hours on a scale from 0% (‘no relief’) to 100% (‘complete relief’). Medication and treatment use were therefore characterised descriptively in the chronic pain group, but detailed medication burden was not modelled in the primary case-control analyses.

In the *post-experiment surveys*, participants rated their current levels of fatigue and pain (as described above), as well as levels of engagement while completing the tasks and their perceived difficulty on numerical rating scales ranging from 0 (‘not engaged’ / ‘not difficult at all’) to 10 (‘fully engaged’ / ‘extremely difficult’).

#### 3.3.2 Cognitive tasks

There were six task conditions across three task modalities: Visuo-verbal (VV) 0-back and 2-back, visuo-spatial (VS) 1-back and 2-back, and auditory-temporal (AU) 1-back and 2-back. The three task types were intended to sample different stimulus modalities while preserving a comparable n-back structure across conditions. Each condition required a binary target/non-target decision and each task modality included a lower-load (0/1-back) and higher-load (2-back) version. Participants were instructed to respond as accurately and as quickly as possible. Figure 2 illustrates example trial sequences. Additional task-specification details, including trial composition, stimulus timing, fixed stimulus orders, and lure structure, are provided in the Supplementary Materials.

**Figure 2.**
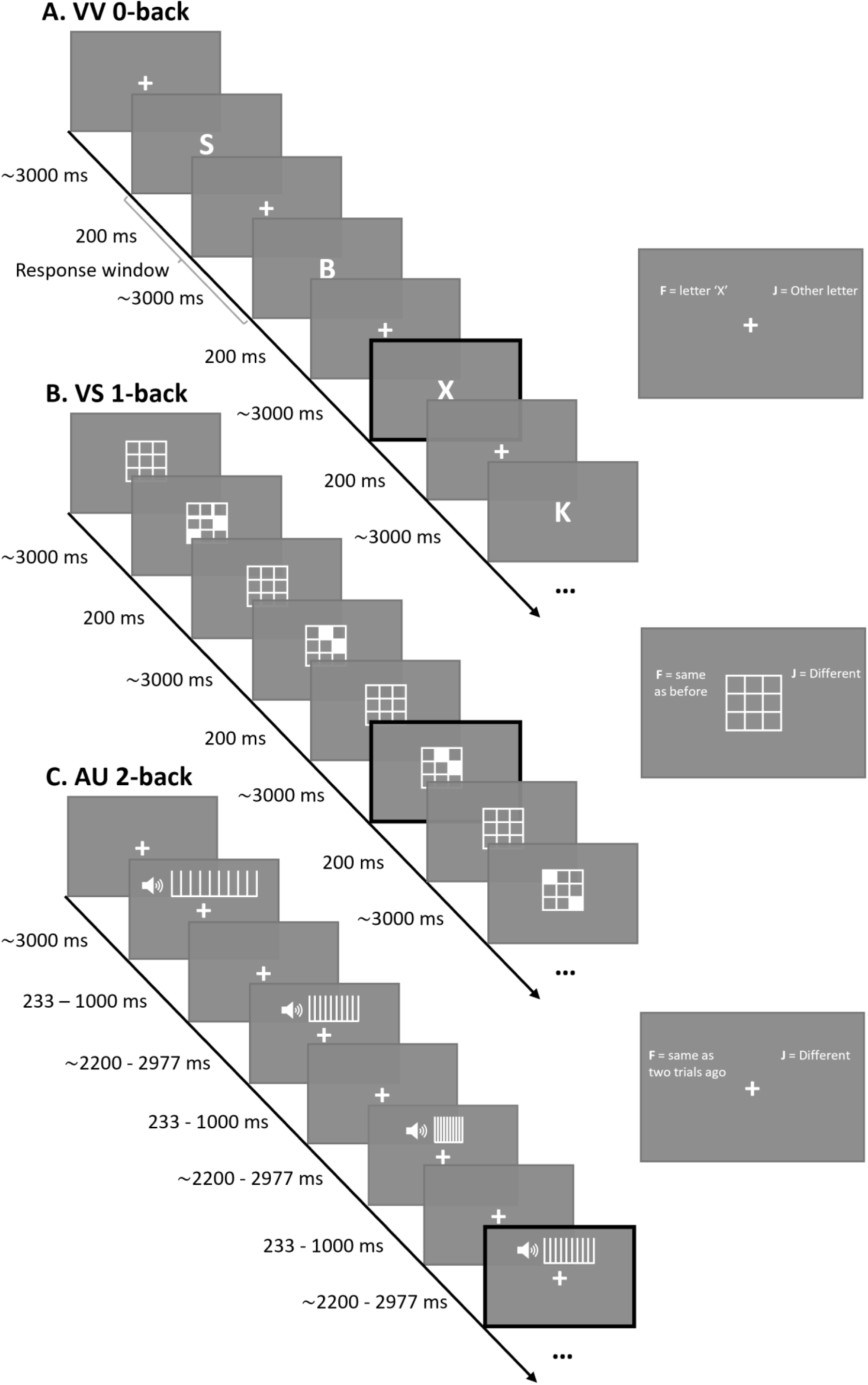
Example trial sequences for the visuo-verbal (VV), visuo-spatial (VS), and auditory-temporal (AU) tasks. Targets are indicated by black frames. In VV 0-back, the target was the letter X. In VS 1-back, the target was a grid pattern matching the previous trial. In AU 2-back, the target was a sound matching the frequency of the sound presented two trials earlier. Bars representing AU frequencies are for illustration only and were not displayed during the task. Instruction prompts remained on the screen throughout the duration of the task (here presented separately from the task timeline for clarity).

In the visuo-verbal task, stimuli were single consonant letters. In the 0-back condition, participants responded “target” whenever the prespecified target letter appeared. In the 2-back condition, the target was a letter matching the one presented two trials earlier. In the visuo-spatial task, stimuli were 3 × 3 grid patterns with two filled squares. In the 1-back condition, a target was a pattern identical to the one on the immediately preceding trial. In the 2-back condition, the target matched the pattern presented two trials earlier. In the auditory-temporal task, stimuli were tone trains varying in temporal frequency. In the 1-back and 2-back conditions, a target was a sound matching the frequency of the sound presented one or two trials earlier, respectively. Stimuli were piloted to ensure that adjacent frequencies were discriminable, and only a fixation cross was displayed during AU tasks. Full stimulus details for the three tasks are provided in the Supplementary Materials.

Each task condition was preceded by a 10-trial practice block with feedback. Participants could repeat practice once before proceeding, and could skip a task if, after retraining, they still did not understand the instructions. No trial-wise feedback was provided during the main tasks. After each task condition, participants were shown their total number of correct responses. Further practice-block and feedback-timing details are provided in the Supplementary Materials.

### 3.4 Statistical analysis

#### 3.4.1 Data processing

The data was processed and analysed using R software version 4.1.1 (R Core Team, 2021). Participants’ performance in each task condition was quantified as (i) overall accuracy (number of correct responses divided by the total number of trials); (ii) average response times (RTs) on correct trials; (iii) in VV 0-back task, additionally as variability of RTs on correct trials (SD of RTs divided by participant’s mean RTs). Additional analyses (Supplementary Materials) included derived metrics from Signal Detection Theory.

We excluded participant’s data if they failed the prespecified completeness and quality checks (responded on <80% trials regardless of accuracy, reported being interrupted or distracted for >30% of a task or experiencing technical difficulties, provided non-logical responses to free-text and multiple-choice questions, or failed the headphones check [< 5/6 correct responses] if coupled with other unusual responses). Twenty-nine participants were excluded. If the insufficient number of responded-to trials only concerned individual tasks, participant’s data was retained and only those tasks were excluded from analysis.

Furthermore, 0- and 1-back task data with accuracy rates <55%, i.e., the upper limit of the distribution of accuracy rates not significantly different from random guessing (Steffens et al., 2020), were excluded from analysis, as were the corresponding 2-back task data. No-better-than-chance accuracy on 0/1-back tasks would indicate that participants were guessing or were not able to discriminate the target and non-target stimuli, yielding both their 0/1-back and 2-back tasks data not interpretable.

The number of participants included in the analysis and those excluded due to insufficient numbers of responded-to trials and no-better-than-chance accuracy is presented in Supplementary Materials. Overall, 23 control participants and 41 participants with chronic pain had at least one task data removed (in either session), however, their baseline characteristics did not significantly differ from those participants whose complete data was included in analysis, except for some differences in pain medications and diagnoses (see Supplementary Materials). Of the 186 participants completing session 1, 154 (81%) completed session 2; detailed comparisons between completers and non-completers are reported in the Supplementary Materials.

RTs exceeding +/−2.5 SDs of individual participant’s mean RT in each task, as well as RTs <200ms, were excluded (2.73% of all responses across all participants, tasks, and sessions).

To allow calculating total mood disturbance scores, missing values in POMS (1% of all responses across all participants and sessions) were replaced by individual participant’s average score on the relevant subscale. Missing responses were found in the post-experiment surveys (pain and fatigue < 1%, engagement and perceived difficulty 16%), sleep disturbance (< 1%), baseline fatigue and hours slept the night before the study (2%), and BPI (1%). There were no missing values on any other questionnaire measures.

#### 3.4.2 Validity and reliability

We preregistered additional sensitivity and reliability checks, including expected worse performance at higher load, test–retest agreement, exploratory convergent/discriminant comparisons across low-load tasks, and within-session deterioration on low-load tasks as an index of fatigue sensitivity. Full methods and results for these checks are reported in the Supplementary Materials.

#### 3.4.3 Group effect analyses: primary

Our primary analyses compared participants with chronic pain with pain-free controls. We used mixed effects models (*lme4* package (Bates et al., 2022) with group (chronic pain, pain-free control), cognitive load (0/1-back, 2-back), and their interaction as fixed effects, participant as random effect, and task performance metrics as outcomes.

Generalised linear mixed models (*glmer*) with binomial family were fitted to trial-level accuracy data, and linear mixed models (*lmer*) to trial-level RT data, which were log-transformed to achieve normal distribution. The main effect of group tested for overall group differences across load conditions, whereas the interaction between group and cognitive load tested whether any group differences were more evident in higher working memory load conditions.

If the interaction was evidenced, we conducted follow-up analyses to examine whether load-related group differences varied across task types. For each task, we subtracted 0/1-back performance from 2-back performance, yielding participant-level load-difference scores. These outcomes were analysed using linear mixed models with group (chronic pain, healthy control), task type (visual, visuo-spatial, auditory-temporal), and group by task type interaction as fixed effects, and participant as random effect. Any significant interaction effects from this analysis were followed up by planned contrasts within each group (with Holm-Bonferroni correction for multiple comparisons) between VV vs VS, and VV vs AU task type.

For ease of interpretation, we additionally translated selected key model estimates into more familiar metrics in the text. Specifically, for models fit on log-transformed RTs, selected coefficients were exponentiated and expressed as percent change in RT (100×(exp(b)−1)); for logistic models, selected log-odds estimates were exponentiated and expressed as odds ratios (OR=exp(b)). All primary model outputs in tables and figures are presented on the original model scale (log-RT or log-odds).

#### 3.4.4 Group effect analyses: secondary

There were large between-group differences in mood disturbance (POMS), sleep disturbance, and baseline fatigue (see Results, Participant Characteristics section). As part of our preregistered secondary analysis, we included these factors as covariates in the mixed effect models. Although there were moderate-large correlations between mood disturbance, sleep disturbance, and baseline fatigue, none of the Pearson’s coefficients exceeded 0.7, and including all three factors did not produce multicollinearity (variance inflation factors < 5). Mood disturbance was scaled (mean-centred) to be incorporated into the mixed effects models.

Age and gender identity were not included as covariates in the primary case–control mixed models because groups were similar on these variables at baseline (Table 1) and our preregistered adjustment focused on symptoms that differed between groups and were expected a priori to influence cognitive performance (mood, sleep, fatigue). We also aimed to limit the number of covariates to preserve precision and avoid overfitting. Age was included only in preregistered within-group (chronic pain) models examining predictors of load-related performance change. Education, race/ethnicity, and more detailed medication burden were not available as covariates; medication use was instead characterised descriptively and opioid use was considered only in preregistered within-group models.

**Table 1.**
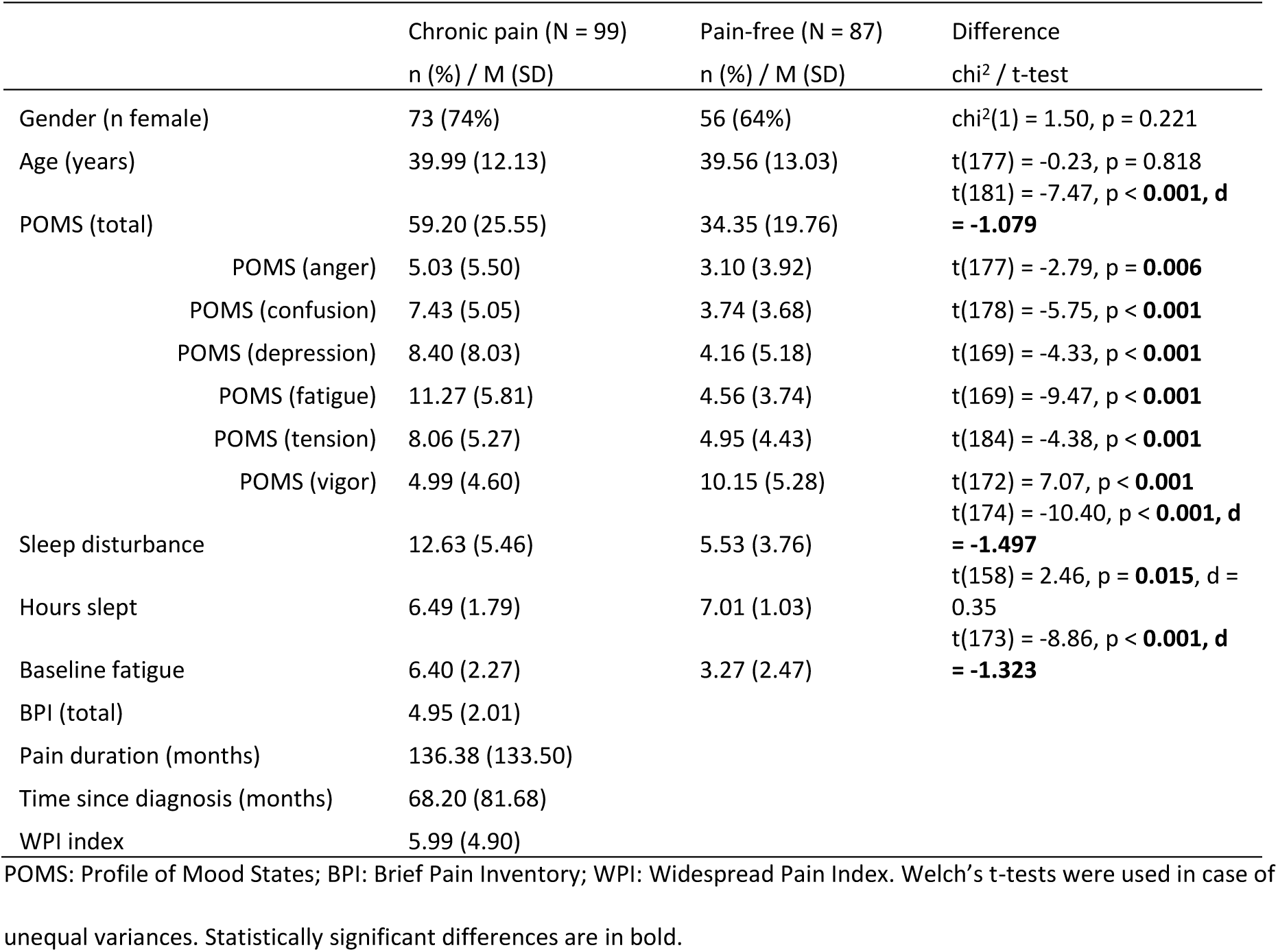
Participant characteristics at baseline and statistical comparisons between groups.

#### 3.4.5 Session 2 analyses

To assess whether any group effects found in session 1 replicate over time in the same participants, we analysed session 2 data as per planned analyses described above. Since sleep disturbance was measured on a scale of the past month (and the time interval between the sessions was 2 weeks), this measure was not repeated in session 2, therefore, session 1 sleep disturbance score was included as a covariate in session 2 models.

#### 3.4.6 Predictors of performance: Within-group regression analyses

We conducted preregistered regression analyses within the chronic pain group to examine whether load-related performance change was associated with pain duration after adjustment for pain severity, mood disturbance, baseline fatigue, sleep disturbance, regular opioid use, and age. Full methods and results are reported in the Supplementary Materials.

#### 3.4.7 Model-based exploratory analyses - HDDM

Group effects were further explored using hierarchical drift diffusion models (HDDMs), which use trial-level accuracy and reaction-time distributions to decompose performance in two-choice tasks into latent components of evidence accumulation, response caution, and non-decision time. In the present n-back tasks, participants judged whether the current stimulus matched the relevant previous stimulus, making performance well suited to a two-boundary decision model (Ratcliff & Rouder, 1998).

Drift rate (*v*) indexes the efficiency of evidence accumulation (lower values indicate poorer or noisier information processing), threshold separation (*a*) indexes response caution (higher values indicate slower but more cautious responding), and non-decision time (*t*) indexes processes outside the decision itself, such as stimulus encoding and response execution (higher values indicate slower non-decision processes). This framework was useful here because slower or less accurate performance in chronic pain could reflect reduced processing efficiency, greater response caution, slower non-decision processes, or some combination of these. Based on the behavioural analyses, we expected lower drift rate under higher load, and where group differences were present, in the chronic pain group. We also examined whether chronic pain was associated with greater response caution or longer non-decision time.

HDDMs were fitted to trial-level accuracy and RT data using HDDM 0.8.0 in Python. Both correct and incorrect responses were included, and RTs were converted from milliseconds to seconds. For each task, models allowing *v*, *a*, and *t* to vary by group, load, or both were compared, and the final model was selected using the lowest Deviance Information Criterion (DIC). Directional posterior probabilities near 1 or 0 indicate stronger evidence that a parameter differed between groups or load conditions in the tested direction, whereas values closer to 0.5 indicate greater overlap. Full model specification, estimation details, and posterior testing procedures are reported in the Supplementary Materials.

## 4 Results

### 4.1 Participant characteristics

The analysed sample comprised 99 participants with chronic pain and 87 pain-free controls. Baseline characteristics of participants with and without chronic pain, and any between-group differences, are reported in Table 1. Groups did not differ in age or gender identity, but the chronic pain group reported substantially greater mood disturbance, sleep disturbance, and baseline fatigue.

The chronic pain group was clinically heterogeneous. Eleven percent had no formal diagnosis, and the most commonly reported diagnoses were fibromyalgia, chronic back pain, migraine, complex regional pain syndrome, osteoarthritis, irritable bowel syndrome, and hypermobility syndrome. Thirty-six percent met the threshold for widespread pain on the Widespread Pain Index. Detailed pain-location and treatment/medication characteristics are reported in the Supplementary Materials.

Usable sample size varied across tasks after prespecified quality-control procedures; detailed included and excluded counts by task, group, and session are reported in Supplementary Table S1.

### 4.2 Validity and reliability

Preregistered sensitivity and reliability checks are reported in Supplementary Materials. In brief, higher load reduced performance across tasks as expected, validating the sensitivity of the tasks to cognitive load. We also found moderate test–retest reliability for most tasks.

### 4.3 Session 1: Cognitive load and group effects on accuracy and reaction times

To summarise, across all tasks, higher load reduced accuracy and slowed responses in both groups, confirming the expected effect of increasing task demands. There was no evidence of a global working-memory deficit in the chronic pain group. Instead, group differences were task-specific, with the clearest behavioural effect seen in the auditory-temporal task under higher load.

#### 4.3.1 Primary and secondary analyses

Mixed effects models on accuracy and reaction time are summarised in Table 2 and Figure 3. There were consistent main effects of cognitive load across all tasks; regardless of group, participants were less accurate and slower in higher (2-back) compared to lower (0/1-back) cognitive load conditions. In the auditory-temporal task, the chronic pain group showed a steeper load-related decline in accuracy than controls, reflected in a significant Group × Load interaction in the AU task (interaction estimate = −0.447 on the log-odds scale [95% CI −0.579 to −0.314], equivalent to an odds ratio of 0.64 [95% CI 0.56 to 0.73]). Although the chronic pain group was more accurate than controls at low load, this advantage did not generalise to high load; simple-effects tests indicated that participants with chronic pain were more accurate at 1-back (p = 0.012) but tended to be less accurate at 2-back task (p = 0.057, Figure 3C). The AU task also showed greater load-related RT slowing in the chronic pain group (Group × Load estimate = 0.035 on the log-RT scale, 95% CI 0.017 to 0.053, equivalent to about a 3.6% [1.7% to 5.4%] greater load-related slowing), with simple-effects tests showing decrements in chronic pain vs controls in the 2-back task (p = 0.048; Figure 3F). In the visuo-verbal task, the main interaction was on RT, with the chronic pain group showing greater slowing under high load (p < 0.001; Figure 3D; Group × Load estimate = 0.065 on the log-RT scale, 95% CI 0.050 to 0.079, equivalent to about a 6.7% [5.1% to 8.2%] greater load-related slowing). For accuracy, interaction effects were not supported by simple-effect analyses. No comparable pattern was observed in the visuo-spatial task.

**Figure 3.**
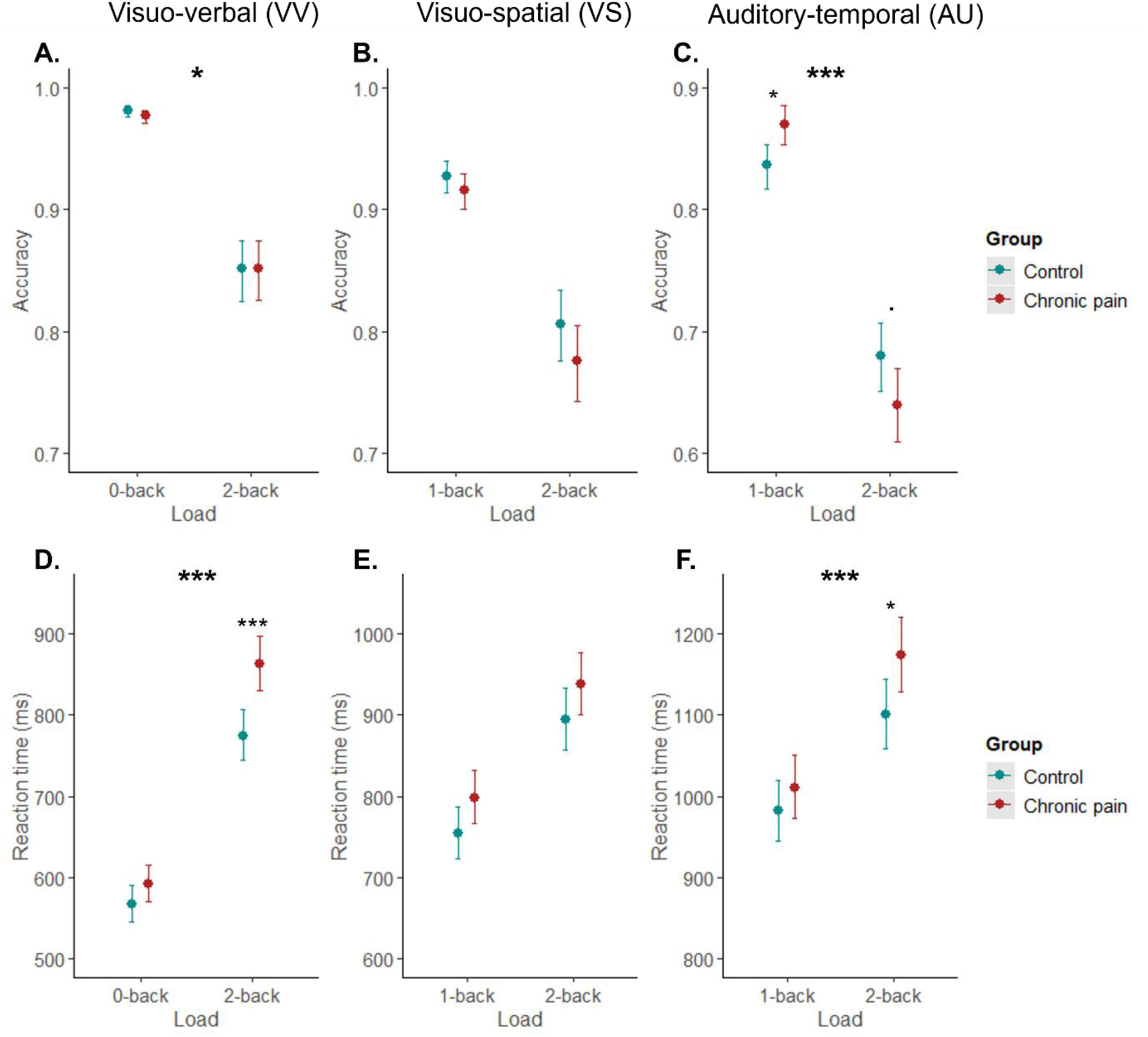
Marginal effects (with 95% confidence intervals) of interaction terms between cognitive load (0/1-back, 2-back) and group (control, chronic pain) on accuracy (A-C) and reaction times (D-F) in each task in session 1. Reaction times have been back-transformed from log-scale for visualisation. Asterisks in bold indicate significant interactions. Asterisks in normal font indicate significant between-group contrasts (after Holm-Bonferroni adjustment). *** p<0.001, * p<0.05,. p<0.1.

**Table 2.**
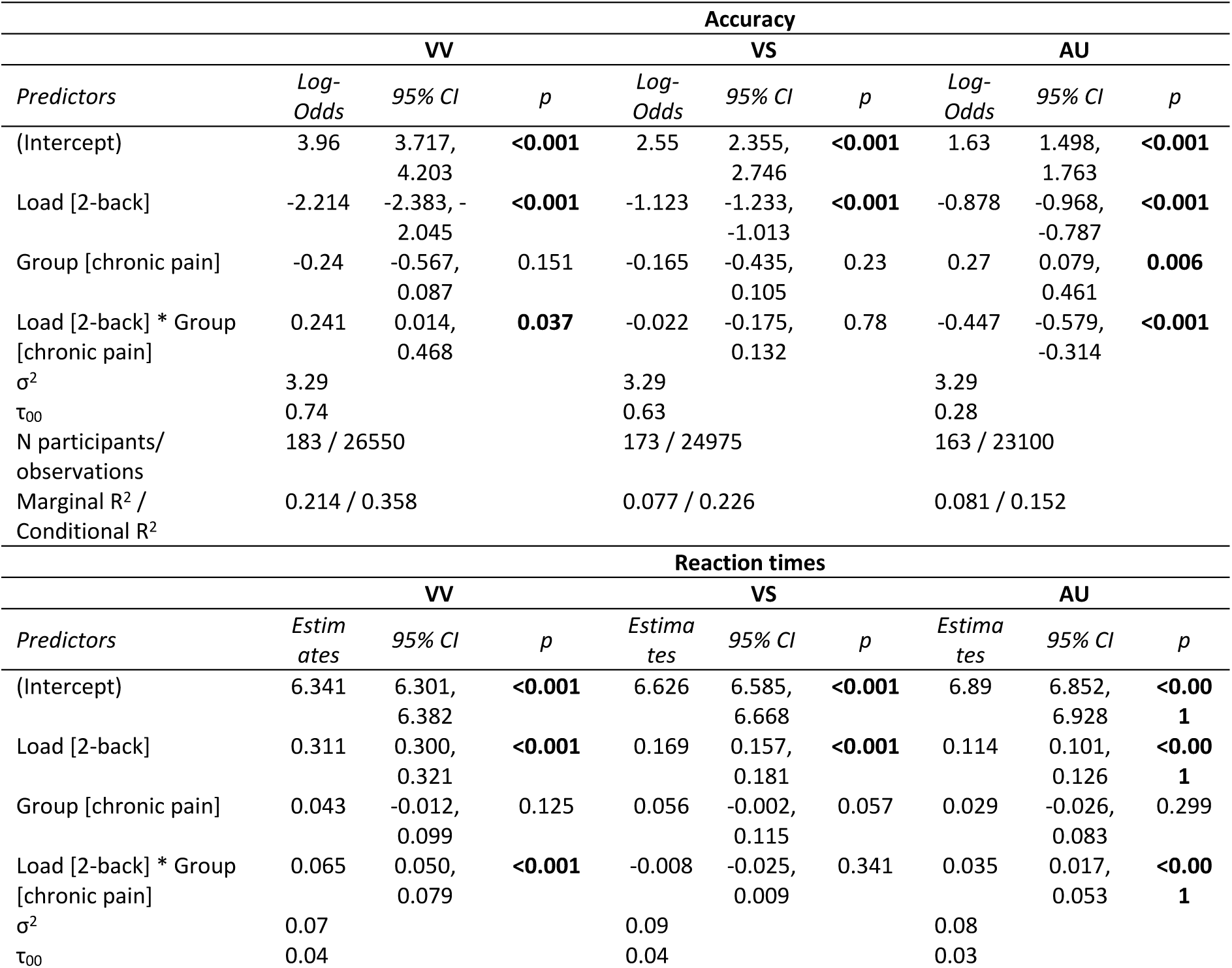
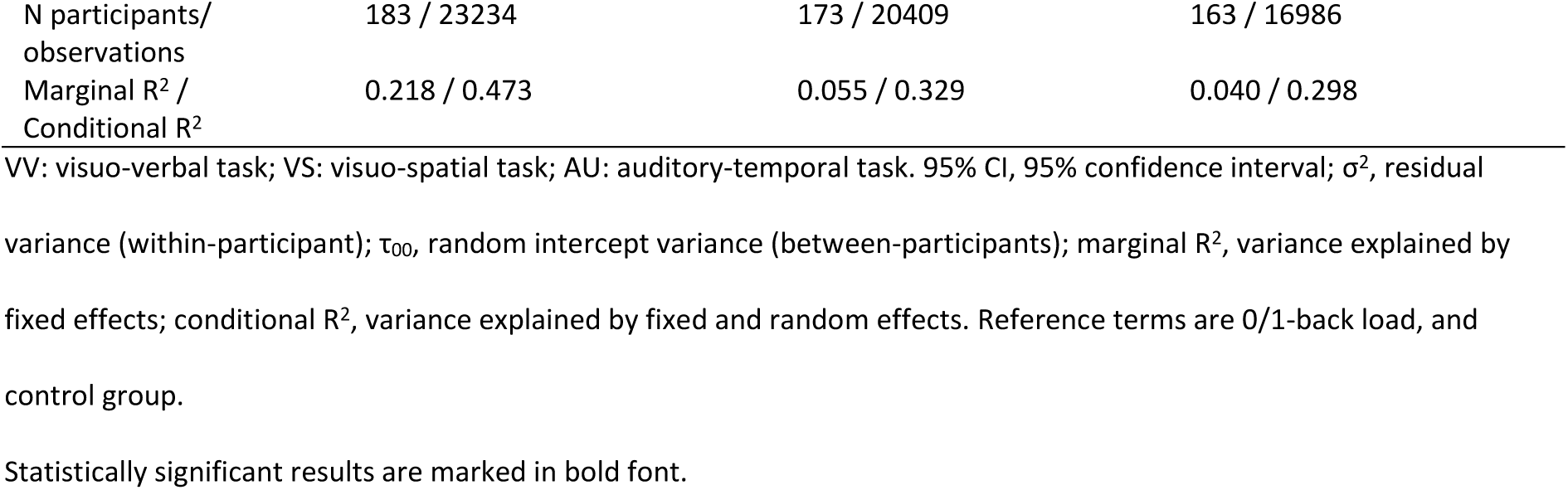
Results of generalised linear mixed models on accuracy and reaction times in each task in session 1.

| Predictors | Accuracy |  |  |  |  |  |  |  |  |
| --- | --- | --- | --- | --- | --- | --- | --- | --- | --- |
|  | VV |  |  | VS |  |  | AU |  |  |
|  | Log-Odds | 95% CI | p | Log-Odds | 95% CI | p | Log-Odds | 95% CI | p |
| (Intercept) | 3.96 | 3.717, 4.203 | **<0.001** | 2.55 | 2.355, 2.746 | **<0.001** | 1.63 | 1.498, 1.763 | **<0.001** |
| Load [2-back] | -2.214 | -2.383, -2.045 | **<0.001** | -1.123 | -1.233, -1.013 | **<0.001** | -0.878 | -0.968, -0.787 | **<0.001** |
| Group [chronic pain] | -0.24 | -0.567, 0.087 | 0.151 | -0.165 | -0.435, 0.105 | 0.23 | 0.27 | 0.079, 0.461 | **0.006** |
| Load [2-back] * Group [chronic pain] | 0.241 | 0.014, 0.468 | **0.037** | -0.022 | -0.175, 0.132 | 0.78 | -0.447 | -0.579, -0.314 | **<0.001** |
| $\sigma^2$ | 3.29 | | | 3.29 | | | 3.29 | | |
| $\tau_{00}$ | 0.74 | | | 0.63 | | | 0.28 | | |
| N participants/ observations | 183 / 26550 |  |  | 173 / 24975 |  |  | 163 / 23100 |  |  |
| Marginal $R^2$ / Conditional $R^2$ | 0.214 / 0.358 | | | 0.077 / 0.226 | | | 0.081 / 0.152 | | |

| Predictors | Reaction times |  |  |  |  |  |  |  |  |
| --- | --- | --- | --- | --- | --- | --- | --- | --- | --- |
|  | VV |  |  | VS |  |  | AU |  |  |
|  | Estimates | 95% CI | p | Estimates | 95% CI | p | Estimates | 95% CI | p |
| (Intercept) | 6.341 | 6.301, 6.382 | **<0.001** | 6.626 | 6.585, 6.668 | **<0.001** | 6.89 | 6.852, 6.928 | **<0.001** |
| Load [2-back] | 0.311 | 0.300, 0.321 | **<0.001** | 0.169 | 0.157, 0.181 | **<0.001** | 0.114 | 0.101, 0.126 | **<0.001** |
| Group [chronic pain] | 0.043 | -0.012, 0.099 | 0.125 | 0.056 | -0.002, 0.115 | 0.057 | 0.029 | -0.026, 0.083 | 0.299 |
| Load [2-back] * Group [chronic pain] | 0.065 | 0.050, 0.079 | **<0.001** | -0.008 | -0.025, 0.009 | 0.341 | 0.035 | 0.017, 0.053 | **<0.001** |
| $\sigma^2$ | 0.07 | | | 0.09 | | | 0.08 | | |
| $\tau_{00}$ | 0.04 | | | 0.04 | | | 0.03 | | |

|  |  |  |  |
| --- | --- | --- | --- |
| N participants/<br>observations | 183 / 23234 | 173 / 20409 | 163 / 16986 |
| Marginal R<sup>2</sup> /<br>Conditional R<sup>2</sup> | 0.218 / 0.473 | 0.055 / 0.329 | 0.040 / 0.298 |

VV: visuo-verbal task; VS: visuo-spatial task; AU: auditory-temporal task. 95% CI, 95% confidence interval; $\sigma^2$ , residual
variance (within-participant); $\tau_{00}$ , random intercept variance (between-participants); marginal R<sup>2</sup>, variance explained by fixed effects; conditional R<sup>2</sup>, variance explained by fixed and random effects. Reference terms are 0/1-back load, and control group.
Statistically significant results are marked in bold font.

In adjusted models including mood disturbance, sleep disturbance, and fatigue, the main behavioural pattern was similar, although VV and AU RT interactions were attenuated. Detailed results from models adjusted for potential confounders are reported in Supplementary Materials.

Since d’ and accuracy outcomes were highly correlated in each task (rs ≥ 0.9) and mixed models showed largely similar patterns for both, we report signal detection metric results in Supplementary Materials.

Overall, these results did not support a generalised across-the-board working memory difficulties in chronic pain. Rather, they indicated that the clearest and most consistent group differences were confined to the auditory-temporal task, with a larger load-related accuracy decrement and greater high-load slowing in the chronic pain group, alongside a more selective high-load slowing effect in the visuo-verbal task.

#### 4.3.2 Follow-up analyses

To test whether any load-dependent differences between groups differed by task-type, we analysed higher-minus-lower load difference scores (Table 3; Figure 4). For accuracy, there was a significant interaction between task type and group, driven by a larger load-related decline in the auditory-temporal task, relative to the visuo-verbal task, in the chronic pain group (task type [AU] × group estimate = −0.079, 95% CI −0.124 to −0.033, that is, about a 7.9 percentage-point larger load-related accuracy decrement in AU relative to VV in the chronic pain group compared with controls). Planned contrasts confirmed that participants with chronic pain showed a greater decline in accuracy from low to high load in the auditory-temporal than visuo-verbal task (p < 0.001), whereas controls did not (p = 1). No corresponding visuo-spatial effect was observed. For RT-difference scores, there was no task type × group interaction. Instead, participants with chronic pain tended to show greater load-related slowing overall (+63ms, 95% CI approximately 0 to 127 ms), while all participants showed greater load-related slowing in the visuo-verbal compared to other tasks (approximately 96 ms and 122 ms smaller RT slowing in VS and AU, respectively, relative to VV; Table 3).

**Figure 4.**
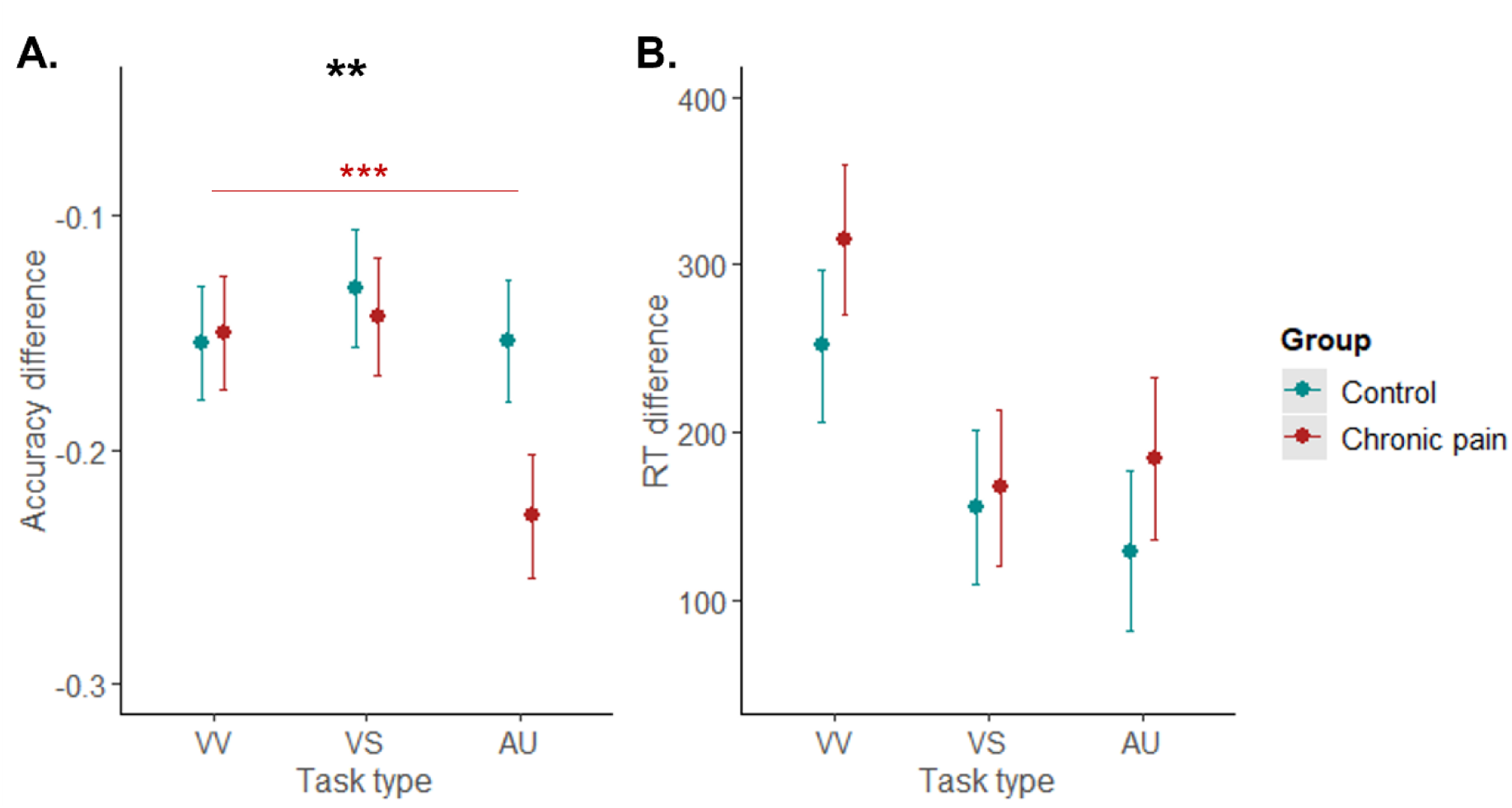
Marginal effects (with 95% confidence intervals) of interaction terms between task type (VV, visuo-verbal; VS, visuo-spatial; AU, auditory-temporal) and group (control, chronic pain) on accuracy difference (A) and reaction time difference scores (B) in session 1. Asterisks in bold indicate significant interaction. Asterisks in normal font indicate significant between-group contrasts (after Holm-Bonferroni adjustment). *** p<0.001, ** p<0.01.

**Figure 5.**
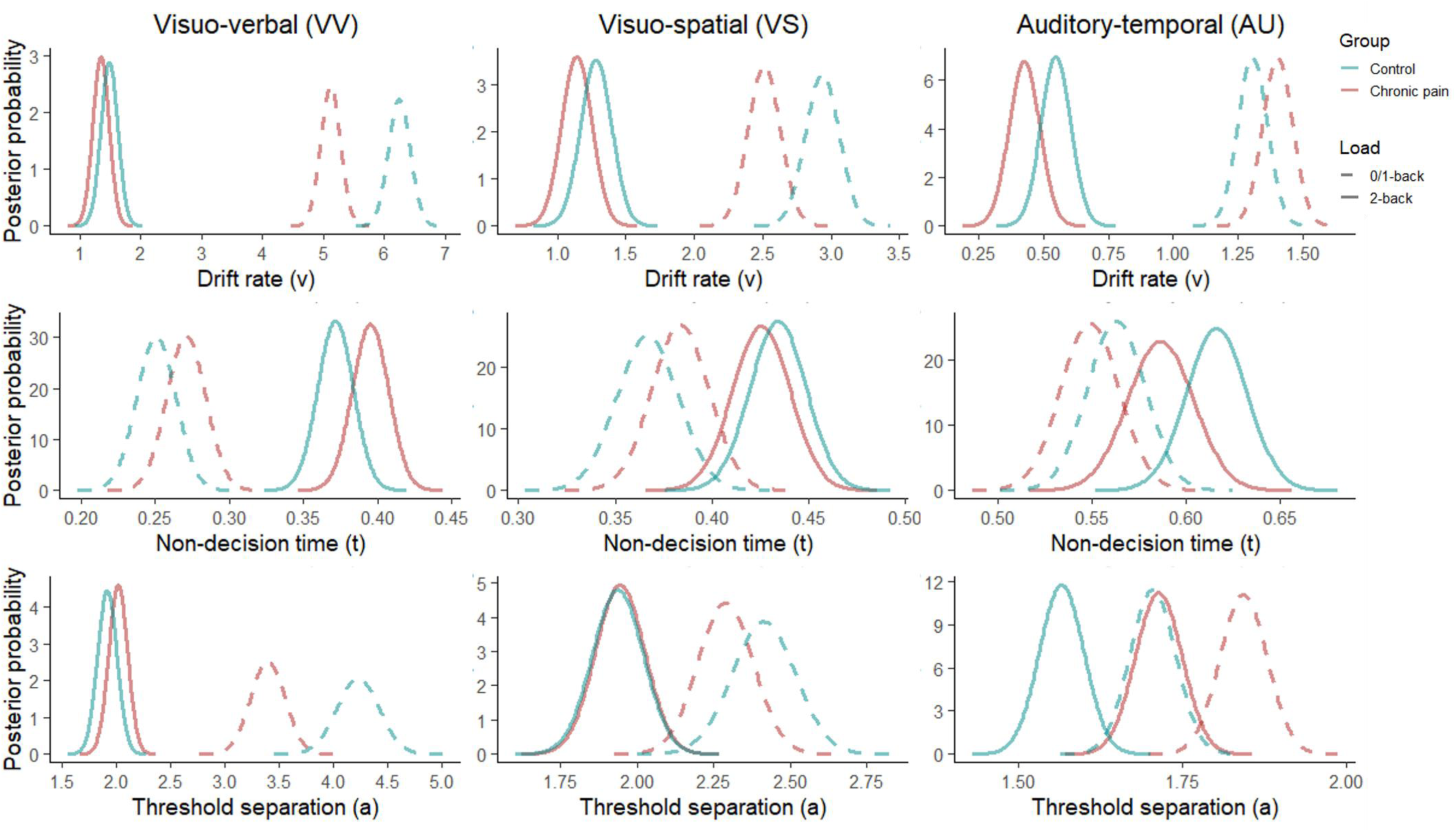
Posterior probability distributions of group (control, chronic pain) and condition (0/1-back, 2-back) means of the HDDM parameters drift rate v (top row), non-decision time t (middle row), and threshold separation a (bottom row), in the visuo-verbal (VV; left column), visuo-spatial (VS; middle column), and auditory-temporal (AU; right column) tasks. Note that the axis limits and intervals are specific to each task and parameter to best visualise the (non-)overlap between posterior distributions.

**Table 3.**
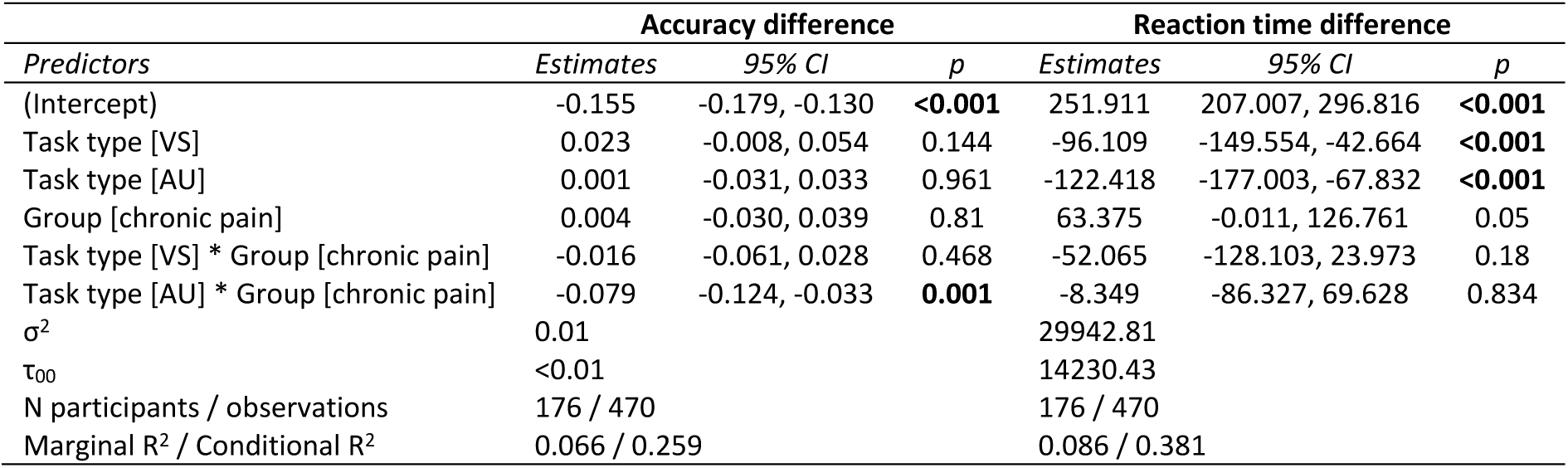
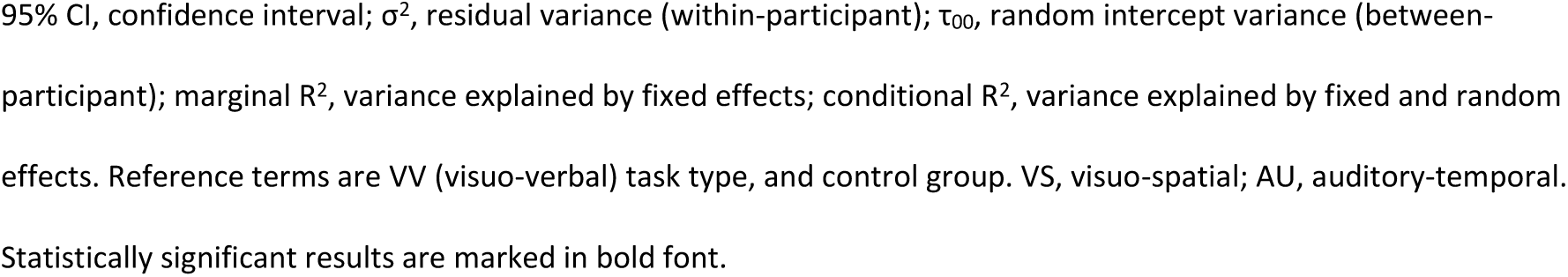
Results of linear mixed models on accuracy and reaction time differences across tasks in session 1.

After adjustment for mood, sleep disturbance, and fatigue, the auditory-specific accuracy pattern remained, whereas the trend-level group difference in RT load cost was attenuated (Supplementary Materials). Preregistered within-group regression analyses in the chronic pain sample are also reported in the Supplementary Materials. In brief, pain duration was not an independent predictor of load-related performance change in any task.

### 4.4 Session 2 replication

Supplementary Materials provide the detailed results from session 2. In brief, similar patterns are observed to those observed in session 1. Higher load again impaired performance across tasks, and the session-1 auditory-temporal pattern was not idiosyncratic to a single session. Some individual interactions differed between sessions, particularly for visuo-verbal and visuo-spatial RTs, but adjusted and unadjusted session-2 results were generally consistent with the main session-1 findings.

### 4.5 Model-based exploratory analyses: HDDMs

For each task, models allowing drift rate, threshold separation, and non-decision time to vary by group, load, or both were compared, and the winning model was selected using the lowest DIC (see Supplementary Materials). In all three tasks, the full model provided the best fit, and posterior predictive checks indicated that these models reproduced the main patterns in the observed data. However, simplified models that attributed more of the pattern to cognitive load alone often fit nearly as well, suggesting that load effects were generally stronger and more consistent than group effects.

Hypothesis testing was performed on the posteriors (see exact probabilities of the differences reported in Table 4). Across all tasks, higher load was associated with lower drift rate, longer non-decision time, and lower threshold separation, indicating that increasing task demands reduced evidence-accumulation efficiency, slowed non-decision processes, and was associated with less cautious responding. Group effects were more selective. There was no convincing evidence of longer non-decision time in the chronic pain group (vs controls) in any task or under any load. In the auditory-temporal task, the chronic pain group showed greater threshold separation than controls across both load conditions, consistent with greater response caution. However, the chronic pain group were less cautious than controls in the VV task under low load, with no difference in the VV high-load task or VS tasks. Drift-rate differences were modest and task-specific, with lower drift rate in the chronic pain group (vs controls) in low-load visual conditions (VV and VS) and only borderline evidence for a group difference in the high-load AU condition.

**Table 4.** Probabilities that the posterior distributions of the v, t, and a parameters differ between the groups or load conditions in the tested directions.

|  | VV |  |  |  | VS |  |  |  | AU |  |  |  |
| --- | --- | --- | --- | --- | --- | --- | --- | --- | --- | --- | --- | --- |
| | | | | $P$ | | | | $P$ | | | | $P$ |
| Drift rate ($v$) | v_control_VV0 | > | v_control_VV2 | **1.00** | v_control_VS1 | > | v_control_VS2 | **1.00** | v_control_AU1 | > | v_control_AU2 | **1.00** |
|  | v_control_VV0 | > | v_chronic_pain_VV0 | **1.00** | v_control_VS1 | > | v_chronic_pain_VS1 | **1.00** | v_control_AU1 | > | v_chronic_pain_AU1 | 0.10 |
|  | v_control_VV2 | > | v_chronic_pain_VV2 | 0.77 | v_control_VS2 | > | v_chronic_pain_VS2 | 0.81 | v_control_AU2 | > | v_chronic_pain_AU2 | 0.95 |
|  | v_chronic_pain_VV0 | > | v_chronic_pain_VV2 | **1.00** | v_chronic_pain_VS1 | > | v_chronic_pain_VS2 | **1.00** | v_chronic_pain_AU1 | > | v_chronic_pain_AU2 | **1.00** |
| Non-decision time ($t$) | t_control_VV2 | > | t_control_VV0 | **1.00** | t_control_VS2 | > | t_control_VS1 | **1.00** | t_control_AU2 | > | t_control_AU1 | **0.99** |
|  | t_chronic_pain_VV0 | > | t_control_VV0 | 0.89 | t_chronic_pain_VS1 | > | t_control_VS1 | 0.79 | t_chronic_pain_AU1 | > | t_control_AU1 | 0.26 |
|  | t_chronic_pain_VV2 | > | t_control_VV2 | 0.92 | t_chronic_pain_VS2 | > | t_control_VS2 | 0.34 | t_chronic_pain_AU2 | > | t_control_AU2 | 0.13 |
|  | t_chronic_pain_VV2 | > | t_chronic_pain_VV0 | **1.00** | t_chronic_pain_VS2 | > | t_chronic_pain_VS1 | **0.97** | t_chronic_pain_AU2 | > | t_chronic_pain_AU1 | **0.96** |
| Threshold separation ($a$) | a_control_VV2 | > | a_control_VV0 | **0.00** | a_control_VS2 | > | a_control_VS1 | **0.00** | a_control_AU2 | > | a_control_AU1 | **0.00** |
|  | a_chronic_pain_VV0 | > | a_control_VV0 | **0.00** | a_chronic_pain_VS1 | > | a_control_VS1 | 0.16 | a_chronic_pain_AU1 | > | a_control_AU1 | **1.00** |
|  | a_chronic_pain_VV2 | > | a_control_VV2 | 0.84 | a_chronic_pain_VS2 | > | a_control_VS2 | 0.53 | a_chronic_pain_AU2 | > | a_control_AU2 | **1.00** |
|  | a_chronic_pain_VV2 | > | a_chronic_pain_VV0 | **0.00** | a_chronic_pain_VS2 | > | a_chronic_pain_VS1 | **0.00** | a_chronic_pain_AU2 | > | a_chronic_pain_AU1 | **0.01** |

$P$ , probability values close to 1 indicate that the parameter in one group or condition differs from the other group or condition in the tested direction (i.e. posterior distributions of this
parameter in the two groups or conditions do not significantly overlap), whereas values close to 0 indicate the difference in the opposite than the tested direction. $P$ s >0.95 or <0.05 are
marked in bold as indicating significant differences.

Overall, the HDDM findings broadly mirrored the behavioural analyses: cognitive load had robust effects across tasks, whereas group differences were smaller and more selective, with the clearest chronic-pain-related difference in the auditory-temporal task being greater response caution rather than consistently slower non-decision processes or a general reduction in evidence-accumulation efficiency.

## 5 Discussion

We examined whether people living with chronic pain differ from pain-free controls on three online tasks (visuo-verbal, visuo-spatial and auditory-temporal) at low and high working-memory load. There was no evidence of global working-memory differences in this mixed chronic pain sample. Higher load reduced accuracy and slowed responses in both groups, but the clearest group difference was a larger load-related accuracy decrement in the auditory-temporal sequence-matching task. Greater high-load slowing in visuo-verbal and auditory-temporal tasks was attenuated after adjustment for mood, sleep disturbance, and fatigue (suggesting these factors may contribute to load-sensitivity), whereas the auditory-temporal task accuracy effect persisted.

However, the auditory-temporal finding should be interpreted cautiously. It is important to note that the auditory-temporal task did not isolate auditory or temporal processing alone, but combined auditory-temporal sequence comparison with working-memory updating, sustained attention, and time pressure. Low-load auditory accuracy was not impaired, arguing against a primary auditory sensory deficit. Within that task, drift-diffusion modelling indicated greater threshold separation (“response caution”) in the chronic pain group, with no convincing group difference in non-decision time (“sensory/motor slowing”). Drift-rate differences were smaller and largely confined to low-load visual conditions. The absence of broad group differences in drift rate or non-decision time argues against a generalised slowing account or a global working-memory capacity deficit. Overall, the results are more consistent with greater response caution in the auditory-temporal task, and this may have contributed to poorer high-load accuracy in that condition, potentially under the time constraints of the task.

These findings may help explain why the literature on working memory in chronic pain has been mixed (Berryman et al., 2013; Kelly et al., 2025). Rather than showing a broad reduction in performance across all tasks, group differences were selective and depended on task characteristics. The clearest effect emerged in the higher-load auditory-temporal task, whereas visual task effects were smaller and mainly limited to low-load conditions. This pattern suggests that at least some previously reported “deficits” may reflect task-specific demands and strategic adjustment under uncertainty, rather than a global loss of working-memory capacity. The small adjusted visuo-spatial effect remains of interest given prior links between chronic pain and functions associated with parietal systems (such as spatial maintenance and manipulation (Bray et al., 2015)), but in the present data it was not load-sensitive, so it should be interpreted cautiously. Because accuracy and mean reaction time conflate working-memory demands with vigilance, processing speed, pacing, and response strategy, diffusion modelling helps provide a more specific account of the processes contributing to these task-specific group differences.

Strengths of the study include preregistration, the multimodal design, the use of both lower- and higher-load conditions, and the combination of trial-level behavioural analyses with diffusion modelling. Measuring mood disturbance, sleep disturbance, and fatigue was also important, as these factors differed markedly between groups and partly accounted for load-related slowing. Together, these design features helped distinguish broad performance differences from more task-specific effects. Lastly, the consistency of the pattern of results between session 1 and session 2 suggests that the main auditory-temporal finding was not a single-session artefact, although the retest sample was smaller and should not be treated as an independent replication.

The study also has important limitations. The chronic pain group was deliberately broad and clinically heterogeneous, including participants with different diagnoses, pain distributions, and treatment histories, so the findings should not be interpreted as characterising all chronic pain conditions uniformly. The sample was relatively young and community-based, and education, race/ethnicity, and more detailed medication burden were not available as covariates. Medication use was characterised descriptively, but not modelled in the main case-control analyses. Online testing increased reach but introduced uncontrolled hardware and environmental variation, particularly relevant to the auditory task, despite headphone checks and other mitigation steps. As data were collected online during the COVID-19 period (March to October 2021), broader contextual influences on stress, routine, and daily functioning may limit the generalisability of the findings to non-pandemic settings. The study was powered for the main case-control comparison, not for fine-grained subgroup analyses or every secondary/exploratory contrast, so smaller or isolated effects should be interpreted cautiously. In addition, usable sample size varied across tasks after quality-control procedures, and drift-diffusion analyses were not covariate adjusted (adding covariates would substantially increase model complexity), so symptom-related factors may still have contributed to some of the smaller visual task effects.

Future work should expand beyond visual working-memory probes to include a wider range of cognitive tasks, including auditory-temporal paradigms such as the one used here, especially where these help distinguish response caution from processing efficiency. It will also be important to test whether the present auditory-temporal pattern replicates in more controlled settings and in more clearly phenotyped pain samples. Stratified recruitment may be needed because working-memory differences could vary across nociceptive, nociplastic and neuropathic presentations (Kelly et al., 2025), and may also differ according to demography, pain severity, and pain duration. Age and gender were broadly comparable between groups in our study and were therefore not included as preregistered covariates, but larger studies are needed to test whether task-specific effects vary across demographic subgroups.

Repeated-measurement designs, together with improved hearing and medication characterisation, will be important for clarifying whether altered response caution reflects consequences of pain, premorbid vulnerability, or both. For example, longitudinal studies in well-characterised high-risk samples, such as post-surgical cohorts (Giusti et al., 2020; Sluka et al., 2023), could test whether performance on specific tasks predicts pain persistence, whereas family or twin designs (Rader et al., 2025) could help separate shared liability from pain-contingent change. In the present data, pain duration did not emerge as an independent predictor of load-related performance change, which is more consistent with a state-related than a simple chronicity account, although this should be interpreted cautiously. Repeated measurements depend on reliable outcomes, and in our data retest indices were generally moderate, with the least stable likely affected by ceiling effects, so reliability may improve further in more controlled settings. Future studies should also continue to measure symptoms such as sleep disturbance, fatigue, and mood, given their likely contribution to variation in working-memory performance.

In conclusion, the present findings do not support a global working-memory problem in chronic pain. Instead, they suggest a more selective pattern, with the clearest group difference emerging in auditory-temporal working memory at higher cognitive load. Diffusion modelling indicated that this pattern was more consistent with greater response caution than with general sensory-motor slowing, while mood disturbance, sleep disturbance, and fatigue appeared to contribute to some of the load-related slowing effects. Taken together, these findings suggest that mixed results in the chronic-pain cognition literature may partly reflect task-specific demands and the use of broad behavioural measures that do not distinguish underlying processes. Future work should therefore combine a wider range of cognitive tasks, including auditory-temporal probes, with clearer phenotyping and measurement of relevant symptoms to test prognosis, mechanism, and clinical relevance more directly.

## 6. Acknowledgements

We thank Emily Sutton, Verity Wild, and Grace O’Hara (University of Liverpool) for assistance with piloting the online tasks, supporting recruitment activities (including dissemination of study advertisements), and study administration on the Gorilla platform (including monitoring participant progress and supporting data management procedures).

## 7 Author Contributions

MH: Conceptualisation, Methodology, Investigation, Data curation, Formal analysis (behavioural and drift diffusion modelling), Software, Visualisation, Writing (original draft, review and editing).

MS: Formal analysis (drift diffusion modelling), Software, Interpretation (computational findings), Writing (review and editing).

CB: Conceptualisation, Funding acquisition, Supervision, Project administration, Interpretation, Writing (original draft, review and editing).

All authors approved the final manuscript and agree to be accountable for all aspects of the work.

## 8 Disclosures

### 8.1 Research funding

This research was funded via the Translational Research Access Programme (TRAP), Faculty of Health & Life Sciences, University of Liverpool, UK. The funding body had no role in the collection, analysis, or interpretation of data, or preparation of the manuscript.

### 8.3 Data and materials availability

The study was preregistered on the Open Science Framework (OSF): https://doi.org/10.17605/OSF.IO/TH7DV. The preregistration specifies the planned sample size, primary hypotheses, exclusion criteria, and analysis plan.

De-identified participant-level data (analysis-ready files used for the reported analyses) and the R scripts used for the primary statistical analyses will be made publicly available on the Open Science Framework (OSF) upon acceptance for publication: https://doi.org/10.17605/OSF.IO/TH7DV. The OSF record currently contains the preregistration, and the data/code components will be added as part of the final publication workflow.

The Gorilla task components (for the individual n-back tasks) are available as “open materials” (VV: https://app.gorilla.sc/openmaterials/1178690; VS: https://app.gorilla.sc/openmaterials/1178693; AU: https://app.gorilla.sc/openmaterials/1178694). Other study materials will be shared on OSF upon acceptance for publication: https://doi.org/10.17605/OSF.IO/TH7DV. This will include task parameters and stimuli lists, questionnaires, and scoring/processing scripts, as well as drift diffusion analysis-ready model outputs used to generate the reported results.

