## Supplementary Materials for "Working memory in chronic pain: evidence for task-specific rather than global differences"

### 1 Methods

#### 1.1 Pre-registered research questions and hypotheses

RQ1. Are the newly developed online tests valid and reliable measures of sustained attention and working memory in individuals with and without chronic pain?

H1a. Participants’ performance on the sustained attention and low cognitive load working memory tasks will be strongly associated (validity).

H1b. Participants’ performance on low cognitive load tasks will deteriorate throughout the duration of the experiment (validity).

H1c. Participants’ performance will be worse in the tasks with higher working memory load compared to the tasks with lower working memory load (validity).

H1d. Participants’ performance on the same tasks in the baseline and retest sessions will be in strong agreement (test-retest reliability).

RQ2. Are there any domain-specific and/or domain-general deficits in sustained attention and working memory performance in individuals with chronic pain compared to healthy participants?

H2a. Participants with chronic pain will have worse performance on all tasks compared to healthy participants, indicating domain-general deficits.

H2b. Any differences in performance of participants with and without chronic pain will be more pronounced in the tasks characterised by higher cognitive load.

H2c. Any differences in performance of participants with and without chronic pain will be more pronounced in domain-specific tasks that rely to a greater extent on the function of the parietal lobes, compared to domain-general tasks that rely mainly on the function of the frontal lobes.

RQ3. Is the performance on sustained attention and working memory tasks associated with chronic pain symptoms?

H3a. Longer disease duration will be associated with worse performance on the tasks requiring frontal and parietal functions.

#### 1.2 **Cognitive tasks – procedural, stimulus and timing details**

There were six task conditions across three task modalities: Visuo-verbal (VV) 0-back and 2-back, visuo-spatial (VS) 1-back and 2-back, and auditory-temporal (AU) 1-back and 2-back. Each consisted of 75 trials, including 23 target trials. Among the non-target trials, there were 4 lure trials in each task condition (i.e. repetition of 1-back stimulus in 0-back task, repetition of 2-back stimulus in 1-back task, or repetition of 1-back or 3-back stimulus in 2-back task). Every task type included nine unique stimuli, each appearing 8-9 times in every task condition, except for the 0-back task, where the target letter appeared 23 times, and every other letter 6-7 times. Because of the dependency across trials specific to n-back tasks, the order of the stimuli was hardcoded. However, to control for possible order effects, in each task condition participants were randomised to one of two possible fixed stimulus orders. Participants were instructed to be as accurate and as fast as possible, and respond to both target and non-target trials using different response keys. Each stimulus was presented for 200 ms (except for the auditory stimuli, where presentation time varied according to stimulus frequency) and each trial ended after ~3200 (+/- 50) ms from stimulus onset.

**Visuo-verbal** stimuli were nine consonants (B, G, H, K, N, R, S, X, Z) presented in white Calibri font (size 124; 2.8 cm high, in the centre of a 5 x 5 cm zone) on grey background, similar to the fixation cross. Their actual size was scaled according to the screen size so that the letters were contained in a zone spanning approximately 39% of the screen height and 36% of the screen width.

In **visuo-spatial** tasks, an empty white 3 x 3 square grid spanning approximately 43% of the screen height and 36% of the screen width was presented on grey background instead of a fixation cross. The stimuli consisted of the same grid, but with two out of nine squares filled in white, forming nine unique patterns.

**Auditory-temporal** stimuli consisted of a sequence of 10 tones (each 400 Hz) played at different frequencies, ranging from 10Hz to 43Hz, with frequency increasing in *1.2 steps between consecutive stimuli (i.e., full range of stimuli included 10, 12, 14, 17, 21, 25, 30, 36, and 43 Hz). The duration of each sound therefore ranged from 233 (43 Hz) to 1000 (10 Hz) ms. These stimulus properties were selected based on piloting with healthy participants to ensure that they were able to easily discriminate different stimulus frequencies. To further aid the discrimination, the stimuli during the tasks were ordered so that there was at least a 2-step difference in frequency between consecutive (1-back) non-target trials (e.g., 10 and 14 Hz stimuli could be presented after each other, but 10 and 12 Hz could not). Additionally, in the 2-back task condition, there was at least a 2-step difference between 2-back non-target trials (e.g., a sequence of 17, 25, and 12 Hz could be presented, but a sequence of 17, 25, and 14 Hz could not, because there would be only 1-step difference between 17 and 14 Hz). Only a white fixation cross on grey background was displayed on the screen during the AU tasks.

After each task condition and completing the post-experiment survey, participants were presented with their number of correct responses (out of 75). No feedback was provided on a trial basis. Each task condition was preceded by 10 training trials (including 3 target stimuli), which had the same structure as the actual task, with addition of a ‘Get ready’ prompt before the first stimulus (starting 300ms from task onset and remaining on screen for 1000ms), a ‘Respond now’ prompt (starting 300ms after stimulus onset and remaining on screen for 800ms) to train participants to respond on every trial, not only the target trials, and feedback (a tick mark for correct, and a cross for erroneous responses) displayed on screen for 200 ms upon response. At the end of the training block, participants were presented with their number of correct responses (out of 10) and were given an option to repeat the training once or continue to the task. Participants were allowed to skip the task if after repeated training they could not understand the instructions.

#### 1.3 **Statistical analysis**

##### 1.3.1 Validity and reliability testing

We preregistered four validity/reliability checks:

1. Worse performance at higher load (a sensitivity check).
2. Strong test–retest agreement (a reliability check).
3. A strong association between the low-load n-back and the 0-back vigilance task, treated as a convergent validity index. In hindsight, this indexes construct overlap rather than validity in the strict psychometric sense and we therefore report them transparently as exploratory convergent/discriminant checks rather than a validity test.
4. 4. Within-session deterioration on low-load tasks as a proxy for fatigue sensitivity. In hindsight, this indexes fatigue sensitivity rather than validity, without interpreting as a validity test.

We estimated intraclass correlation coefficients (ICC function in *psych* package (Revelle, 2021) between participants’ performance on 0-back visuo-verbal task, 1-back visuo-spatial task, and 1-back auditory-temporal task (using two-way model, consistency, single measurement form, i.e. ICC3; (McGraw & Wong, 1996), which should be high if low working memory load tasks performance relies largely on sustained attention (H1a). To test whether performance deteriorated throughout the experiment (H1b), we compared participants’ data on the first and last completed 0/1-back task (regardless of the task type). H1c that performance declines with increasing working memory load would be evidenced by the main effect of cognitive load in the mixed effects models as specified in the main Methods section. To assess test-retest reliability of each task (H1d), we estimated the ICC between participants’ performance on each task in the first session and in the second session (using two-way model, absolute agreement, single measurement form, i.e. ICC2; McGraw & Wong, 1996). ICCs < 0.5 would indicate poor, 0.5 - 0.75 moderate, 0.75 - 0.9 good, and > 0.90 excellent reliability.

We explored whether transient pain intensity, mood disturbance, fatigue, and/or sleep could explain any intra-individual differences in sustained attention and working memory performance of participants with chronic pain across two sessions by correlating changes in these variables between sessions with changes in task performance metrics between sessions.

##### 1.3.2 **Signal Detection Theory metrics**

In addition to the measures outlined in the main Methods section, accuracy data from the tasks were quantified using Signal Detection Theory: sensitivity (discriminability) index d’, which is a signal detection metric calculated as z(hit rate) – z(false alarm rate), where z is the inverse of the standard normal cumulative distribution, hit rate is the number of correctly identified targets divided by the total number of targets, and false alarm rate is the number of non-targets erroneously identified as targets divided by the total number of non-targets (dprime function in *psycho* package; (Makowski, 2018; Pallier, 2002). Linear mixed models were fitted to d’ participant-level data using methods detailed in the main Methods section. In addition, difference in performance (d’) between 0/1-back and 2-back versions of each task (i.e. visual, visuo-spatial, and auditory-temporal) was the primary outcome of regression models to assess predictors of working memory performance.

We calculated another signal detection metric called response bias or criterion (beta), as -1/2 * (z(hit rate) + z(false alarm rate)). Higher values would indicate more conservative responding, whereas lower values would indicate more liberal responding. We explored between-group differences on this metric as per planned analyses, and fitted linear mixed models to log-transformed beta outcomes.

##### 1.3.3 **Included and excluded participant data**

Table S 1 presents the number of participants included in the analysis and those excluded due to insufficient numbers of responded-to trials and no-better-than-chance accuracy.

Table S 1. Number of participants included in analysis per task, session, and group.

| Task | Group | | | | | | | |
| --- | --- | --- | --- | --- | --- | --- | --- | --- |
|  | Patients | | | | Controls | | | |
|  | Session 1 | | Session 2 | | Session 1 | | Session 2 | |
|  | Included | Excluded | Included | Excluded | Included | Excluded | Included | Excluded |
| VV0 | 96 | 2 | 75 | 4 | 86 | 0 | 74 | 2 |
| VV2 | 85 | 11 | 75 | 3 | 85 | 2 | 75 | 1 |
| VS1 | 87 | 12 | 72 | 7 | 83 | 3 | 75 | 0 |
| VS2 | 81 | 15 | 73 | 6 | 80 | 5 | 71 | 4 |
| AU1 | 79 | 21 | 72 | 6 | 81 | 5 | 70 | 7 |
| AU2 | 72 | 25 | 66 | 12 | 76 | 9 | 71 | 6 |

VV0: visuo-verbal 0-back task; VV2: visuo-verbal 2-back task; VS1: visuo-spatial 1-back task; VS2: visuo-spatial 2-back task; AU1: auditory-temporal 1-back task; AU2: auditory-temporal 2-back task.

There were some statistically significant differences between participants with complete data and those with at least one task data removed: participants with chronic pain with removed data were older (M = 43.83; SD = 11.99) compared to those with complete included data (M = 37.03; SD = 11.38; Welch’s t(81) = -1.19, p = .006); more commonly reported regular use of NSAIDs (37% vs. 24%), strong opioids (24% vs. 12%), antidepressants (37% vs. 22%), and less often used regular physical therapies (22% vs. 41%) for pain relief; more frequently reported a diagnosis of chronic primary back pain (15% vs. 2%), osteoarthritis (22% vs. 5%), fibromyalgia (27% vs. 19%), and radiculopathy (7% vs. 0%), and less frequently chronic low back pain (17% vs. 22%), tension headache (0% vs. 7%), and migraine (10% vs. 19%).

Overall, 87 control participants and 99 participants with chronic pain completed the first study session (186 in total), and 81% of those, i.e., 77 control participants and 77 participants with chronic pain completed the retest session (154 in total). The average number of days between the two sessions was longer for participants with (M = 11.09; SD = 4.26) than without chronic pain (M = 8.60; SD = 2.91; Welch’s t(134) = -4.22, p < .001). There were no significant differences on baseline characteristics between the participants who did and who did not complete the retest session, except for some differences in pain severity, medication and diagnoses. Completing participants with chronic pain had lower BPI score (M = 4.63; SD = 2.05) than non-completing participants with chronic pain (M = 6.01; SD = 1.46; Welch’s t(48) = 3.53, p = .001), greater proportion of non-completing participants reported regular use of NSAIDs (45% vs. 25%), mild (18% vs. 10%) and strong opioids (27% vs. 14%), antiepileptics (36% vs. 12%), antidepressants (36% vs. 26%), and topical analgesics (15% vs. 4%), but smaller proportion of physical therapies (14% vs. 39%) for pain relief; and greater proportion of non-completing participants was diagnosed with chronic back pain (32% vs. 17%), chronic primary back pain (14% vs. 5%), osteoporosis (32% vs. 6%), irritable bowel syndrome (18% vs. 10%), and pain after nerve injury (18% vs. 5%).

##### 1.3.4 Predictors of performance: Within-group regression analyses

To examine whether longer pain duration, independent of other participant characteristics, was associated with worse task performance (differences in accuracy and RTs between 0/1-back and 2-back condition of each task), we used multiple linear regression models including the following pre-specified predictors: log-transformed pain duration (months), pain severity (BPI score), mood disturbance (POMS score), baseline fatigue, sleep disturbance, regular opioid use (yes, no), and age (years). We considered using current pain and fatigue ratings given after completing each task (as outcomes were analysed as difference scores, these would be averaged ratings after 0/1-back and 2-back condition of the same task type), however, these current ratings were highly correlated with BPI pain severity (rs 0.79-0.81) and baseline fatigue (rs 0.59-0.71). Considering that the BPI is a validated and standardised measure, and for consistency with the previous analyses using baseline fatigue as a covariate, we included BPI pain severity and baseline fatigue in the current regression models. Furthermore, RT variability in the 0-back visual task was the outcome of a regression model to assess predictors of sustained attention performance.

##### 1.3.5 HDDM parameters and model fitting details

In the drift diffusion framework, decision-making in two-choice tasks is operationalised as an evidence accumulation process until one of the two possible choice boundaries is reached (Ratcliff & Rouder, 1998). In the current working memory tasks, a decision whether the current stimulus matches the stimulus presented on n-back trial (target or non-target) required retrieval of stimuli presented in the previous trials from working memory. Based on participants’ accuracy, mean RTs, and RT distributions, several parameters corresponding to different underlying processes can be estimated (Ratcliff & Rouder, 1998; Voss et al., 2004).

Drift rate (*v*) represents the speed of the evidence accumulation process, i.e., the rate at which it approaches a response boundary. It is thought to reflect the efficiency of cognitive processing and depend on the quality of information, e.g., drift rate would be higher when discriminating easy stimuli with high signal-to-noise ratio compared to more difficult or noisy stimuli. In the context of memory, drift rate has been interpreted as indexing the quality of the match between the test stimulus and the memory trace affecting the memory evidence accumulation process (Ratcliff & McKoon, 2008), and it has been found to decrease with increasing working memory load (Shepherdson et al., 2018; Thurm et al., 2018). Thus, within the present study, drift rate is interpreted as evidence accumulation efficiency during working-memory performance, and following our primary predictions, it should be lower in high compared to low load conditions, in participants with chronic pain compared to control participants, and any decrease in drift rate in chronic pain group should be more apparent in high than low load conditions.

Threshold separation (*a*) is the distance between two boundaries, which determines the amount of evidence that needs to be accumulated before a decision is reached. This component is thought to quantify the degree of speed-accuracy trade-off, where a higher threshold corresponds to more cautious (slower but more accurate) responses, and a lower threshold corresponds to more impulsive (faster but less accurate) responses. Since individuals with chronic pain are often aware of their memory difficulties (K. S. Baker et al., 2018), it would be conceivable that they might adapt a more cautious responding, in particular under higher cognitive load.

Non-decision time (*t*) represents the duration of stimulus encoding and response execution processes. As decreased processing and psychomotor speed have been frequently reported in chronic pain (Higgins et al., 2018), we would expect this group to have longer non-decision time than controls.

HDDM uses Bayesian methods to estimate the posterior distributions of these parameters (thus providing measures of their uncertainty) simultaneously at group and participant levels, where participant-level parameter estimates are constrained by the group-level distributions they are drawn from (Wiecki et al., 2013).

HDDMs were fitted to trial-level accuracy and RT data using HDDM 0.8.0 package for Python (version 3) (Wiecki et al., 2013). In contrast to the mixed models analyses, RTs of both correct and incorrect responses were used, and were transformed from milliseconds to seconds. Participants’ responses were coded according to the stimulus-coding, i.e., corresponded to categorisation of the stimulus as a non-target (0, lower boundary) or a target (1, upper boundary). For every model, we drew 5000 samples and discarded the first 500 samples as burn-in.

For each task, to find the model that best explained the data, we first fitted a reference null model (with *v*, *t*, and *a* fixed), and a full model (where *v*, *t*, and *a* were allowed to vary by group and cognitive load). Next, in step 1, we fitted models where *v* and *a* could each vary by group, load, or both group and load (while *t* remained fixed), since these were the two parameters of interest related to cognitive processing. In step 2, we took the best model from step 1 and tested whether varying *t* by group, load, or both factors improved the step 1 model. Bias parameter (*z*), which reflects the starting point of the drift process (equidistant or closer to one of the boundaries) remained fixed across the tested models. This was because the ratio of targets to non-targets was the same across both load conditions, both groups of participants completed the same tasks and in counterbalanced order, and there were no grounds to expect that any pre-existing bias would differ between groups or load conditions. The final model selection for each task was based on the lowest Deviance Information Criterion (DIC) (Spiegelhalter et al., 2002).

The hypothesis tests on the posterior distributions of each parameter (according to the winning model structure) examined whether drift rate is greater in the control than chronic pain group, and greater in the low than high load condition; non-decision time is greater (longer) in the chronic pain than control group, and greater in the high than low load condition; threshold separation is greater (wider) in the chronic pain than control group, and greater in the high than low load condition.

Probability values obtained from the hypothesis tests represent the proportion of the posteriors in which the parameter for one condition or group is greater than the other group or condition. Values close to 1 or 0 would indicate that the posterior distributions of this parameter in the two groups or conditions do not significantly overlap. Values close to 1 would indicate the difference in the tested direction whereas values close to 0 would indicate the difference in the opposite than the tested direction. Significance was defined as <5% overlap, i.e., probability values >0.95 or <0.05 (Lawlor et al., 2020).

### Results

#### 1.4 Participant characteristics

Participants with chronic pain indicated their *most painful* body part; these were in the lower limb (28% of participants), lower back (18%), upper back (15%), head (15%), upper limb (14%), abdomen (4%), chest (2%), or other (3%), although 89% reported usually experiencing pain in more than one location. In addition, they reported their *most frequent* usual pain location; these were in the lower back (55%), right and left upper leg (44% each), neck (43%), right shoulder (42%), and right lower leg (40%). We quantified patients’ number of painful locations according to the Widespread Pain Index (WPI; (Wolfe et al., 2010)). Thirty-six percent of participants with chronic pain scored ≥7 on WPI, indicating widespread pain.

While 11% of participants with chronic pain had not received a formal diagnosis, the most common reported diagnoses included fibromyalgia (22%), chronic back pain (20%), migraine (15%), complex regional pain syndrome (14%), osteoarthritis (12%), irritable bowel syndrome (12%), and hypermobility syndrome (10%).

Most often reported treatments and medications *regularly used* to relieve pain symptoms were paracetamol (45%), physical therapies (33%), NSAIDs (29%), antidepressants (28%), psychological therapies (24%), strong opioids (17%), antiepileptics (17%), complementary or alternative therapies (15%), and mild opioids (12%). Some patients also reported using topical analgesics (6%), electrotherapy (6%), steroids (4%), and other treatments or medications (5%). Five percent of patients did not use any regular treatments or medications for their pain. Most common ‘*as needed’* pain treatments or medications included paracetamol (43%), NSAIDs (36%), mild opioids (29%), strong opioids (16%), physical therapies (16%), and topical analgesics (11%). Some patients also reported using antidepressants (8%), psychological therapies (7%), electrotherapies (7%), complementary or alternative therapies (6%), steroids (3%), antiepileptics (1%), or other (4%). Four percent of patients did not use any *‘as needed’* treatments or medications.

Distribution, linearity and correlation between continuous baseline characteristic variables are in Figure S 1 and Figure S 2.

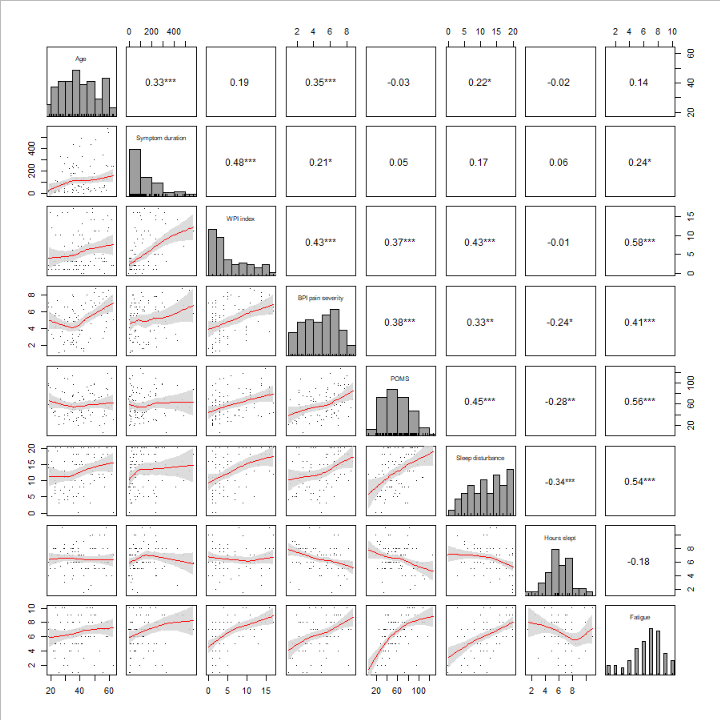
Figure S 1: Distribution, linearity and correlation between continuous characteristic variables at baseline in the chronic pain group.

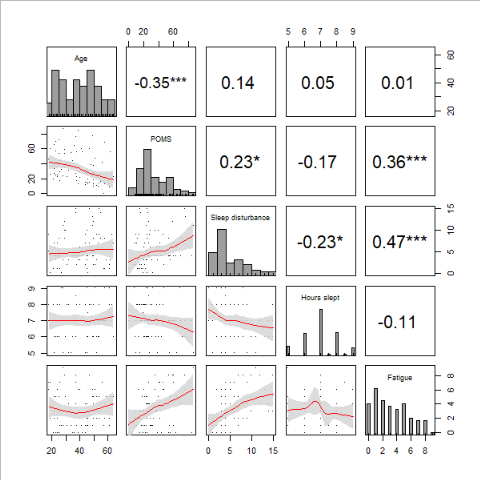
Figure S 2: Distribution, linearity and correlation between continuous characteristic variables at baseline in the healthy control group.

#### 1.5 Validity and reliability

##### 1.5.1 **Association of participants’ performance on the sustained attention and low cognitive load working memory tasks (H1a)**

In contrast to the hypothesis that participants' performance on the sustained attention and low cognitive load working memory tasks would be strongly associated, most performance metrics had poor intraclass correlation, except for RTs between the VV 0-back and VS 1-back tasks (Table S 2). These results suggest that 0-back and 1-back tasks measure distinct constructs and working memory does not substantially rely on sustained attention.

Table S 2. Intraclass correlation coefficients (ICC) with 95% confidence intervals (CI) between 0-back and 1-back tasks performance metrics in session 1, across all participants.

|  |  | ICC [95% CI] |
| --- | --- | --- |
| Accuracy | VV0 & AU1 | 0.05 [-0.07 to 0.17] |
|  | VV0 & VS1 | 0.13 [0.01 to 0.25] |
| RT | VV0 & AU1 | 0.38 [0.28 to 0.48] |
|  | VV0 & VS1 | 0.58 [0.49 to 0.65] |
| RT variability | VV0 & AU1 | 0.24 [0.12 to 0.35] |
|  | VV0 & VS1 | 0.27 [0.15 to 0.38] |
| d' | VV0 & AU1 | 0.18 [0.06 to 0.29] |
|  | VV0 & VS1 | 0.31 [0.19 to 0.41] |

##### 1.5.2 **Time effects on performance in low cognitive load tasks (H1b)**

The hypothesis that performance on low cognitive load tasks would deteriorate throughout the session was not supported, as paired Wilcoxon tests (the data were not normally distributed) showed no significant differences between the first and last completed 0/1-back task (regardless of task type) on any of the performance metrics in session 1 (ps 0.068 – 0.545; N = 158 [77 chronic pain, 81 controls]). The same results were found in session 2 (ps 0.074 - 0.676; N = 135 [68 chronic pain, 67 controls]). Note that only participants with available data on all low cognitive load tasks were included in these analyses, and participants were pooled across groups. Figure S 3 and Figure S 4 demonstrate that both groups exhibited similar results patterns.

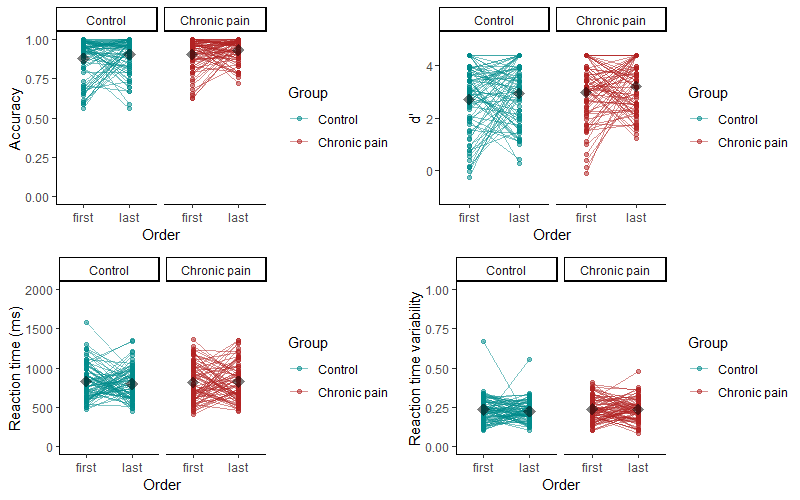
Figure S 3: Session 1 performance on first vs. last administered task (regardless of type of task), by group.

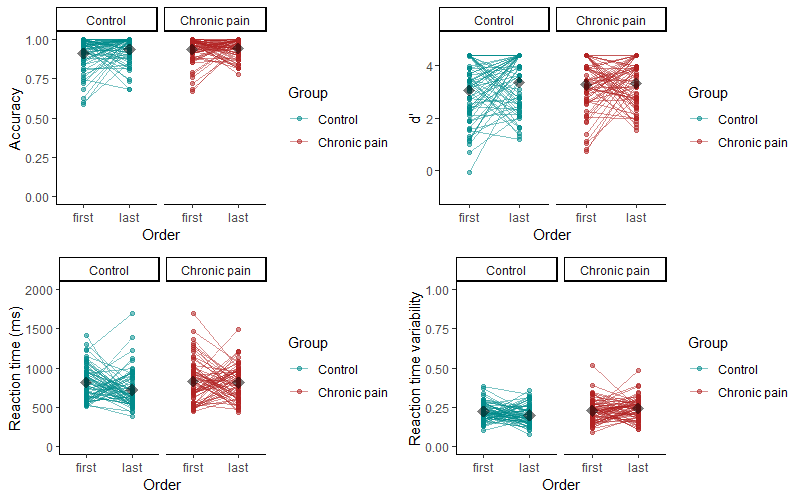
Figure S 4. Session 2 performance on first vs. last administered task (regardless of type of task), by group.

##### 1.5.3 Test-retest reliability between session 1 and session 2 (H1d)

Participants' performance on the same tasks were tested for agreement across the baseline and retest sessions (test-retest reliability). The results (Table S 3, also shown in Figure S 5 and Figure S 6) indicated moderate test-retest reliability for most tasks, both for accuracy and RT measures, except for very poor reliability for VV 0-back task accuracy in the chronic pain group (likely due to largely ceiling performance with a few outliers who had perfect performance in one session but poorer in the other session), poor reliability for VS 1-back task accuracy in the control group, and marginally poor/moderate reliability for the chronic pain group, and good reliability for the AU 1-back task RTs in the chronic pain group.

We explored whether the cases of poor retest reliability could be associated with transient mood, fatigue, sleep, or pain intensity. Indeed, the difference between session 1 and 2 in VS 1-back accuracy was correlated with the difference in the numbers of hours slept the night before the session in the control group (r = 0.29, p = 0.015), but no other significant correlations were found.

Table S 3. Test-retest reliability (ICC, 95% CI) for accuracy, d-prime (d’) and reaction time (RT)

| **Task** | **Chronic pain** | | | | **Control** | | | |
| --- | --- | --- | --- | --- | --- | --- | --- | --- |
|  | **n** | **Accuracy** | **d'** | **RT** | **n** | **Accuracy** | **d'** | **RT** |
| VV0 | 97 | 0.00 (-0.16 to 0.16) | 0.04 (-0.12 to 0.19) | 0.60 (0.48 to 0.70) | 85 | 0.70 (0.60 to 0.78) | 0.63 (0.50 to 0.73) | 0.71 (0.61 to 0.79) |
| VV2 | 91 | 0.61 (0.49 to 0.71) | 0.61 (0.49 to 0.71) | 0.67 (0.56 to 0.75) | 85 | 0.67 (0.56 to 0.76) | 0.77 (0.67 to 0.83) | 0.67 (0.57 to 0.76) |
| VS1 | 92 | 0.49 (0.34 to 0.62) | 0.45 (0.30 to 0.57) | 0.61 (0.47 to 0.72) | 85 | 0.36 (0.20 to 0.50) | 0.40 (0.24 to 0.54) | 0.58 (0.45 to 0.69) |
| VS2 | 89 | 0.61 (0.47 to 0.71) | 0.64 (0.52 to 0.74) | 0.61 (0.48 to 0.71) | 83 | 0.66 (0.53 to 0.75) | 0.64 (0.51 to 0.74) | 0.64 (0.45 to 0.76) |
| AU1 | 85 | 0.52 (0.37 to 0.64) | 0.52 (0.36 to 0.64) | 0.80 (0.73 to 0.86) | 84 | 0.53 (0.38 to 0.65) | 0.52 (0.38 to 0.64) | 0.65 (0.52 to 0.75) |
| AU2 | 81 | 0.55 (0.38 to 0.67) | 0.50 (0.33 to 0.64) | 0.67 (0.56 to 0.76) | 81 | 0.70 (0.58 to 0.79) | 0.65 (0.51 to 0.75) | 0.63 (0.50 to 0.73) |

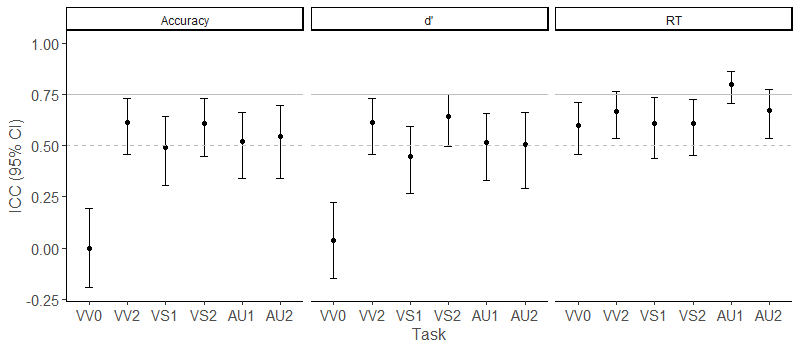

Figure S 5: Test-retest reliability in patients for accuracy, d-prime (d’) and reaction time (RT)

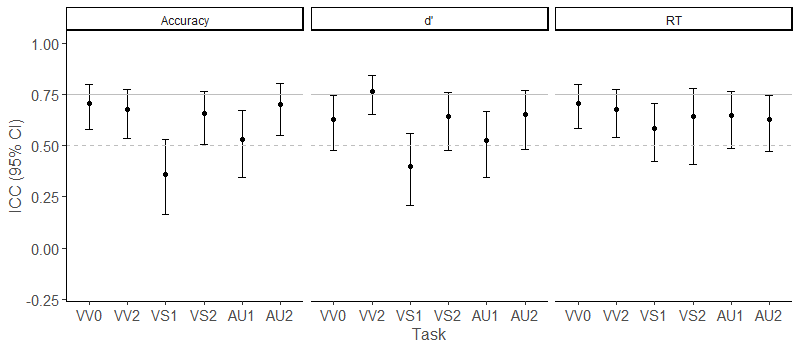

Figure S 6: Test-retest reliability in pain-free controls for accuracy, d-prime (d’) and reaction time (RT)

#### 1.6 Cognitive load and group effects: Session 1 primary analyses adjusted for potential confounders

##### 1.6.1 Accuracy and reaction times adjusted for covariates

Mixed effects models on accuracy and reaction time (Table S 4) outcomes, adjusted for potential confounders mood disturbance, sleep disturbance, and baseline fatigue, found consistent main effects of cognitive load across all tasks, where regardless of group, participants were less accurate and slower in higher (2-back) compared to lower (0/1-back) cognitive load conditions.

A significant effect of Group was observed in the VS task, where participants with chronic pain were overall less accurate than controls, but there was no significant interaction between group and load in this task. Reaction times were no different between groups in the VS task.

In AU task, chronic pain participants showed overall greater accuracy, which must be interpreted in the presence of an interaction on this task (Figure S 7C); this effect appears to be driven by the tendency of participants with chronic pain to be more accurate than controls in 1-back condition (p = 0.098), but they were less accurate in 2-back condition (p = 0.099). These differences, however, were not statistically significant. A significant interaction was also found for reaction times (Figure S 7F), with a trend towards slower reaction times among participants with chronic pain compared to controls in the 2-back, but not in 0/1-back tasks. However, the between-group contrast at higher load was not statistically significant. A similar significant interaction was also found for reaction times in the VV (Figure S 7D) task in the absence of a significant between-group simple effect at higher load (ps ≥ 0.144).

Table S 4. Results of generalised linear mixed models for accuracy and reaction times in each task in session 1, **adjusted** for mood disturbance, sleep disturbance, and fatigue.

|  | **Accuracy** | | | | | | | | | | | |
| --- | --- | --- | --- | --- | --- | --- | --- | --- | --- | --- | --- | --- |
|  | **VV** | | | | **VS** | | | | | **AU** | | |
| *Predictors* | *Log-Odds* | *95% CI* | | *p* | *Log-Odds* | *95% CI* | | *p* | | *Log-Odds* | *95% CI* | *p* |
| (Intercept) | 3.467 | 3.113, 3.822 | | **<0.001** | 1.984 | 1.676, 2.292 | | **<0.001** | | 1.424 | 1.203, 1.645 | **<0.001** |
| Load [2-back] | -2.262 | -2.435, -2.090 | | **<0.001** | -1.115 | -1.226, -1.005 | | **<0.001** | | -0.864 | -0.955, -0.773 | **<0.001** |
| Group [chronic pain] | -0.222 | -0.621, 0.176 | | 0.274 | -0.376 | -0.702, -0.049 | | **0.024** | | 0.25 | 0.001, 0.498 | **0.049** |
| Mood disturbance | -0.345 | -0.513, -0.178 | | **<0.001** | -0.307 | -0.464, -0.150 | | **<0.001** | | -0.138 | -0.250, -0.027 | **0.015** |
| Sleep disturbance | 0.018 | -0.015, 0.051 | | 0.297 | 0.032 | 0.003, 0.062 | | **0.033** | | 0.003 | -0.020, 0.025 | 0.801 |
| Fatigue | 0.083 | 0.015, 0.151 | | **0.017** | 0.072 | 0.011, 0.134 | | **0.022** | | 0.036 | -0.011, 0.082 | 0.137 |
| Load [2-back] * Group [chronic pain] | 0.132 | -0.105, 0.370 | | 0.274 | -0.025 | -0.181, 0.130 | | 0.75 | | -0.454 | -0.589, -0.320 | **<0.001** |
| σ^2^ | 3.29 | | | | 3.29 | | | | | 3.29 | | |
| τ_00_ | 0.65 | | | | 0.53 | | | | | 0.26 | | |
| N participants / observations | 177 / 25725 | | | | 168 / 24225 | | | | | 158 / 22350 | | |
| Marginal R^2^ / Conditional R^2^ | 0.245 / 0.370 | | | | 0.095 / 0.221 | | | | | 0.082 / 0.150 | | |
|  | **Reaction times** | | | | | | | | | | | |
|  | **VV** | | | | **VS** | | | | | **AU** | | |
| *Predictors* | *Estim.* | | *95% CI* | *p* | *Estim.* | *95% CI* | *p* | | *Estim.* | | *95% CI* | *p* |
| (Intercept) | 6.306 | | 6.235, 6.378 | **<0.001** | 6.561 | 6.485, 6.637 | **<0.001** | | 6.852 | | 6.783, 6.922 | **<0.001** |
| Load [2-back] | 0.313 | | 0.302, 0.323 | **<0.001** | 0.171 | 0.159, 0.183 | **<0.001** | | 0.113 | | 0.100, 0.126 | **<0.001** |
| Group [chronic pain] | <0.001 | | -0.074, 0.075 | 0.999 | 0.037 | -0.041, 0.115 | 0.348 | | <0.001 | | -0.076, 0.075 | 0.99 |
| Mood disturbance | 0.001 | | -0.036, 0.037 | 0.969 | -0.029 | -0.068, 0.010 | 0.145 | | -0.011 | | -0.047, 0.025 | 0.544 |
| Sleep disturbance | 0.004 | | -0.003, 0.012 | 0.216 | 0.002 | -0.005, 0.010 | 0.515 | | 0.003 | | -0.004, 0.010 | 0.389 |
| Fatigue | 0.003 | | -0.011, 0.018 | 0.649 | 0.011 | -0.004, 0.027 | 0.161 | | 0.004 | | -0.011, 0.019 | 0.568 |
| Load [2-back] * Group [chronic pain] | 0.069 | | 0.055, 0.084 | **<0.001** | -0.01 | -0.027, 0.007 | 0.257 | | 0.05 | | 0.031, 0.068 | **<0.001** |
| σ^2^ | 0.07 | | | | 0.09 | | | | 0.08 | | | |
| τ_00_ | 0.04 | | | | 0.04 | | | | 0.03 | | | |
| N participants / observations | 177 / 22519 | | | | 168 / 19776 | | | | 158 / 16384 | | | |
| Marginal R^2^ / Conditional R^2^ | 0.228 / 0.482 | | | | 0.064 / 0.333 | | | | 0.047 / 0.308 | | | |

VV: visuo-verbal task; VS: visuo-spatial task; AU: auditory-temporal task. 95% CI, 95% confidence interval; σ^2^, residual variance (within-participant); τ_00_, random intercept variance (between-participants); marginal R^2^, variance explained by fixed effects; conditional R^2^, variance explained by fixed and random effects. Reference terms are 0/1-back load, and control group.
Statistically significant results are marked in bold font.

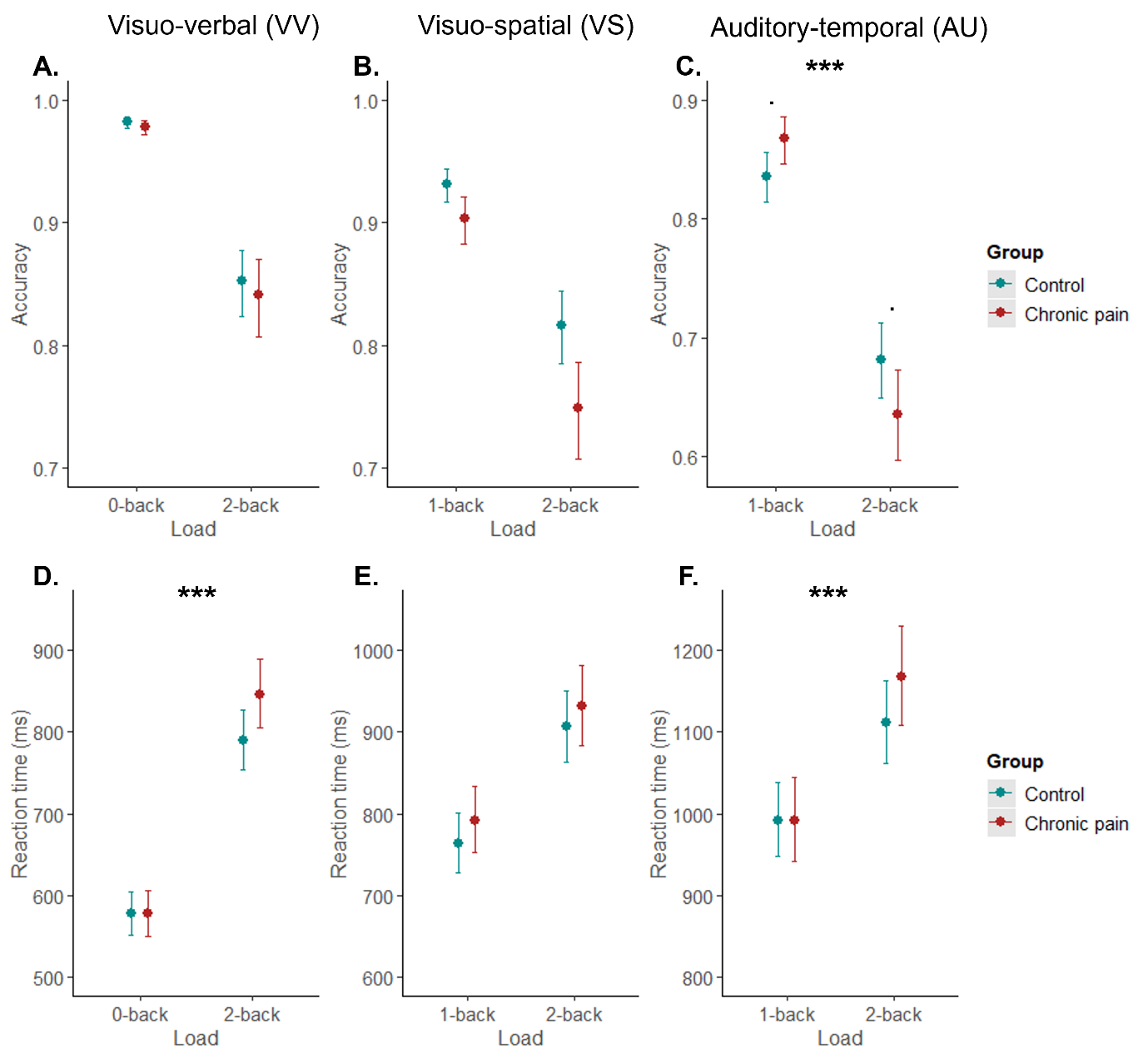
Figure S 7. Marginal effects (with 95% confidence intervals) of interaction terms between cognitive load (0/1-back, 2-back) and group (control, chronic pain) on accuracy (A-C) and reaction times (D-F) in each task in session 1, **adjusted** for mood disturbance, sleep disturbance, and fatigue. Reaction times have been back-transformed from log-scale for visualisation. Asterisks in bold indicate significant interactions. Asterisks in normal font indicate significant between-group contrasts (after Holm-Bonferroni adjustment). *** p<0.001, . p<0.1.

##### 1.6.2 Accuracy and reaction time differences adjusted for covariates

A linear mixed model on accuracy differences (Table S 5), adjusted for potential confounders mood disturbance, sleep disturbance, and baseline fatigue, showed a significant interaction between task type and group (Figure S 8A). Planned contrasts within each group revealed a greater decline in accuracy from lower to higher cognitive load in AU than VV task in participants with chronic pain (p < 0.001), but not in controls (p = 0.925). No differences were found between VS and VV outcomes in either group (ps ≥ 0.308). There were no interactions on reaction time difference, however, participants showed greater slowing of reaction times with increasing cognitive load in VV compared to VS and AU tasks, regardless of group (Table S 5).

Table S 5. Results of linear mixed models on accuracy and reaction time differences across tasks in session 1, **adjusted** for mood disturbance, sleep disturbance, and fatigue.

|  | **Accuracy difference** | | | **Reaction time difference** | | |
| --- | --- | --- | --- | --- | --- | --- |
| *Predictors* | *Estimates* | *95% CI* | *p* | *Estimates* | *95% CI* | *p* |
| (Intercept) | -0.188 | -0.222, -0.153 | **<0.001** | 171.174 | 105.774, 236.574 | **<0.001** |
| Task type [VS] | 0.026 | -0.005, 0.057 | 0.101 | -95.321 | -147.759, -42.882 | **<0.001** |
| Task type [AU] | 0.006 | -0.026, 0.037 | 0.733 | -123.925 | -177.500, -70.350 | **<0.001** |
| Group [chronic pain] | <0.001 | -0.040, 0.041 | 0.987 | 29.805 | -45.322, 104.932 | 0.437 |
| Mood disturbance | -0.027 | -0.042, -0.011 | **0.001** | -34.539 | -65.092, -3.986 | **0.027** |
| Sleep disturbance | 0.003 | 0.000, 0.006 | **0.032** | 5.156 | -0.872, 11.183 | 0.094 |
| Fatigue | <0.001 | -0.006, 0.006 | 0.986 | 11.395 | -1.002, 23.792 | 0.072 |
| Task type [VS] * Group [chronic pain] | -0.014 | -0.058, 0.031 | 0.539 | -59.806 | -135.171, 15.560 | 0.12 |
| Task type [AU] * Group [chronic pain] | -0.079 | -0.124, -0.033 | **0.001** | 0.122 | -77.158, 77.402 | 0.998 |
| σ^2^ | 0.01 | | | 28471.13 | | |
| τ_00_ | <0.01 | | | 12916.58 | | |
| N participants / observations | 170 / 455 | | | 170 / 455 | | |
| Marginal R^2^ / Conditional R^2^ | 0.105 / 0.276 | | | 0.128 / 0.400 | | |

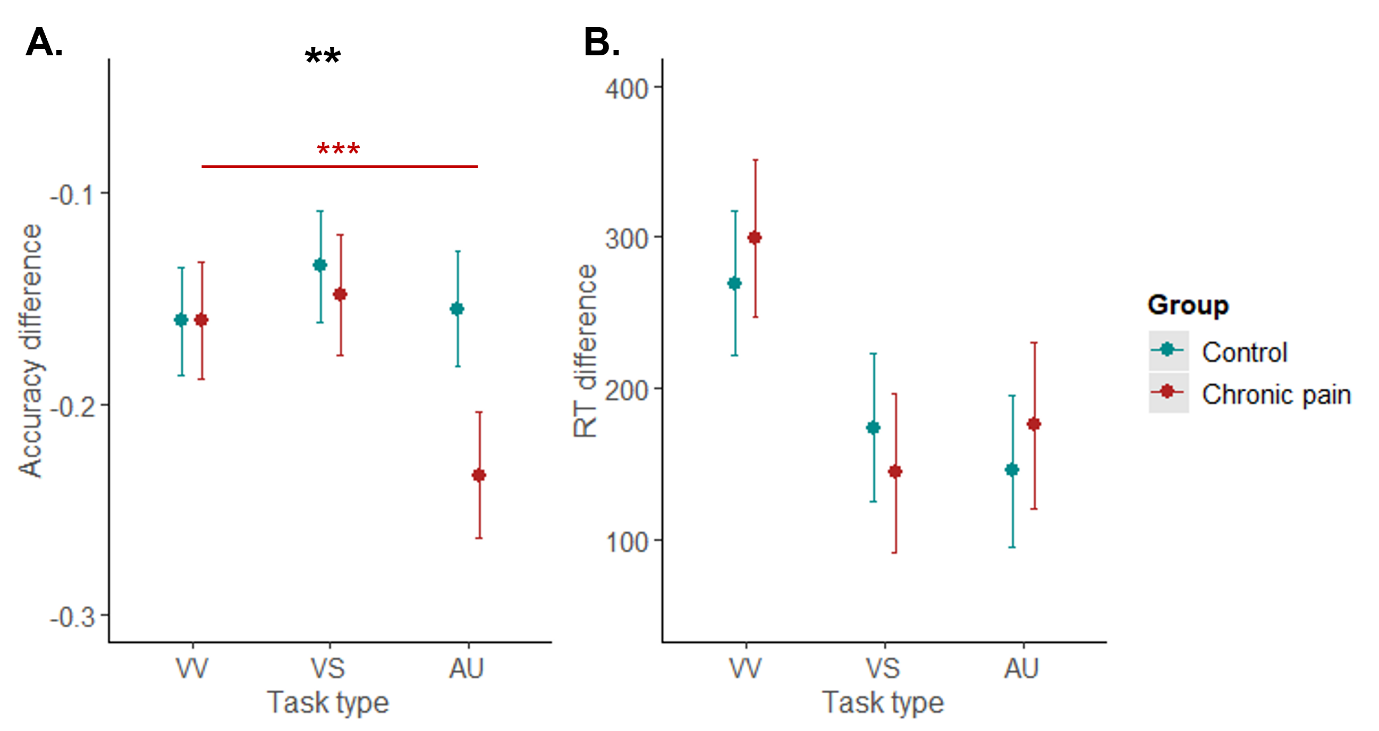
95% CI, confidence interval; σ^2^, residual variance (within- participant); τ_00_, random intercept variance (between- participants); marginal R^2^, variance explained by fixed effects; conditional R^2^, variance explained by fixed and random effects. Reference terms are VV (visuo-verbal) task type, and control group. VS, visuo-spatial; AU, auditory-temporal.
Statistically significant results are marked in bold font.

Figure S 8. Marginal effects (with 95% confidence intervals) of interaction terms between task type (VV, visuo-verbal; VS, visuo-spatial; AU, auditory-temporal) and group (control, chronic pain) on accuracy difference (A) and reaction time difference scores (B) in session 1, adjusted for mood disturbance, sleep disturbance, and fatigue. Asterisks in bold indicate significant interaction. Asterisks in normal font indicate significant between-group contrasts (after Holm-Bonferroni adjustment). *** p<0.001, ** p<0.01.

#### 1.7 Cognitive load and group effects: Session 2 primary analyses

##### 1.7.1 Accuracy and reaction times

As shown in Figure S 9, the results in session 2 demonstrated similar patterns to those observed in session 1 (Figure 3). A consistent effect of cognitive load was found across both outcomes (Table S 6 and Table S 7). There was no longer a significant interaction on accuracy in the VV task (the contrasts following this interaction in session 1 also found no significant between-group differences). There was a consistent effect of group on accuracy in AU task, as well as an interaction suggesting group differences in the same direction as in session 1, although only the advantage of chronic pain group in 1-back condition was marginally significant in session 2 (p = 0.066). Analysis of RTs found a consistent interaction in the VV task with chronic pain group being slower than controls in the 2-back condition (p = 0.002). There was also a consistent interaction in the AU task with chronic pain group being slower than controls in the 2-back condition (p = 0.001), and additionally in the 1-back condition (p = 0.039; this difference was not significant in session 1). The main effect of group was also significant in session 2. We found an additional interaction in the VS task (not present in session 1), with chronic pain group being marginally slower than controls in the 2-back condition (p = 0.080).

Table S 6. Results of generalised linear mixed models on accuracy in each task in session 2.

|  | **VV** | | | **VS** | | | **AU** | | |
| --- | --- | --- | --- | --- | --- | --- | --- | --- | --- |
| *Predictors* | *Log-Odds* | *95% CI* | *p* | *Log-Odds* | *95% CI* | *p* | *Log-Odds* | *95% CI* | *p* |
| (Intercept) | 4.377 | 4.074, 4.679 | **<0.001** | 2.795 | 2.560, 3.031 | **<0.001** | 1.933 | 1.777, 2.089 | **<0.001** |
| Load [2-back] | -2.359 | -2.564, -2.154 | **<0.001** | -1.171 | -1.298, -1.045 | **<0.001** | -1.017 | -1.119, -0.915 | **<0.001** |
| Group [chronic pain] | -0.111 | -0.532, 0.311 | 0.607 | 0.011 | -0.322, 0.344 | 0.947 | 0.243 | 0.020, 0.465 | **0.032** |
| Load [2-back] * Group [chronic pain] | 0.16 | -0.129, 0.450 | 0.278 | -0.042 | -0.221, 0.138 | 0.649 | -0.395 | -0.544, -0.246 | **<0.001** |
| σ^2^ | 3.29 | | | 3.29 | | | 3.29 | | |
| τ_00_ | 0.98 | | | 0.85 | | | 0.33 | | |
| N participants / observations | 152 / 22425 | | | 150 / 21825 | | | 145 / 20925 | | |
| Marginal R^2^ / Conditional R^2^ | 0.233 / 0.409 | | | 0.079 / 0.268 | | | 0.095 / 0.177 | | |

σ^2^, residual variance (within- participant); τ_00_, random intercept variance (between- participants); marginal R^2^, variance explained by fixed effects; conditional R^2^, variance explained by fixed and random effects. Reference terms are 0/1-back load, and control group.
Statistically significant results are marked in bold font.

Table S 7. Results of linear mixed models on reaction times in each task in session 2.

|  | **VV** | | | **VS** | | | **AU** | | |
| --- | --- | --- | --- | --- | --- | --- | --- | --- | --- |
| *Predictors* | *Estimates* | *95% CI* | *p* | *Estimates* | *95% CI* | *p* | *Estimates* | *95% CI* | *p* |
| (Intercept) | 6.332 | 6.289, 6.375 | **<0.001** | 6.594 | 6.551, 6.637 | **<0.001** | 6.863 | 6.823, 6.904 | **<0.001** |
| Load [2-back] | 0.287 | 0.276, 0.298 | **<0.001** | 0.125 | 0.114, 0.137 | **<0.001** | 0.093 | 0.081, 0.106 | **<0.001** |
| Group [chronic pain] | 0.032 | -0.028, 0.092 | 0.296 | 0.04 | -0.021, 0.100 | 0.201 | 0.061 | 0.004, 0.118 | **0.037** |
| Load [2-back] * Group [chronic pain] | 0.072 | 0.057, 0.088 | **<0.001** | 0.025 | 0.008, 0.042 | **0.004** | 0.045 | 0.028, 0.063 | **<0.001** |
| σ^2^ | 0.07 | | | 0.08 | | | 0.08 | | |
| τ_00_ | 0.03 | | | 0.03 | | | 0.03 | | |
| N participants / observations | 152 / 20024 | | | 150 / 18414 | | | 145 / 16057 | | |
| Marginal R^2^ / Conditional R^2^ | 0.203 / 0.456 | | | 0.044 / 0.327 | | | 0.044 / 0.313 | | |

σ^2^, residual variance (within- participants); τ_00_, random intercept variance (between- participants); marginal R^2^, variance explained by fixed effects; conditional R^2^, variance explained by fixed and random effects. Reference terms are 0/1-back load, and control group.
Statistically significant results are marked in bold font.

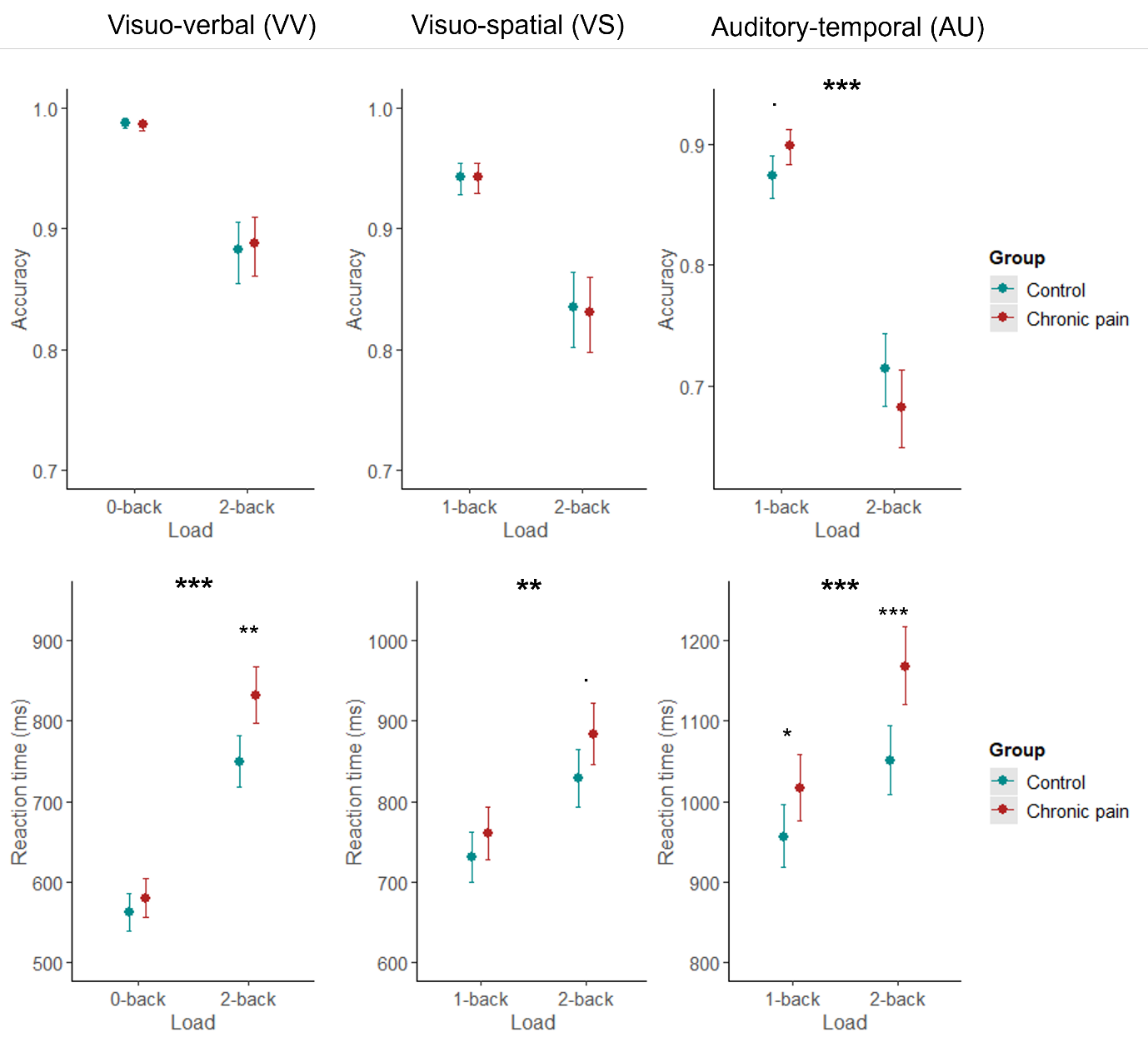
Figure S 9. Marginal effects (with 95% confidence intervals) of interaction terms between cognitive load (0/1-back, 2-back) and group (control, chronic pain) on accuracy (A-C) and reaction times (D-F) in each task in session 2. Reactions times have been back-transformed from log-scale for visualisation. Asterisks in bold indicate significant interactions. Asterisks in normal font indicate significant between-group contrasts (after Holm-Bonferroni adjustment). *** p<0.001, ** p<0.01, . p<0.1.

##### 1.7.2 Accuracy and reaction times adjusting for potential confounders

As shown in Figure S 10, most results patterns in session 2 were similar to those observed in session 1 (Figure S 7). A consistent effect of cognitive load was found across both outcomes (Table S 8 and Table S 9). The group effects on accuracy in VS and AU tasks found in session 1 were no longer significant. There was a consistent interaction in the AU task, although the contrasts found no significant between-group differences (which were marginally significant in session 1). Analyses of RTs found consistent interactions in VV and AU tasks, yet similar to session 1, without significant between-group contrasts. There was an additional interaction on the VS task, yet again without significant between-group contrasts.

Table S 8. Results of generalised linear mixed models for accuracy in each task in session 2, **adjusted** for mood disturbance, sleep disturbance, and fatigue.

|  | **VV** | | | **VS** | | | **AU** | | |
| --- | --- | --- | --- | --- | --- | --- | --- | --- | --- |
| *Predictors* | *Log-Odds* | *95% CI* | *p* | *Log-Odds* | *95% CI* | *p* | *Log-Odds* | *95% CI* | *p* |
| (Intercept) | 4.02 | 3.624, 4.416 | **<0.001** | 2.615 | 2.271, 2.960 | **<0.001** | 1.792 | 1.568, 2.016 | **<0.001** |
| Load [2-back] | -2.36 | -2.565, -2.155 | **<0.001** | -1.172 | -1.299, -1.045 | **<0.001** | -1.016 | -1.118, -0.914 | **<0.001** |
| Group [chronic pain] | -0.099 | -0.606, 0.408 | 0.702 | 0.045 | -0.380, 0.469 | 0.837 | 0.249 | -0.030, 0.528 | 0.081 |
| Mood disturbance | -0.364 | -0.558, -0.170 | **<0.001** | -0.198 | -0.378, -0.017 | **0.032** | -0.139 | -0.255, -0.022 | **0.02** |
| Sleep disturbance | -0.001 | -0.042, 0.039 | 0.943 | -0.01 | -0.049, 0.028 | 0.594 | 0.007 | -0.019, 0.034 | 0.585 |
| Fatigue | 0.086 | 0.000, 0.171 | **0.05** | 0.061 | -0.020, 0.142 | 0.137 | 0.017 | -0.037, 0.071 | 0.544 |
| Load [2-back] * Group [chronic pain] | 0.164 | -0.126, 0.454 | 0.268 | -0.041 | -0.220, 0.139 | 0.657 | -0.396 | -0.545, -0.247 | **<0.001** |
| σ^2^ | 3.29 | | | 3.29 | | | 3.29 | | |
| τ_00_ | 0.88 | | | 0.81 | | | 0.32 | | |
| N participants / observations | 152 / 22425 | | | 150 / 21825 | | | 145 / 20925 | | |
| Marginal R^2^ / Conditional R^2^ | 0.251 / 0.409 | | | 0.086 / 0.267 | | | 0.098 / 0.177 | | |

VV: visuo-verbal task; VS: visuo-spatial task; AU: auditory-temporal task. σ^2^, residual variance (within- participants); τ_00_, random intercept variance (between-participant); marginal R^2^, variance explained by fixed effects; conditional R^2^, variance explained by fixed and random effects. Reference terms are 0/1-back load, and control group.
Statistically significant results are marked in bold font.

Table S 9. Results of linear mixed models for reaction times in each task in session 2, **adjusted** for mood disturbance, sleep disturbance, and fatigue.

|  | **VV** | | | **VS** | | | **AU** | | |
| --- | --- | --- | --- | --- | --- | --- | --- | --- | --- |
| *Predictors* | *Estim.* | *95% CI* | *p* | *Estim.* | *95% CI* | *p* | *Estim.* | *95% CI* | *p* |
| (Intercept) | 6.244 | 6.180, 6.308 | **<0.001** | 6.503 | 6.439, 6.568 | **<0.001** | 6.801 | 6.739, 6.862 | **<0.001** |
| Load [2-back] | 0.287 | 0.276, 0.297 | **<0.001** | 0.125 | 0.114, 0.137 | **<0.001** | 0.094 | 0.081, 0.106 | **<0.001** |
| Group [chronic pain] | -0.054 | -0.132, 0.024 | 0.174 | -0.026 | -0.103, 0.052 | 0.521 | 0.028 | -0.046, 0.103 | 0.457 |
| Mood disturbance | -0.025 | -0.059, 0.010 | 0.164 | -0.041 | -0.076, -0.007 | **0.019** | -0.037 | -0.070, -0.003 | **0.031** |
| Sleep disturbance | 0.009 | 0.001, 0.016 | **0.021** | 0.009 | 0.002, 0.017 | **0.014** | 0.007 | -0.001, 0.014 | 0.069 |
| Fatigue | 0.013 | -0.002, 0.028 | 0.097 | 0.009 | -0.006, 0.025 | 0.236 | 0.004 | -0.011, 0.019 | 0.62 |
| Load [2-back] * Group [chronic pain] | 0.073 | 0.057, 0.088 | **<0.001** | 0.025 | 0.008, 0.042 | **0.004** | 0.045 | 0.028, 0.063 | **<0.001** |
| σ^2^ | 0.07 | | | 0.08 | | | 0.08 | | |
| τ_00_ | 0.03 | | | 0.03 | | | 0.03 | | |
| N participants / observations | 152 / 20024 | | | 150 / 18414 | | | 145 / 16057 | | |
| Marginal R^2^ / Conditional R^2^ | 0.223 / 0.459 | | | 0.068 / 0.330 | | | 0.058 / 0.316 | | |

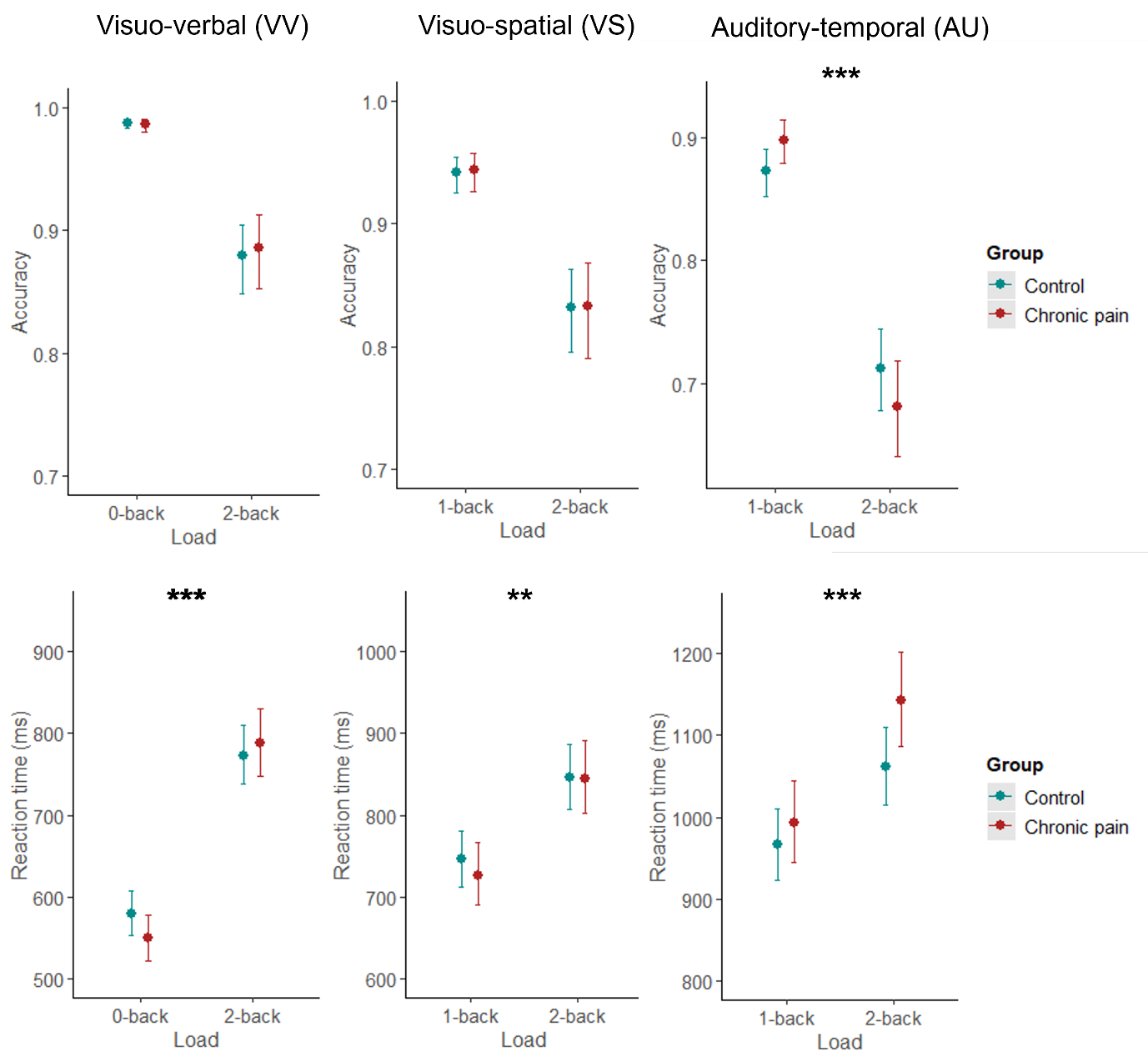
VV: visuo-verbal task; VS: visuo-spatial task; AU: auditory-temporal task. σ^2^, residual variance (within-participants); τ_00_, random intercept variance (between-participants); marginal R^2^, variance explained by fixed effects; conditional R^2^, variance explained by fixed and random effects. Reference terms are 0/1-back load, and control group.
Statistically significant results are marked in bold font.

Figure S 10. Marginal effects (with 95% confidence intervals) of interaction terms between cognitive load (0/1-back, 2-back) and group (control, chronic pain) on accuracy (A-C) and reaction times (D-F) in each task in session 2, **adjusted** for mood disturbance, sleep disturbance, and fatigue. Reaction times have been back-transformed from log-scale for visualisation. Asterisks in bold indicate significant interactions. Asterisks in normal font indicate significant between-group contrasts (after Holm-Bonferroni adjustment). *** p<0.001, ** p<0.01.

##### 1.7.3 Accuracy and reaction time differences

The results from session 2 (Table S 10, Figure S 11) presented consistent patterns to those found in session 1 (Table 3, Figure 4). There was a significant interaction on accuracy difference scores suggesting a greater decline in accuracy from lower to higher cognitive load in AU than VV task in participants with chronic pain (p < 0.001), but not in controls (p = 0.471). Additionally, a significant effect of group on RTs (only marginal in session 1) suggested that participants with chronic pain overall tended towards greater slowing of reaction times with increasing cognitive load than control participants.

Table S 10. Results of linear mixed models on accuracy and reaction time differences across tasks in session 2.

|  | **Accuracy difference** | | | **Reaction time difference** | | |
| --- | --- | --- | --- | --- | --- | --- |
| *Predictors* | *Estimates* | *95% CI* | *p* | *Estimates* | *95% CI* | *p* |
| (Intercept) | -0.139 | -0.163, -0.114 | **<0.001** | 226.676 | 182.620, 270.731 | **<0.001** |
| Task type [VS] | 0.019 | -0.011, 0.048 | 0.211 | -112.849 | -163.194, -62.503 | **<0.001** |
| Task type [AU] | -0.021 | -0.051, 0.008 | 0.156 | -121.584 | -172.761, -70.406 | **<0.001** |
| Group [chronic pain] | 0.016 | -0.018, 0.051 | 0.357 | 82.86 | 20.372, 145.348 | **0.009** |
| Task type [VS] * Group [chronic pain] | -0.019 | -0.060, 0.022 | 0.361 | -47.997 | -119.518, 23.524 | 0.188 |
| Task type [AU] * Group [chronic pain] | -0.073 | -0.115, -0.031 | **0.001** | -21.385 | -94.196, 51.426 | 0.565 |
| σ^2^ | 0.01 | | | 23654.43 | | |
| τ_00_ | <0.01 | | | 13900.89 | | |
| N participants / observations | 152 / 422 | | | 152 / 422 | | |
| Marginal R^2^ / Conditional R^2^ | 0.090 / 0.371 | | | 0.120 / 0.446 | | |

σ^2^, residual variance (within-participant); τ_00_, random intercept variance (between- participants); marginal R^2^, variance explained by fixed effects; conditional R^2^, variance explained by fixed and random effects. Reference terms are VV (visuo-verbal) task type, and control group. VS, visuo-spatial; AU, auditory-temporal.
Statistically significant results are marked in bold font.

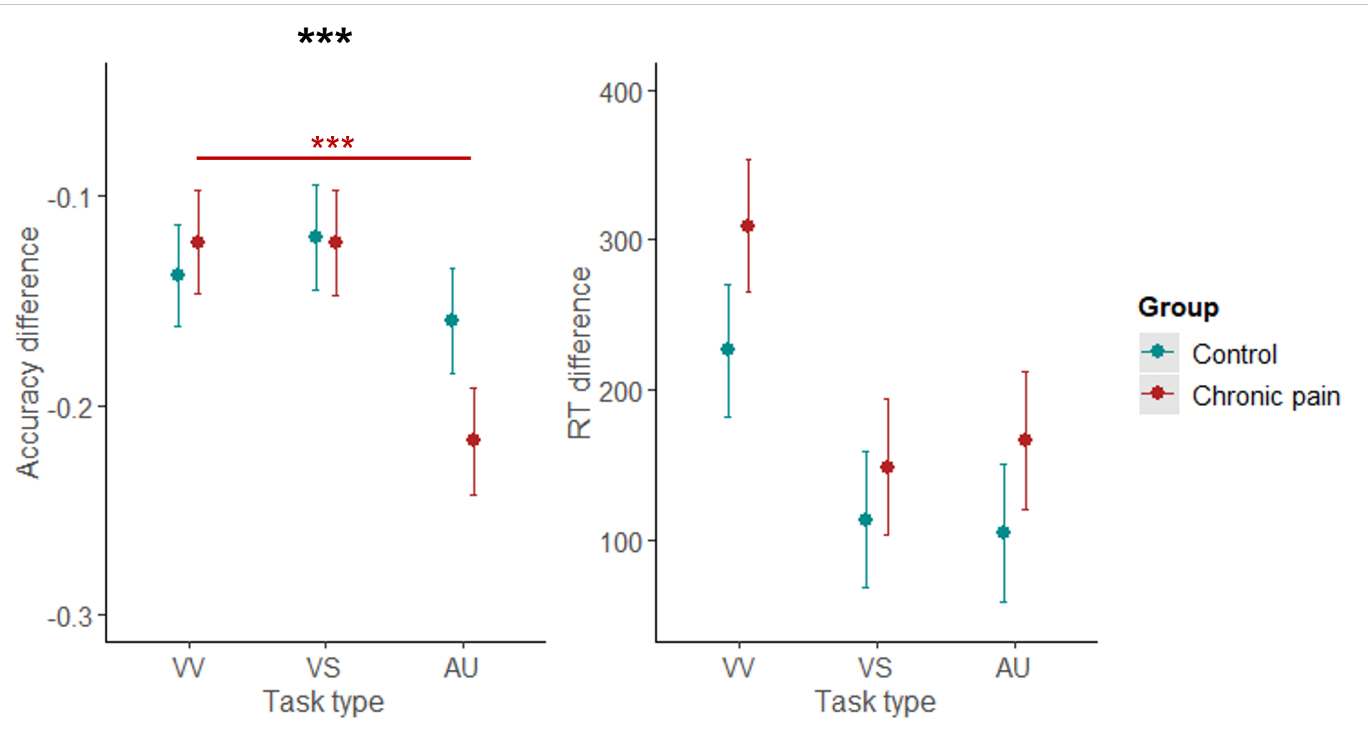
Figure S 11. Marginal effects (with 95% confidence intervals) of interaction terms between task type (VV, visuo-verbal; VS, visuo-spatial; AU, auditory-temporal) and group (control, chronic pain) on accuracy difference (A) and reaction time difference scores (B) in session 2. Asterisks in bold indicate significant interaction. Asterisks in normal font indicate significant between-group contrasts (after Holm-Bonferroni adjustment). *** p<0.001.

##### 1.7.4 Accuracy and reaction time differences adjusting for potential confounders

The results from session 2 were consistent with those from session 1 (Table S 11, Figure S 12), showing a significant interaction on accuracy difference scores where only participants with chronic pain (p < 0.001) had a greater decline in accuracy from lower to higher cognitive load in AU than VV task.

Table S 11. Results of linear mixed models on accuracy and reaction time differences across tasks in session 2, **adjusted** for mood disturbance, sleep disturbance, and fatigue.

|  | **Accuracy difference** | | | **Reaction time difference** | | |
| --- | --- | --- | --- | --- | --- | --- |
| *Predictors* | *Estimates* | *95% CI* | *p* | *Estimates* | *95% CI* | *p* |
| (Intercept) | -0.155 | -0.187, -0.123 | **<0.001** | 169.932 | 111.457, 228.407 | **<0.001** |
| Task type [VS] | 0.018 | -0.011, 0.047 | 0.212 | -113.084 | -163.378, -62.789 | **<0.001** |
| Task type [AU] | -0.022 | -0.051, 0.008 | 0.152 | -122.373 | -173.485, -71.261 | **<0.001** |
| Group [chronic pain] | 0.02 | -0.021, 0.061 | 0.346 | 58.196 | -16.199, 132.591 | 0.125 |
| Mood disturbance | -0.019 | -0.034, -0.004 | **0.013** | -40.158 | -68.070, -12.246 | **0.005** |
| Sleep disturbance | 0.001 | -0.002, 0.004 | 0.563 | 7.221 | 1.279, 13.164 | **0.017** |
| Fatigue | 0.001 | -0.005, 0.008 | 0.672 | 1.078 | -11.326, 13.482 | 0.865 |
| Task type [VS] * Group [chronic pain] | -0.019 | -0.060, 0.022 | 0.371 | -47.486 | -118.935, 23.962 | 0.193 |
| Task type [AU] * Group [chronic pain] | -0.073 | -0.115, -0.031 | **0.001** | -20.601 | -93.325, 52.123 | 0.579 |
| σ^2^ | 0.01 | | | 23616.96 | | |
| τ_00_ | <0.01 | | | 12626.24 | | |
| N participants / observations | 152 / 455 | | | 152 / 455 | | |
| Marginal R^2^ / Conditional R^2^ | 0.110 / 0.377 | | | 0.160 / 0.452 | | |

σ^2^, residual variance (within-participant); τ_00_, random intercept variance (between-participants); marginal R^2^, variance explained by fixed effects; conditional R^2^, variance explained by fixed and random effects. Reference terms are VV (visuo-verbal) task type, and control group. VS, visuo-spatial; AU, auditory-temporal.

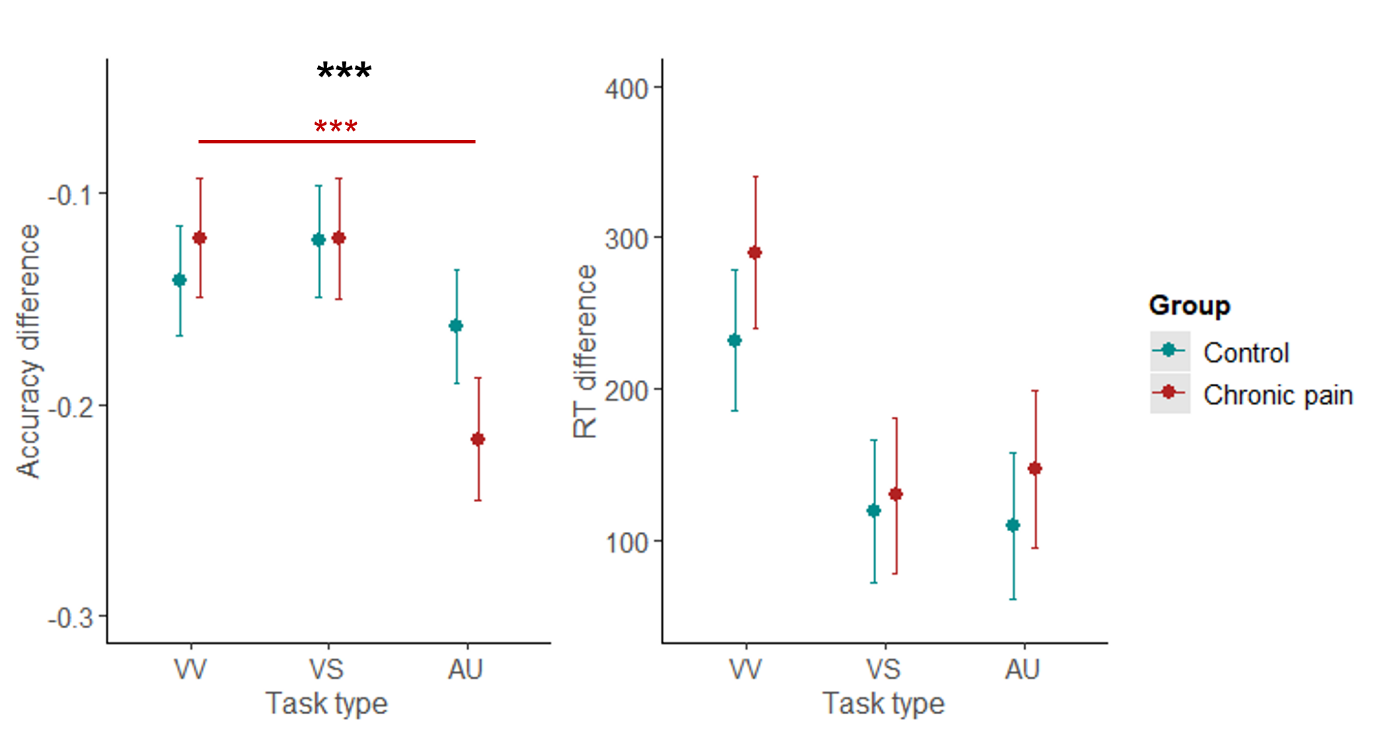
Statistically significant results are marked in bold font.

Figure S 12. Marginal effects (with 95% confidence intervals) of interaction terms between task type (VV, visuo-verbal; VS, visuo-spatial; AU, auditory-temporal) and group (control, chronic pain) on accuracy difference (A) and reaction time difference scores (B) in session 2, adjusted for mood disturbance, sleep disturbance, and fatigue. Asterisks in bold indicate significant interaction. Asterisks in normal font indicate significant between-group contrasts (after Holm-Bonferroni adjustment). *** p<0.001.

#### 1.8 Cognitive load and group effects: Session 1 secondary analyses

##### 1.8.1 Signal Detection Theory metrics (d’ and beta)

Mixed effects models on signal detection theory metrics d’ (Table S 12) and beta (Table S 13) found consistent main effects of cognitive load across all tasks, where regardless of group, participants showed better sensitivity and more conservative bias in lower (0/1-back) compared to higher (2-back) cognitive load conditions. There were no significant effects of group. An interaction between cognitive load and group was found for d’ in AU task (Figure S 13C), however, contrasts only revealed significantly better sensitivity in 1-back compared to 2-back condition within each group (ps < 0.001), but no significant between-group differences within each load condition (ps ≥ 0.142). There were no other significant interactions (Figure S 13).

Table S 12. Linear mixed models results for d’ in each task in session 1.

|  | **VV** | | | **VS** | | | **AU** | | |
| --- | --- | --- | --- | --- | --- | --- | --- | --- | --- |
| *Predictors* | *Estim.* | *95% CI* | *p* | *Estim.* | *95% CI* | *p* | *Estim.* | *95% CI* | *p* |
| (Intercept) | 3.812 | 3.628, 3.997 | **<0.001** | 2.995 | 2.786, 3.204 | **<0.001** | 2.058 | 1.874, 2.242 | **<0.001** |
| L  oad [2-back] | -1.819 | -2.042, -1.597 | **<0.001** | -1.369 | -1.576, -1.163 | **<0.001** | -1.217 | -1.409, -1.026 | **<0.001** |
| Group [chronic pain] | -0.095 | -0.349, 0.159 | 0.463 | -0.177 | -0.468, 0.114 | 0.233 | 0.242 | -0.020, 0.504 | 0.07 |
| Load [2-back] * Group [chronic pain] | 0.098 | -0.214, 0.410 | 0.537 | 0.071 | -0.220, 0.362 | 0.634 | -0.441 | -0.716, -0.167 | **0.002** |
| σ^2^ | 0.55 | | | 0.44 | | | 0.37 | | |
| τ_00_ | 0.21 | | | 0.49 | | | 0.35 | | |
| N participants / observations | 183 / 351 | | | 173 / 330 | | | 163 / 308 | | |
| Marginal R^2^ / Conditional R^2^ | 0.508 / 0.645 | | | 0.324 / 0.681 | | | 0.424 / 0.706 | | |

σ^2^, residual variance (within-participant); τ_00_, random intercept variance (between-participants); marginal R^2^, variance explained by fixed effects; conditional R^2^, variance explained by fixed and random effects. Reference terms are 0/1-back load, and control group. VV, visuo-verbal; VS, visuo-spatial; AU, auditory-temporal.
Statistically significant results are marked in bold font.

Table S 13. Linear mixed models results for beta in each task in session 1.

|  | **VV** | | | **VS** | | | **AU** | | |
| --- | --- | --- | --- | --- | --- | --- | --- | --- | --- |
| *Predictors* | *Estim.* | *95% CI* | *p* | *Estim.* | *95% CI* | *p* | *Estim.* | *95% CI* | *p* |
| (Intercept) | 0.927 | 0.780, 1.074 | **<0.001** | 0.753 | 0.585, 0.920 | **<0.001** | 0.319 | 0.172, 0.466 | **<0.001** |
| Load [2-back] | -0.328 | -0.537, -0.119 | **0.002** | -0.465 | -0.666, -0.265 | **<0.001** | -0.255 | -0.435, -0.074 | **0.006** |
| Group [chronic pain] | -0.095 | -0.298, 0.107 | 0.355 | -0.009 | -0.243, 0.225 | 0.94 | 0.103 | -0.106, 0.312 | 0.334 |
| Load [2-back] * Group [chronic pain] | -0.023 | -0.314, 0.268 | 0.877 | 0.067 | -0.215, 0.350 | 0.639 | -0.107 | -0.365, 0.151 | 0.416 |
| σ^2^ | 0.48 | | | 0.42 | | | 0.33 | | |
| τ_00_ | <0.01 | | | 0.18 | | | 0.13 | | |
| N participants / observations | 183 / 351 | | | 173 / 330 | | | 163 / 308 | | |
| Marginal R^2^ / Conditional R^2^ | 0.061 / NA | | | 0.073 / 0.350 | | | 0.052 / 0.317 | | |

σ^2^, residual variance (within-participant); τ_00_, random intercept variance (between-participants); marginal R^2^, variance explained by fixed effects; conditional R^2^, variance explained by fixed and random effects. Reference terms are 0/1-back load, and control group. VV, visuo-verbal; VS, visuo-spatial; AU, auditory-temporal.
Statistically significant results are marked in bold font.

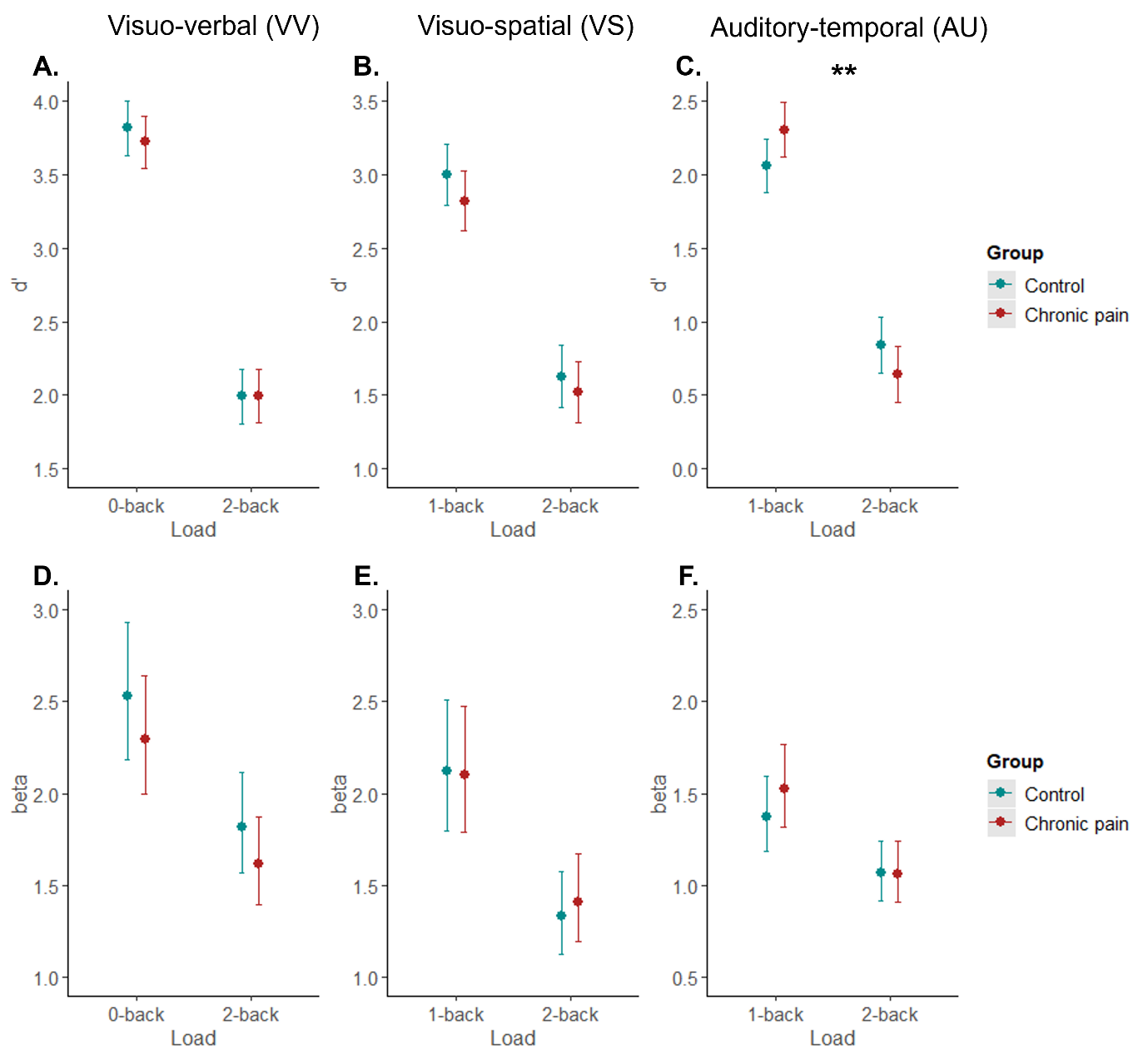
Figure S 13. Marginal effects (with 95% confidence intervals) of interaction terms between cognitive load (0/1-back, 2-back) and group (control, chronic pain) on d’ (A-C) and beta (D-F) in each task in session 1. Betas have been back-transformed from log-scale for visualisation. Asterisks in bold indicate significant interactions. ** p<0.01.

##### 1.8.2 Signal Detection Theory metrics (d’ and beta) adjusted for potential confounders

Mixed effects models on signal detection theory metrics d’ (Table S 14) and beta (Table S 15), adjusted for potential confounders mood disturbance, sleep disturbance, and baseline fatigue, found consistent main effects of cognitive load across all tasks, where regardless of group, participants showed better sensitivity and more conservative bias in lower (0/1-back) compared to higher (2-back) cognitive load conditions. There were no significant effects of group. An interaction between cognitive load and group was found for d’ in AU task (Figure S 14C), however, contrasts only revealed significantly better sensitivity in 1-back compared to 2-back condition within each group (ps < 0.001), but no significant between-group differences within each load condition (ps ≥ 0.211). There were no other significant interactions (Figure S 14).

Relative to the analysis without adjustments for potential confounders, the current models show that performance patterns remain largely consistent after accounting for participants’ mood and sleep disturbances and fatigue.

Table S 14. Results of generalised linear mixed models for d’ in each task in session 1, **adjusted** for mood disturbance, sleep disturbance, and fatigue.

|  | **VV** | | | **VS** | | | **AU** | | |
| --- | --- | --- | --- | --- | --- | --- | --- | --- | --- |
| *Predictors* | *Estim.* | *95% CI* | *p* | *Estim.* | *95% CI* | *p* | *Estim.* | *95% CI* | *p* |
| (Intercept) | 3.38 | 3.111, 3.648 | **<0.001** | 2.493 | 2.160, 2.825 | **<0.001** | 1.756 | 1.455, 2.056 | **<0.001** |
| Load [2-back] | -1.835 | -2.049, -1.621 | **<0.001** | -1.356 | -1.563, -1.148 | **<0.001** | -1.204 | -1.399, -1.008 | **<0.001** |
| Group [chronic pain] | -0.105 | -0.402, 0.192 | 0.489 | -0.295 | -0.652, 0.062 | 0.106 | 0.168 | -0.169, 0.505 | 0.329 |
| Mood disturbance | -0.287 | -0.413, -0.160 | **<0.001** | -0.307 | -0.473, -0.141 | **<0.001** | -0.162 | -0.310, -0.013 | **0.033** |
| Sleep disturbance | 0.015 | -0.010, 0.040 | 0.237 | 0.021 | -0.010, 0.053 | 0.18 | -0.001 | -0.031, 0.028 | 0.924 |
| Fatigue | 0.068 | 0.016, 0.119 | **0.01** | 0.072 | 0.007, 0.137 | **0.031** | 0.07 | 0.008, 0.132 | **0.027** |
| Load [2-back] * Group [chronic pain] | 0.041 | -0.262, 0.343 | 0.792 | 0.057 | -0.239, 0.353 | 0.706 | -0.455 | -0.738, -0.173 | **0.002** |
| σ^2^ | 0.5 | | | 0.44 | | | 0.37 | | |
| τ_00_ | 0.17 | | | 0.42 | | | 0.33 | | |
| N participants / observations | 177 / 340 | | | 168 / 320 | | | 158 / 298 | | |
| Marginal R^2^ / Conditional R^2^ | 0.566 / 0.678 | | | 0.373 / 0.679 | | | 0.435 / 0.702 | | |

σ^2^, residual variance (within-participant); τ_00_, random intercept variance (between-participants); marginal R^2^, variance explained by fixed effects; conditional R^2^, variance explained by fixed and random effects. Reference terms are 0/1-back load, and control group. VV, visuo-verbal; VS, visuo-spatial; AU, auditory-temporal.
Statistically significant results are marked in bold font.

Table S 15. Results of linear mixed models for beta in each task in session 1, **adjusted** for mood disturbance, sleep disturbance, and fatigue.

|  | **VV** | | | **VS** | | | **AU** | | |
| --- | --- | --- | --- | --- | --- | --- | --- | --- | --- |
| *Predictors* | *Estim.* | *95% CI* | *p* | *Estim.* | *95% CI* | *p* | *Estim.* | *95% CI* | *p* |
| (Intercept) | 0.796 | 0.584, 1.009 | **<0.001** | 0.253 | 0.002, 0.504 | **0.048** | 0.379 | 0.146, 0.613 | **0.001** |
| Load [2-back] | -0.31 | -0.519, -0.101 | **0.004** | -0.464 | -0.661, -0.267 | **<0.001** | -0.252 | -0.436, -0.068 | **0.007** |
| Group [chronic pain] | -0.122 | -0.363, 0.120 | 0.324 | -0.165 | -0.440, 0.111 | 0.241 | 0.188 | -0.078, 0.455 | 0.166 |
| Mood disturbance | -0.052 | -0.147, 0.044 | 0.288 | -0.282 | -0.403, -0.161 | **<0.001** | -0.013 | -0.125, 0.100 | 0.828 |
| Sleep disturbance | 0.011 | -0.007, 0.030 | 0.232 | 0.044 | 0.021, 0.067 | **<0.001** | -0.001 | -0.024, 0.022 | 0.93 |
| Fatigue | 0.007 | -0.032, 0.045 | 0.732 | 0.034 | -0.014, 0.081 | 0.168 | -0.02 | -0.068, 0.027 | 0.404 |
| Load [2-back] * Group [chronic pain] | -0.094 | -0.388, 0.200 | 0.531 | 0.086 | -0.194, 0.366 | 0.548 | -0.102 | -0.367, 0.164 | 0.454 |
| σ^2^ | 0.48 | | | 0.4 | | | 0.34 | | |
| τ_00_ | <0.01 | | | 0.14 | | | 0.13 | | |
| N participants / observations | 177 / 340 | | | 168 / 320 | | | 158 / 298 | | |
| Marginal R^2^ / Conditional R^2^ | 0.074 / NA | | | 0.174 / 0.386 | | | 0.055 / 0.317 | | |

σ^2^, residual variance (within-participant); τ_00_, random intercept variance (between-participants); marginal R^2^, variance explained by fixed effects; conditional R^2^, variance explained by fixed and random effects. Reference terms are 0/1-back load, and control group. VV, visuo-verbal; VS, visuo-spatial; AU, auditory-temporal.
Statistically significant results are marked in bold font.

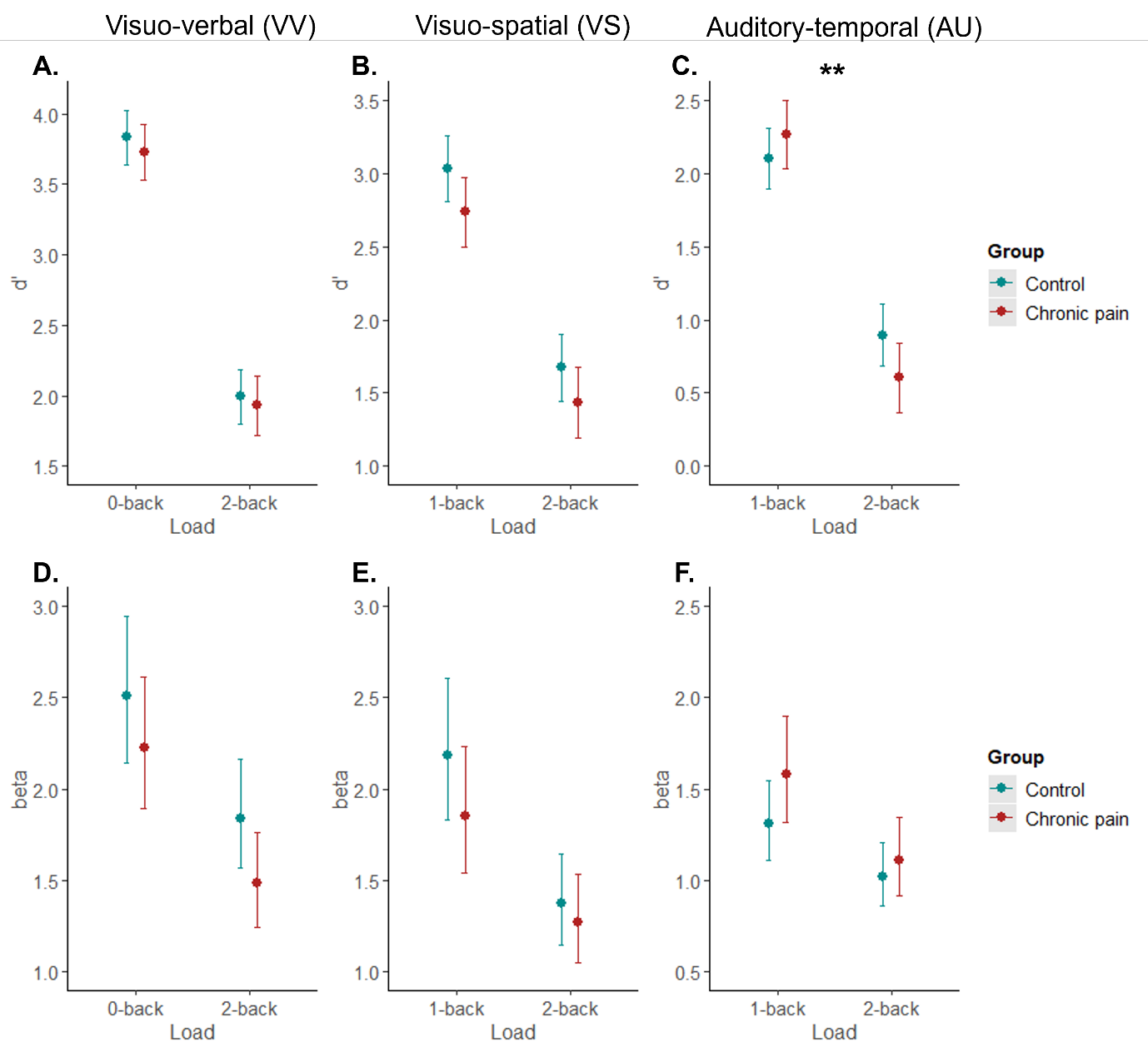
Figure S 14. Marginal effects (with 95% confidence intervals) of interaction terms between cognitive load (0/1-back, 2-back) and group (control, chronic pain) on d’ (A-C) and beta (D-F) in each task in session 1, **adjusted** for mood disturbance, sleep disturbance, and fatigue. Betas have been back-transformed from log-scale for visualisation. Asterisks in bold indicate significant interactions. ** p<0.01.

##### 1.8.3 Signal Detection Theory metrics (d’ and beta) – difference values between high and low cognitive load

Performance was quantified as a difference in d’ and beta between 0/1-back and 2-back conditions of each task. Therefore, lower d’ difference scores reflect greater decline in sensitivity with increasing cognitive load, and lower beta difference scores reflect greater reduction in (conservative) response bias with increasing cognitive load.

Linear mixed model on d’ difference (Table S 16) showed a significant interaction between task type and group (Figure S 15A). Planned contrasts within each group revealed less decline in sensitivity from lower to higher cognitive load in VS than VV task, both in participants with chronic pain (p = 0.007), and controls (p = 0.005). Controls also showed less decline in sensitivity in AU than VV task (p < 0.001), while participants with chronic pain did not (p = 0.799). Main effects of task type further suggested overall less decline in sensitivity in VS and AU tasks compared to VV task, regardless of group. There were no significant effects on beta difference (Table S 16).

Table S 16. Results of linear mixed models on d’ and beta differences across tasks in session 1.

|  | **d' difference** | | | **beta difference** | | |
| --- | --- | --- | --- | --- | --- | --- |
| *Predictors* | *Estimates* | *95% CI* | *p* | *Estimates* | *95% CI* | *p* |
| (Intercept) | -1.826 | -2.031, -1.620 | **<0.001** | -0.935 | -1.591, -0.279 | **0.005** |
| Task type [VS] | 0.459 | 0.175, 0.743 | **0.002** | -0.636 | -1.579, 0.306 | 0.186 |
| Task type [AU] | 0.608 | 0.319, 0.897 | **<0.001** | 0.053 | -0.906, 1.011 | 0.914 |
| Group [chronic pain] | 0.105 | -0.186, 0.396 | 0.478 | 0.135 | -0.793, 1.063 | 0.776 |
| Task type [VS] * Group [chronic pain] | -0.031 | -0.434, 0.371 | 0.879 | 0.365 | -0.970, 1.699 | 0.592 |
| Task type [AU] * Group [chronic pain] | -0.57 | -0.981, -0.158 | **0.007** | -0.519 | -1.883, 0.844 | 0.455 |
| σ^2^ | 0.85 | | | 9.41 | | |
| τ_00_ | 0.07 | | | <0.01 | | |
| N participants / observations | 176 / 470 | | | 176 / 470 | | |
| Marginal R^2^ / Conditional R^2^ | 0.056 / 0.132 | | | 0.007 / NA | | |

σ^2^, residual variance (within-participants); τ_00_, random intercept variance (between-participants); marginal R^2^, variance explained by fixed effects; conditional R^2^, variance explained by fixed and random effects. Reference terms are VV (visuo-verbal) task type, and control group. VS, visuo-spatial; AU, auditory-temporal.
Statistically significant results are marked in bold font.

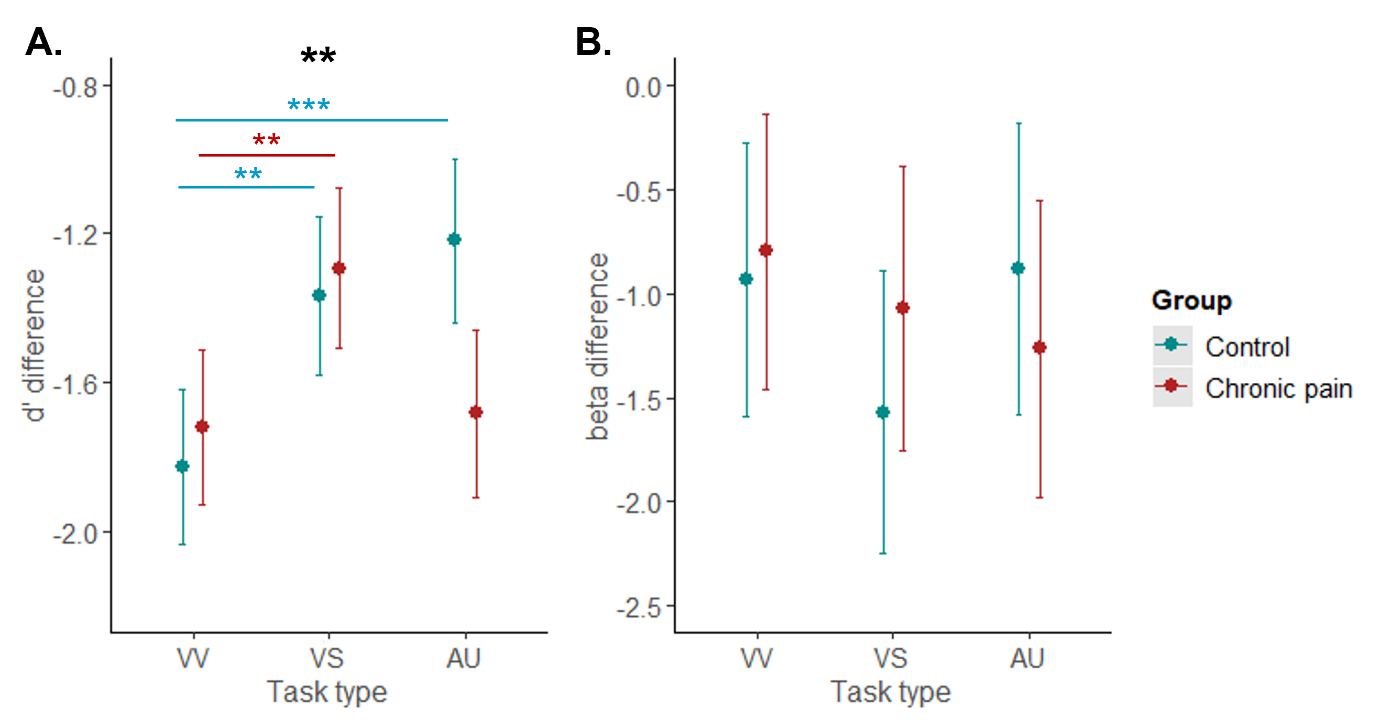
Figure S 15. Marginal effects (with 95% confidence intervals) of interaction terms between task type (VV, visuo-verbal; VS, visuo-spatial; AU, auditory-temporal) and group (control, chronic pain) on d’ difference (A) and beta difference scores (B) in session 1. Asterisks in bold indicate significant interaction. Asterisks in normal font indicate significant between-group contrasts (after Holm-Bonferroni adjustment). *** p<0.001, ** p<0.01.

##### 1.8.4 Signal Detection Theory metrics (d’ and beta) adjusted for potential confounders – difference values between high and low cognitive load

Linear mixed model on d’ difference (Table S 17), adjusted for potential confounders mood disturbance, sleep disturbance, and baseline fatigue, showed a significant interaction between task type and group (Figure S 16A). Planned contrasts within each group revealed less decline in sensitivity from lower to higher cognitive load in VS than VV task, both in participants with chronic pain (p = 0.002), and controls (p = 0.002). Controls also showed less decline in sensitivity in AU than VV task (p < 0.001), while participants with chronic pain did not (p = 0.524). Main effects of task type further suggested overall less decline in sensitivity in VS and AU tasks compared to VV task, regardless of group. There were no significant effects on beta difference (Table S 17).

Table S 17. Results of linear mixed models on d’ and beta differences across tasks in session 1, adjusting for potential confounders.

|  | **d' difference** | | | **beta difference** | | |
| --- | --- | --- | --- | --- | --- | --- |
| *Predictors* | *Estimates* | *95% CI* | *p* | *Estimates* | *95% CI* | *p* |
| (Intercept) | -1.939 | -2.220, -1.657 | **<0.001** | -0.622 | -1.497, 0.253 | 0.163 |
| Task type [VS] | 0.481 | 0.203, 0.759 | **0.001** | -0.786 | -1.722, 0.151 | 0.1 |
| Task type [AU] | 0.638 | 0.354, 0.921 | **<0.001** | -0.098 | -1.051, 0.855 | 0.84 |
| Group [chronic pain] | 0.09 | -0.246, 0.426 | 0.598 | -0.201 | -1.265, 0.863 | 0.711 |
| Mood disturbance | -0.127 | -0.250, -0.005 | **0.042** | 0.106 | -0.261, 0.474 | 0.571 |
| Sleep disturbance | 0.022 | -0.002, 0.047 | 0.071 | 0.014 | -0.059, 0.087 | 0.714 |
| Fatigue | -0.026 | -0.076, 0.024 | 0.304 | -0.059 | -0.209, 0.090 | 0.437 |
| Task type [VS] * Group [chronic pain] | 0.016 | -0.383, 0.414 | 0.938 | 0.73 | -0.609, 2.070 | 0.285 |
| Task type [AU] * Group [chronic pain] | -0.542 | -0.950, -0.135 | **0.009** | -0.231 | -1.600, 1.138 | 0.741 |
| σ^2^ | 0.81 | | | 9.17 | | |
| τ_00_ | 0.08 | | | <0.01 | | |
| N participants / observations | 170 / 455 | | | 170 / 455 | | |
| Marginal R^2^ / Conditional R^2^ | 0.085 / 0.167 | | | 0.009 / NA | | |

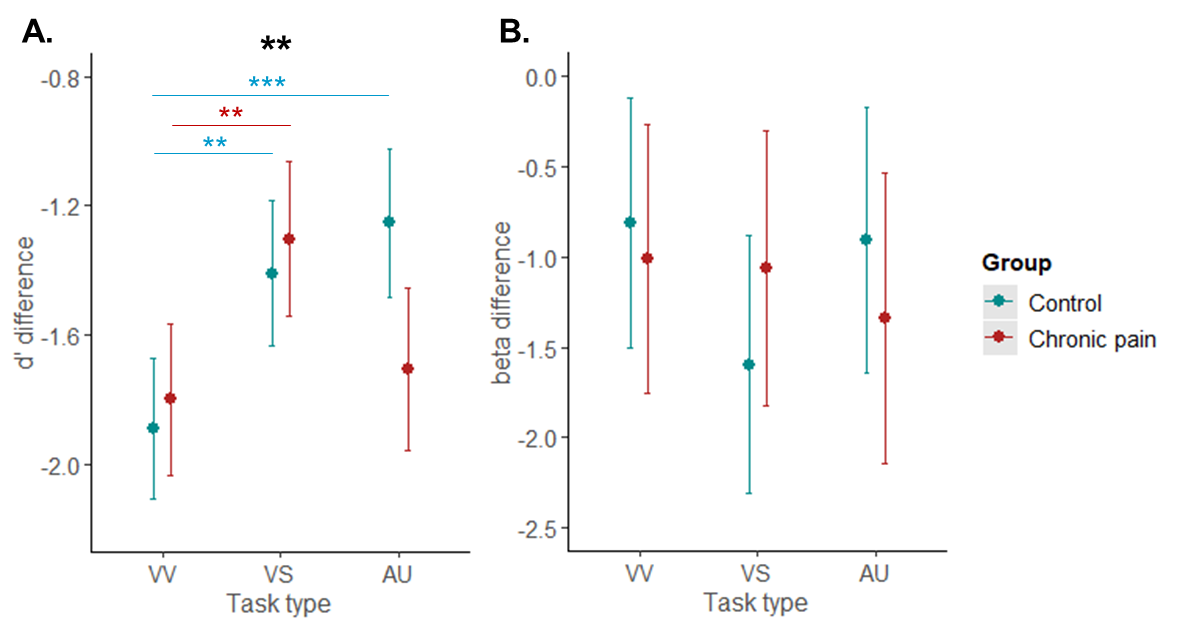
σ^2^, residual variance (within-participant); τ_00_, random intercept variance (between-participants); marginal R^2^, variance explained by fixed effects; conditional R^2^, variance explained by fixed and random effects. Reference terms are VV (visuo-verbal) task type, and control group. VS, visuo-spatial; AU, auditory-temporal.
Statistically significant results are marked in bold font.

Figure S 16. Marginal effects (with 95% confidence intervals) of interaction terms between task type (VV, visuo-verbal; VS, visuo-spatial; AU, auditory-temporal) and group (control, chronic pain) on d’ difference (A) and beta difference scores (B) in session 1, adjusted for mood disturbance, sleep disturbance, and fatigue. Asterisks in bold indicate significant interaction. Asterisks in normal font indicate significant between-group contrasts (after Holm-Bonferroni adjustment). *** p<0.001, ** p<0.01.

#### 1.9 Predictors of performance: Session 1 secondary analyses

##### 1.9.1 Predictors of accuracy and reaction time difference scores

Pain duration was not a significant independent predictor of change in accuracy or RTs (Table S 18) with increasing load in any of the tasks (see also Figure S 17). Only greater sleep disturbance predicted less decline in accuracy with increasing load in the visuo-verbal task (estimate = 0.007 on the accuracy-difference scale, 95% CI 0.001 to 0.014, equivalent to ~0.7 percentage points less accuracy decline per one-unit increase in sleep disturbance), and older age predicted greater decline in accuracy (visuo-verbal task – ~3 percentage points greater decline per decade) and RTs (visuo-verbal and auditory-temporal tasks – roughly ~58ms and ~55ms greater slowing per decade). Predictors of d' difference scores, as well as predictors of sustained attention performance (RT variability in 0-back visual task) are reported in the Supplement.

Table S 18 Predictors of accuracy difference scores and reaction time difference scores (2-back – 0/1-back) for each task type.

|  | **Accuracy difference** | | | | | | | | | |
| --- | --- | --- | --- | --- | --- | --- | --- | --- | --- | --- |
|  | **VV** | | | **VS** | | | **AU** | | | |
| *Predictors* | *Estim.* | *95% CI* | *p* | *Estim.* | *95% CI* | *p* | *Estim.* | *95% CI* | *p* | |
| (Intercept) | -0.076 | -0.208, 0.055 | 0.252 | -0.068 | -0.195, 0.059 | 0.287 | -0.133 | -0.260, -0.005 | **0.041** | |
| Pain duration (log) | 0.008 | -0.016, 0.032 | 0.523 | 0.003 | -0.021, 0.028 | 0.787 | 0 | -0.024, 0.024 | 0.986 | |
| Pain severity | -0.002 | -0.019, 0.014 | 0.775 | 0.003 | -0.016, 0.021 | 0.771 | -0.005 | -0.022, 0.011 | 0.522 | |
| Opioids use [yes] | -0.015 | -0.079, 0.048 | 0.634 | -0.016 | -0.083, 0.052 | 0.643 | 0.036 | -0.031, 0.102 | 0.288 | |
| Mood disturbance | -0.001 | -0.002, 0.000 | 0.123 | -0.001 | -0.002, 0.001 | 0.272 | -0.001 | -0.003, 0.000 | 0.095 | |
| Sleep disturbance | 0.007 | 0.001, 0.014 | **0.026** | 0.004 | -0.002, 0.010 | 0.216 | 0.004 | -0.003, 0.011 | 0.209 | |
| Fatigue | -0.002 | -0.019, 0.015 | 0.797 | -0.002 | -0.019, 0.016 | 0.863 | -0.002 | -0.022, 0.017 | 0.83 | |
| Age | -0.003 | -0.006, -0.001 | **0.019** | -0.003 | -0.005, 0.000 | 0.058 | -0.001 | -0.004, 0.001 | 0.284 | |
| N | 78 | | | 73 | | | 66 | | | |
| R^2^ / R^2^ adjusted | 0.154 / 0.069 | | | 0.084 / -0.015 | | | 0.115 / 0.009 | | | |
|  | **Reaction time difference** | | | | | | | | | |
|  | **VV** | | | **VS** | | | **AU** | | | |
| *Predictors* | *Estim.* | *95% CI* | *p* | *Estim.* | *95% CI* | *p* | *Estim.* | *95% CI* | | *p* |
| (Intercept) | 94.701 | -142.595, 331.997 | 0.429 | 148.159 | -131.216, 427.534 | 0.293 | -55.327 | -307.614, 196.960 | | 0.662 |
| Pain duration (log) | -1.709 | -45.242, 41.824 | 0.938 | 2.343 | -51.641, 56.326 | 0.931 | 11.235 | -35.946, 58.416 | | 0.635 |
| Pain severity | -24.403 | -54.207, 5.402 | 0.107 | -20.115 | -60.936, 20.707 | 0.329 | -7.109 | -40.233, 26.015 | | 0.669 |
| Opioids use [yes] | 47.771 | -66.510, 162.051 | 0.407 | 35.459 | -113.608, 184.526 | 0.636 | -48.626 | -180.426, 83.175 | | 0.463 |
| Mood disturbance | 1.332 | -0.971, 3.634 | 0.253 | -2.153 | -5.183, 0.876 | 0.161 | 1.457 | -1.284, 4.199 | | 0.292 |
| Sleep disturbance | 8.911 | -2.848, 20.670 | 0.135 | 6.925 | -7.040, 20.890 | 0.326 | 1.467 | -12.154, 15.088 | | 0.83 |
| Fatigue | -13.477 | -43.503, 16.550 | 0.374 | 2.29 | -36.657, 41.236 | 0.907 | -9.346 | -48.145, 29.452 | | 0.631 |
| Age | 5.778 | 1.073, 10.484 | **0.017** | 3.009 | -2.801, 8.819 | 0.305 | 5.49 | 0.218, 10.761 | | **0.042** |
| N | 78 | | | 73 | | | 66 | | | |
| R^2^ / R^2^ adjusted | 0.165 / 0.081 | | | 0.090 / -0.008 | | | 0.113 / 0.006 | | | |

VV: visuo-verbal task; VS: visuo-spatial task; AU: auditory-temporal task. Estim., unstandardised regression coefficients with 95% confidence intervals.95% CI, 95% confidence interval; N, number of participants; R^2^, explained variance.
Statistically significant results are marked in bold font.

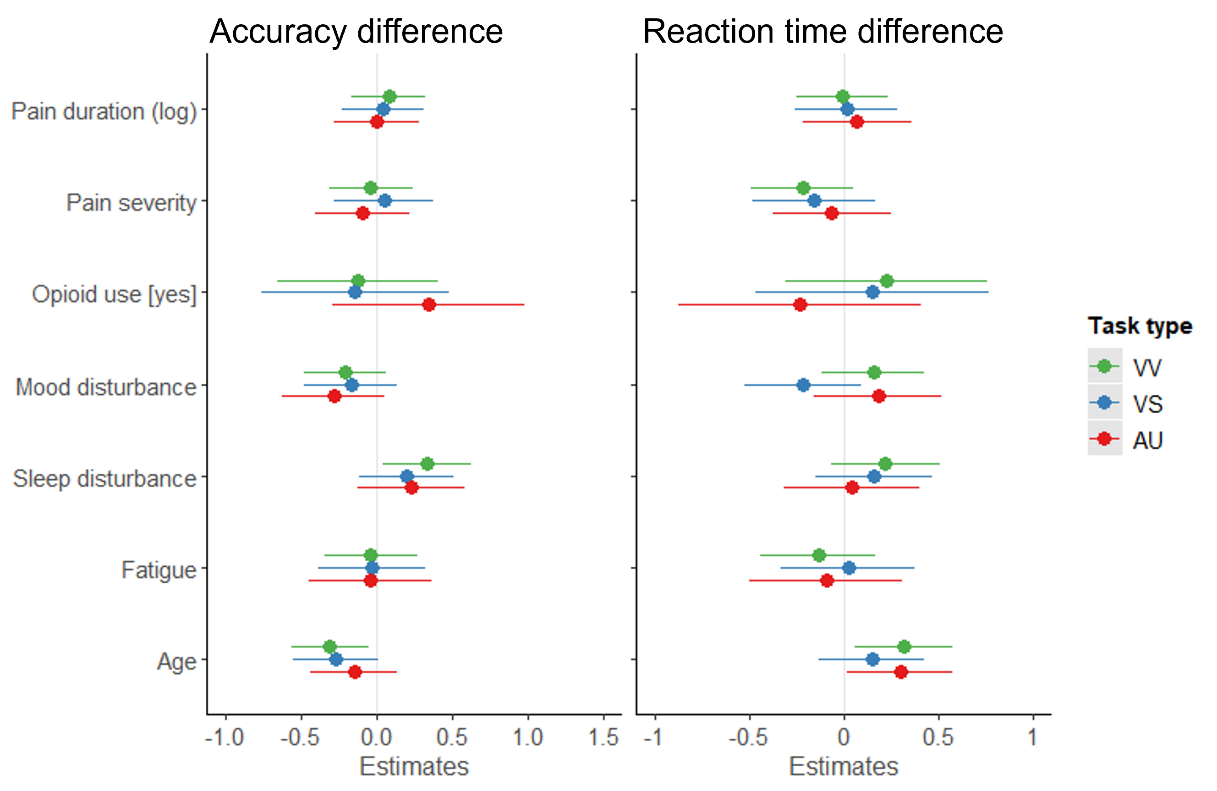

Figure S 17. Standardised coefficients with 95% confidence intervals

##### 1.9.2 Predictors of d’ difference scores

Pain duration did not independently predict change in sensitivity in any task (Table S 19). Only greater sleep disturbance predicted less decline in sensitivity with increasing cognitive load in visuo-verbal task.

Table S 19. Predictors of d’ difference scores (2-back – 0/1-back) for each task type.

|  | **VV** | | | **VS** | | | **AU** | | |
| --- | --- | --- | --- | --- | --- | --- | --- | --- | --- |
| *Predictors* | *Estim.* | *95% CI* | *p* | *Estim.* | *95% CI* | *p* | *Estim.* | *95% CI* | *p* |
| (Intercept) | -1.217 | -2.294, -0.141 | **0.027** | -0.7 | -1.893, 0.493 | 0.246 | -1.498 | -2.440, -0.556 | **0.002** |
| Pain duration (log) | -0.028 | -0.225, 0.170 | 0.78 | 0.018 | -0.213, 0.248 | 0.878 | -0.037 | -0.213, 0.139 | 0.674 |
| Pain severity | -0.011 | -0.146, 0.124 | 0.87 | 0.041 | -0.134, 0.215 | 0.643 | 0.029 | -0.095, 0.152 | 0.645 |
| Opioids use [yes] | -0.002 | -0.520, 0.517 | 0.995 | -0.119 | -0.755, 0.518 | 0.711 | 0.155 | -0.337, 0.647 | 0.531 |
| Mood disturbance | -0.008 | -0.018, 0.002 | 0.131 | -0.003 | -0.016, 0.010 | 0.665 | -0.006 | -0.016, 0.004 | 0.241 |
| Sleep disturbance | 0.072 | 0.018, 0.125 | **0.009** | -0.006 | -0.066, 0.054 | 0.843 | 0.037 | -0.014, 0.088 | 0.153 |
| Fatigue | -0.041 | -0.177, 0.095 | 0.547 | 0 | -0.167, 0.166 | 0.997 | -0.05 | -0.195, 0.095 | 0.492 |
| Age | -0.015 | -0.037, 0.006 | 0.153 | -0.015 | -0.040, 0.009 | 0.22 | 0 | -0.020, 0.019 | 0.967 |
| Observations | 78 | | | 73 | | | 66 | | |
| R^2^ / R^2^ adjusted | 0.120 / 0.033 | | | 0.037 / -0.067 | | | 0.086 / -0.024 | | |

VV: visuo-verbal task; VS: visuo-spatial task; AU: auditory-temporal task. Estim., unstandardised regression coefficients with 95% confidence intervals.95% CI, 95% confidence interval; R^2^, explained variance.
Statistically significant results are marked in bold font.

##### 1.9.3 Predictors of sustained attention performance

Lower fatigue was the only independent predictor of greater RT variability in the sustained attention task (Table S 20).

Table S 20. Predictors of reaction time variability in the 0-back visuo-verbal task.

|  | **0-back RT variability** | | |
| --- | --- | --- | --- |
| *Predictors* | *Estimates* | *95% CI* | *p* |
| (Intercept) | 0.199 | 0.141, 0.257 | **<0.001** |
| Pain duration (log) | -0.009 | -0.020, 0.002 | 0.124 |
| Pain severity | 0.005 | -0.002, 0.013 | 0.168 |
| Opioids use [yes] | 0.011 | -0.019, 0.040 | 0.47 |
| Mood disturbance | 0 | -0.000, 0.001 | 0.334 |
| Sleep disturbance | 0.002 | -0.001, 0.005 | 0.156 |
| Fatigue | -0.009 | -0.017, -0.002 | **0.02** |
| Age | 0 | -0.001, 0.002 | 0.506 |
| Observations | 90 | | |
| R^2^ / R^2^ adjusted | 0.142 / 0.069 | | |

Estim., unstandardised regression coefficients with 95% confidence intervals.95% CI, 95% confidence interval; R^2^, explained variance.
Statistically significant results are marked in bold font.

##### 1.9.4 Predictors of accuracy and reaction times for each task type and load condition

To explore whether pain duration, pain intensity, emotional distress/mood disturbance, fatigue, sleep disturbance, medication, and age predict the performance of participants with chronic pain on each individual task, we fitted multiple regression models with these predictors and each un-subtracted performance metric for each 0/1-back and 2-back task as outcomes (Table S 21). Similar to the main analysis, current pain and fatigue ratings were highly correlated with BPI pain severity (rs 0.76-0.81) and baseline fatigue (rs 0.49-0.75), respectively, thus the latter measures were included in the regression models, along with log-transformed pain duration, regular opioid use, mood disturbance, sleep disturbance, and age. Pain duration was an independent predictor of accuracy in 0-back visuo-verbal task and 1-back visuo-spatial task; however, the direction of this effect was opposite to what we hypothesised, i.e., longer pain duration was associated with better accuracy in these tasks. Additionally, greater sleep disturbance was related to better accuracy on visuo-spatial 1-back and 2-back tasks, whereas greater mood disturbance was associated with slower RTs on the auditory-temporal 2-back task. Finally, older age predicted lower accuracy on visuo-verbal and visuo-spatial 2-back tasks, and slower RTs on all tasks except for the auditory-temporal 2-back task.

Table S 21. Predictors of accuracy and reaction times for each task type and load condition (0/1-back, 2-back) separately.

| **Accuracy** |  |  |  |  |  |  |  |  |  |  |  |  |  |  |  |  |  |  |
| --- | --- | --- | --- | --- | --- | --- | --- | --- | --- | --- | --- | --- | --- | --- | --- | --- | --- | --- |
|  | **VV 0-back** |  |  | **VV 2-back** |  |  | **VS 1-back** |  |  | **VS 2-back** |  |  | **AU 1-back** |  |  | **AU 2-back** |  |  |
| *Predictors* | *Estim.* | *95% CI* | *p* | *Estim.* | *95% CI* | *p* | *Estim.* | *95% CI* | *p* | *Estim.* | *95% CI* | *p* | *Estim.* | *95% CI* | *p* | *Estim.* | *95% CI* | *p* |
| (Intercept) | 0.931 | 0.888, 0.975 | **<0.001** | 0.848 | 0.716, 0.979 | **<0.001** | 0.837 | 0.745, 0.930 | **<0.001** | 0.771 | 0.638, 0.904 | **<0.001** | 0.862 | 0.747, 0.977 | **<0.001** | 0.72 | 0.594, 0.846 | **<0.001** |
| Pain duration (log) | 0.009 | 0.001, 0.018 | **0.031** | 0.018 | -0.007, 0.042 | 0.151 | 0.02 | 0.002, 0.038 | **0.029** | 0.019 | -0.006, 0.044 | 0.13 | 0.013 | -0.009, 0.034 | 0.239 | 0.015 | -0.009, 0.039 | 0.205 |
| Pain severity | 0.002 | -0.004, 0.008 | 0.488 | 0 | -0.017, 0.016 | 0.96 | -0.009 | -0.022, 0.004 | 0.158 | -0.004 | -0.022, 0.014 | 0.632 | -0.006 | -0.021, 0.009 | 0.436 | -0.01 | -0.026, 0.007 | 0.258 |
| Opioids use [yes] | -0.013 | -0.035, 0.009 | 0.243 | -0.029 | -0.093, 0.034 | 0.359 | -0.027 | -0.076, 0.021 | 0.259 | -0.033 | -0.101, 0.034 | 0.328 | -0.001 | -0.060, 0.059 | 0.986 | 0.026 | -0.041, 0.093 | 0.437 |
| Mood disturbance | 0 | -0.000, 0.000 | 0.932 | -0.001 | -0.002, 0.000 | 0.119 | 0 | -0.001, 0.001 | 0.503 | -0.001 | -0.002, 0.000 | 0.145 | 0 | -0.001, 0.002 | 0.389 | -0.001 | -0.002, 0.001 | 0.328 |
| Sleep disturbance | -0.002 | -0.004, 0.001 | 0.145 | 0.006 | -0.001, 0.012 | 0.072 | 0.005 | 0.001, 0.010 | **0.022** | 0.008 | 0.002, 0.015 | **0.013** | -0.002 | -0.008, 0.004 | 0.532 | 0.001 | -0.006, 0.007 | 0.847 |
| Fatigue | 0.004 | -0.002, 0.010 | 0.167 | 0.002 | -0.014, 0.019 | 0.784 | 0.001 | -0.012, 0.014 | 0.881 | -0.001 | -0.019, 0.017 | 0.905 | -0.002 | -0.017, 0.013 | 0.789 | -0.001 | -0.020, 0.019 | 0.958 |
| Age | 0 | -0.001, 0.001 | 0.628 | -0.003 | -0.006, -0.001 | **0.013** | -0.001 | -0.003, 0.001 | 0.436 | -0.003 | -0.006, -0.000 | **0.038** | -0.001 | -0.003, 0.002 | 0.614 | -0.002 | -0.005, 0.000 | 0.111 |
| N | 90 |  |  | 78 |  |  | 82 |  |  | 75 |  |  | 74 |  |  | 67 |  |  |
| R^2^ / R^2^ adjusted | 0.113 / 0.038 |  |  | 0.169 / 0.086 |  |  | 0.219 / 0.145 |  |  | 0.185 / 0.100 |  |  | 0.047 / -0.054 |  |  | 0.126 / 0.022 |  |  |
| **Reaction times** |  |  |  |  |  |  |  |  |  |  |  |  |  |  |  |  |  |  |
|  | **VV 0-back** |  |  | **VV 2-back** |  |  | **VS 1-back** |  |  | **VS 2-back** |  |  | **AU 1-back** |  |  | **AU 2-back** |  |  |
| *Predictors* | *Estim.* | *95% CI* | *p* | *Estim.* | *95% CI* | *p* | *Estim.* | *95% CI* | *p* | *Estim.* | *95% CI* | *p* | *Estim.* | *95% CI* | *p* | *Estim.* | *95% CI* | *p* |
| (Intercept) | 397.291 | 250.401, 544.182 | **<0.001** | 592.527 | 336.261, 848.794 | **<0.001** | 671.611 | 477.739, 865.484 | **<0.001** | 789.703 | 477.881, 1101.525 | **<0.001** | 944.684 | 709.539, 1179.829 | **<0.001** | 904.556 | 576.921, 1232.190 | **<0.001** |
| Pain duration (log) | -4.501 | -33.011, 24.009 | 0.754 | -7.742 | -54.755, 39.271 | 0.744 | -9.713 | -47.182, 27.755 | 0.607 | -6.887 | -65.832, 52.058 | 0.816 | 10.637 | -33.377, 54.650 | 0.631 | 8.352 | -53.864, 70.568 | 0.789 |
| Pain severity | 1.899 | -17.452, 21.250 | 0.846 | -27.126 | -59.314, 5.061 | 0.097 | -4.393 | -31.380, 22.594 | 0.747 | -26.532 | -68.953, 15.889 | 0.216 | 5.605 | -24.630, 35.839 | 0.712 | -12.445 | -56.161, 31.270 | 0.571 |
| Opioids use [yes] | -72.581 | -146.891, 1.729 | 0.055 | 0.726 | -122.691, 124.142 | 0.991 | 18.924 | -81.634, 119.483 | 0.709 | 63.271 | -95.834, 222.375 | 0.43 | -2.217 | -124.326, 119.892 | 0.971 | -37.418 | -210.280, 135.444 | 0.666 |
| Mood disturbance | 1.434 | -0.118, 2.985 | 0.07 | 1.942 | -0.545, 4.428 | 0.124 | 0.994 | -1.126, 3.115 | 0.353 | -0.809 | -4.162, 2.543 | 0.631 | 2.136 | -0.200, 4.471 | 0.072 | 4.358 | 0.743, 7.973 | **0.019** |
| Sleep disturbance | 0.707 | -6.705, 8.120 | 0.85 | 9.728 | -2.971, 22.427 | 0.131 | -2.766 | -12.255, 6.722 | 0.563 | 5.094 | -10.285, 20.472 | 0.511 | -1.99 | -14.227, 10.248 | 0.746 | 7.68 | -9.817, 25.178 | 0.383 |
| Fatigue | -8.813 | -28.630, 11.004 | 0.379 | -18.682 | -51.109, 13.745 | 0.254 | -5.156 | -32.424, 22.111 | 0.707 | 2.976 | -39.075, 45.026 | 0.888 | -26.732 | -57.503, 4.038 | 0.087 | -47.854 | -98.569, 2.861 | 0.064 |
| Age | 5.389 | 2.382, 8.395 | **0.001** | 9.526 | 4.445, 14.608 | **<0.001** | 6.352 | 2.357, 10.346 | **0.002** | 8.233 | 1.877, 14.588 | **0.012** | 3.115 | -1.779, 8.008 | 0.208 | 8.408 | 1.653, 15.163 | **0.016** |
| N | 90 | | | 78 | | | 82 | | | 75 | | | 74 | | | 67 | | |
| R^2^ / R^2^ adjusted | 0.207 / 0.140 | | | 0.242 / 0.166 | | | 0.142 / 0.061 | | | 0.148 / 0.059 | | | 0.097 / 0.001 | | | 0.192 / 0.096 | | |

VV: visuo-verbal task; VS: visuo-spatial task; AU: auditory-temporal task. Estim., unstandardised regression coefficients with 95% confidence intervals.95% CI, 95% confidence interval; R^2^, explained variance.
Statistically significant results are marked in bold font.

#### 1.11 Hierarchical Drift Diffusion Models (HDDMs)

##### 1.11.1 Session 1: Model selection

The full models, where each considered parameter was allowed to vary both by group and cognitive load, were found to be the winning models according to the lowest DICs (Table S 22), that is, these models best explained the data at hand (Spiegelhalter et al., 2002). Posterior predictive checks indicated that these models could reproduce the key patterns in the data, as the summary statistics of the new datasets (n = 500) simulated from the posteriors of the fitted models were all within the 95% credible intervals of the summary statistics of the observed dataset (Table S 23). Altogether, while the winning full models suggest that both group and cognitive load affect each of the considered parameters, this is not always reflected in the extent of overlap of their posterior distributions.

While the winning full models suggest that both group and cognitive load affect each of the considered parameters, this is not always reflected in the extent of overlap of their posterior distributions. It is noteworthy that similar but simplified models, including only the effect of cognitive load on certain parameters, achieved DICs relatively close to the lowest DIC values (see Table S 22).

For instance, in the VV task, there was considerable overlap between the posteriors of non-decision time separated by group, suggesting that group differences had limited impact on this parameter. Models without the effect of group on drift rate and/or non-decision time (M11 and M13) showed DICs in a similar range to that of the full model. To verify whether the effects of group on drift rate and threshold separation in this task were robust, we ran a post-hoc model omitting the group effect on t, which achieved DIC -7166.20 (compared to -7181.10 of the full model).

In the VS task, the posteriors of non-decision time and threshold separation also showed considerable overlap when separated by group, suggesting limited effects of group on these parameters. DICs of the models omitting the effects of group on drift rate, threshold separation, and non-decision time (M11 and M13) were very similar to that of the full model. Therefore, to verify whether the effect of group on drift rate in this task was robust, we ran an additional post-hoc model including both the effects of group and cognitive load on drift rate, but only the effects of cognitive load on non-decision time and threshold separation, and indeed this model performed nearly as well as the full model (DIC = 18220.67 compared to 18220.38).

In the AU task, there was considerable overlap between the posteriors of drift rate and non-decision time separated by group, suggesting that group differences had limited effect on these parameters. Indeed, the models that omitted the group effect on drift rate (M11), or both drift rate and non-decision time (M13), were the second- and third-best models according to their DICs, respectively.

Therefore, the models that did not account for the effects of group on v (M11) and t (M13) in the VV and AU tasks, and for the effects of group on v (M11), a (M12), and t (M13) in the VS task, appeared to explain the observed data comparably well to the full models (M01). This suggests that group differences likely make rather minor contribution to explaining the observed patterns of drift rate, non-decision time, and threshold separation, which appear to be primarily driven by the effects of cognitive load.

Table S 22. Tested model structures with their corresponding DICs for each task type.

|  | **Model** | **Model structure (varied parameters)** | | | **Model selection** | | |
| --- | --- | --- | --- | --- | --- | --- | --- |
|  |  | ***v*** | ***t*** | ***a*** | **VV DIC** | **VS DIC** | **AU DIC** |
| Null | M00 |  |  |  | 18656.64 | 35076.85 | 42779.37 |
| Full | M01 | group, load | group, load | group, load | **-7181.10** | **18220.38** | **34909.50** |
| Step 1 | M02 | group, load |  | group | -4101.36 | 20357.04 | 36366.69 |
|  | M03 | group |  | group | 7730.73 | 23986.47 | 38844.86 |
|  | M04 | load |  | group | -4102.98 | 20370.27 | 36364.51 |
|  | M05 | group, load |  | group, load | -5496.70 | 19435.49 | 36024.69 |
|  | M06 | group |  | group, load | -225.82 | 21406.74 | 37934.06 |
|  | M07 | load |  | group, load | *-5516.32* | 19425.69 | *36020.81* |
|  | M08 | group, load |  | load | -5507.99 | 19428.53 | 36029.19 |
|  | M09 | group |  | load | -218.39 | 21429.15 | 37936.49 |
|  | M10 | load |  | load | -5505.08 | *19412.27* | 36026.41 |
| Step 2 | M11 (VV & AU) | load | group, load | group, load | *-7154.75* |  | *34911.84* |
|  | M12 (VV & AU) | load | group | group, load | -5495.35 |  | 36027.79 |
|  | M13 (VV & AU) | load | load | group, load | -7140.63 |  | 34926.24 |
|  | M11 (VS) | load | group, load | load |  | 18224.56 |  |
|  | M12 (VS) | load | group | load |  | 19419.47 |  |
|  | M13 (VS) | load | load | load |  | *18222.36* |  |

*a*, threshold separation; AU, auditory-temporal task; DIC, deviance information criterion; VV, visuo-verbal task; *t*, non-decision time; *v*, drift rate; VS, visuo-spatial task.
DICs in italics indicate the winning models at each step, whereas DICs in bold indicate the final winning models (lowest DICs).

The finding that cognitive load affected a non-decisional parameter *t* is somewhat unexpected, as the amount of required cognitive resources in the task should not change the time required for stimulus encoding and response execution. However, this effect of faster non-decision time under lower cognitive load could potentially be explained by sensory priming in 1-back conditions, whereby the processing of the target stimulus could have been facilitated by the presentation of the same stimulus on the preceding trial. Similarly, in the 0-back condition, participants could have been primed to recognise one specific stimulus in the stream of letters. Alternatively, slower non-decision time under higher cognitive load could potentially be explained by the participants being motivated to respond faster while having to maintain two previous stimuli in their working memory, so that they can use the delay between the stimuli to rehearse the memory content before it needs to be updated again in the upcoming trial. Unfortunately the drift diffusion framework does not allow to dissociate the sensory encoding and response execution components of non-decision time. Another potential explanation is that non-decision time could include the time needed to retrieve the representation of n-back stimuli (memory access; (Ratcliff & McKoon, 2008; Shepherdson et al., 2018) – this process would take longer when more items need to be maintained in working memory.

The lack of group effects on drift rate are also somewhat surprising considering previously found overall slowing of reaction times in individuals with chronic pain. Longer non-decision time was also previously observed in older, compared to younger, age groups in working memory tasks (Archambeau et al., 2020; Thurm et al., 2018); see also (Ratcliff et al., 2004, 2010) for recognition memory. So, it is possible that we did not observe a similar effect in our study because both groups were age-matched.

Table S 23. Posterior predictive checks: summary statistics over the simulated datasets compared to the observed data.

|  | **VV** | | | | | | | | **VS** | | | | | | | | **AU** | | | | | | | |
| --- | --- | --- | --- | --- | --- | --- | --- | --- | --- | --- | --- | --- | --- | --- | --- | --- | --- | --- | --- | --- | --- | --- | --- | --- |
| Statistic | Observed mean | Mean | SD | SEM | MSE | 95% CI | Quantile | Mahalanobis | Observed mean | Mean | SD | SEM | MSE | 95% CI | Quantile | Mahalanobis | Observed mean | Mean | SD | SEM | MSE | 95% CI | Quantile | Mahalanobis |
| Accuracy | 0.31 | 0.50 | 0.44 | 0.04 | 0.23 | Yes | 46.89 | 0.43 | 0.33 | 0.52 | 0.41 | 0.04 | 0.21 | Yes | 43.87 | 0.47 | 0.36 | 0.50 | 0.33 | 0.02 | 0.13 | Yes | 40.96 | 0.42 |
| Mean RT UB | 0.73 | 0.80 | 0.29 | 0.00 | 0.09 | Yes | 52.87 | 0.23 | 0.83 | 0.85 | 0.26 | 0.00 | 0.07 | Yes | 54.30 | 0.08 | 1.14 | 1.16 | 0.29 | 0.00 | 0.08 | Yes | 51.47 | 0.09 |
| SD RT UB | 0.32 | 0.24 | 0.19 | 0.01 | 0.04 | Yes | 73.42 | 0.43 | 0.37 | 0.29 | 0.19 | 0.01 | 0.04 | Yes | 72.50 | 0.42 | 0.42 | 0.38 | 0.21 | 0.00 | 0.04 | Yes | 62.66 | 0.17 |
| 10Q UB | 0.44 | 0.57 | 0.19 | 0.02 | 0.05 | Yes | 23.71 | 0.66 | 0.46 | 0.59 | 0.18 | 0.02 | 0.05 | Yes | 22.57 | 0.72 | 0.68 | 0.80 | 0.20 | 0.01 | 0.06 | Yes | 28.00 | 0.59 |
| 30Q UB | 0.55 | 0.65 | 0.22 | 0.01 | 0.06 | Yes | 38.61 | 0.44 | 0.62 | 0.67 | 0.20 | 0.00 | 0.04 | Yes | 45.08 | 0.27 | 0.89 | 0.92 | 0.23 | 0.00 | 0.05 | Yes | 50.46 | 0.14 |
| 50Q UB | 0.64 | 0.74 | 0.26 | 0.01 | 0.08 | Yes | 45.03 | 0.37 | 0.75 | 0.77 | 0.24 | 0.00 | 0.06 | Yes | 53.63 | 0.10 | 1.06 | 1.05 | 0.27 | 0.00 | 0.07 | Yes | 56.83 | 0.03 |
| 70Q UB | 0.77 | 0.86 | 0.34 | 0.01 | 0.12 | Yes | 51.97 | 0.26 | 0.92 | 0.92 | 0.31 | 0.00 | 0.09 | Yes | 58.52 | 0.01 | 1.28 | 1.25 | 0.34 | 0.00 | 0.12 | Yes | 59.10 | 0.08 |
| 90Q UB | 1.14 | 1.09 | 0.50 | 0.00 | 0.25 | Yes | 64.08 | 0.11 | 1.32 | 1.20 | 0.47 | 0.01 | 0.23 | Yes | 67.17 | 0.26 | 1.66 | 1.63 | 0.51 | 0.00 | 0.26 | Yes | 58.52 | 0.06 |
| Mean RT LB | -0.74 | -0.79 | 0.30 | 0.00 | 0.09 | Yes | 44.11 | 0.16 | -0.93 | -0.98 | 0.31 | 0.00 | 0.10 | Yes | 48.41 | 0.17 | -1.12 | -1.16 | 0.31 | 0.00 | 0.10 | Yes | 49.00 | 0.13 |
| SD RT LB | 0.33 | 0.23 | 0.19 | 0.01 | 0.05 | Yes | 75.78 | 0.56 | 0.35 | 0.29 | 0.19 | 0.00 | 0.04 | Yes | 68.10 | 0.29 | 0.40 | 0.37 | 0.22 | 0.00 | 0.05 | Yes | 60.32 | 0.15 |
| 10Q LB | 0.45 | 0.57 | 0.22 | 0.02 | 0.06 | Yes | 26.46 | 0.57 | 0.58 | 0.70 | 0.25 | 0.01 | 0.08 | Yes | 31.57 | 0.48 | 0.69 | 0.82 | 0.24 | 0.02 | 0.08 | Yes | 30.74 | 0.54 |
| 30Q LB | 0.55 | 0.65 | 0.24 | 0.01 | 0.07 | Yes | 41.42 | 0.41 | 0.72 | 0.80 | 0.27 | 0.01 | 0.08 | Yes | 44.27 | 0.30 | 0.87 | 0.93 | 0.26 | 0.00 | 0.07 | Yes | 49.25 | 0.21 |
| 50Q LB | 0.64 | 0.74 | 0.28 | 0.01 | 0.08 | Yes | 48.74 | 0.33 | 0.85 | 0.91 | 0.30 | 0.00 | 0.09 | Yes | 49.91 | 0.21 | 1.05 | 1.06 | 0.29 | 0.00 | 0.09 | Yes | 56.27 | 0.03 |
| 70Q LB | 0.79 | 0.85 | 0.34 | 0.00 | 0.12 | Yes | 55.75 | 0.18 | 1.03 | 1.06 | 0.35 | 0.00 | 0.13 | Yes | 54.85 | 0.11 | 1.26 | 1.25 | 0.36 | 0.00 | 0.13 | Yes | 58.25 | 0.04 |
| 90Q LB | 1.17 | 1.07 | 0.49 | 0.01 | 0.25 | Yes | 67.22 | 0.21 | 1.39 | 1.34 | 0.49 | 0.00 | 0.24 | Yes | 62.38 | 0.09 | 1.65 | 1.61 | 0.52 | 0.00 | 0.27 | Yes | 58.28 | 0.06 |

95% CI, summary statistics within 95% credible interval; LB, lower boundary; UB, upper boundary; Q, quantile; RT, reaction time; SD, standard deviation; Mean, mean of the simulated data; SEM, standard error of the mean; MSE, mean-squared error; Mahalanobis – Mahalanobis distance;

##### 1.11.2 Session 2: Comparison of full model effects to session 1

To assess whether similar parameter distributions replicated at the retest stage, we fitted the winning full HDDMs to session 2 data (Figure S 18). Cognitive load had a consistent effect on all parameters across tasks, as observed in session 1 (except for t in AU tasks for controls, but trending in the same direction; P = 0.93).

The effects on drift rate were largely consistent with those in session 1, that is, controls had higher drift rate in the VV 0-back task (P = 0.99) and marginally in AU 2-back task (P = 0.95) compared to participants with chronic pain, however, the group difference in VS 1-back task observed in session 1 was not apparent in session 2 (P = 0.32).

Similar to session 1, group did not considerably affect non-decision time in any of the tasks in session 2 (Ps 0.17 – 0.94).

In contrast to session 1, group did not affect threshold separation in visuo-verbal 0-back task (P = 0.09); instead, there was a tendency for chronic pain participants to have greater threshold separation than controls in visuo-spatial 1-back task in session 2 (P = 0.95), which was not the case in session 1. The effect of group on threshold separation in auditory-temporal task was in line with session 1, i.e., participants with chronic pain had greater threshold separation than controls in both 1-back and 2-back task conditions (Ps = 1).

Overall, while consistent pattern and direction of results was observed across sessions 1 and 2, some differences appear to be less pronounced in session 2. One contributing factor could be that the then umber of observations available for session 2 analyses was smaller than for session 1.

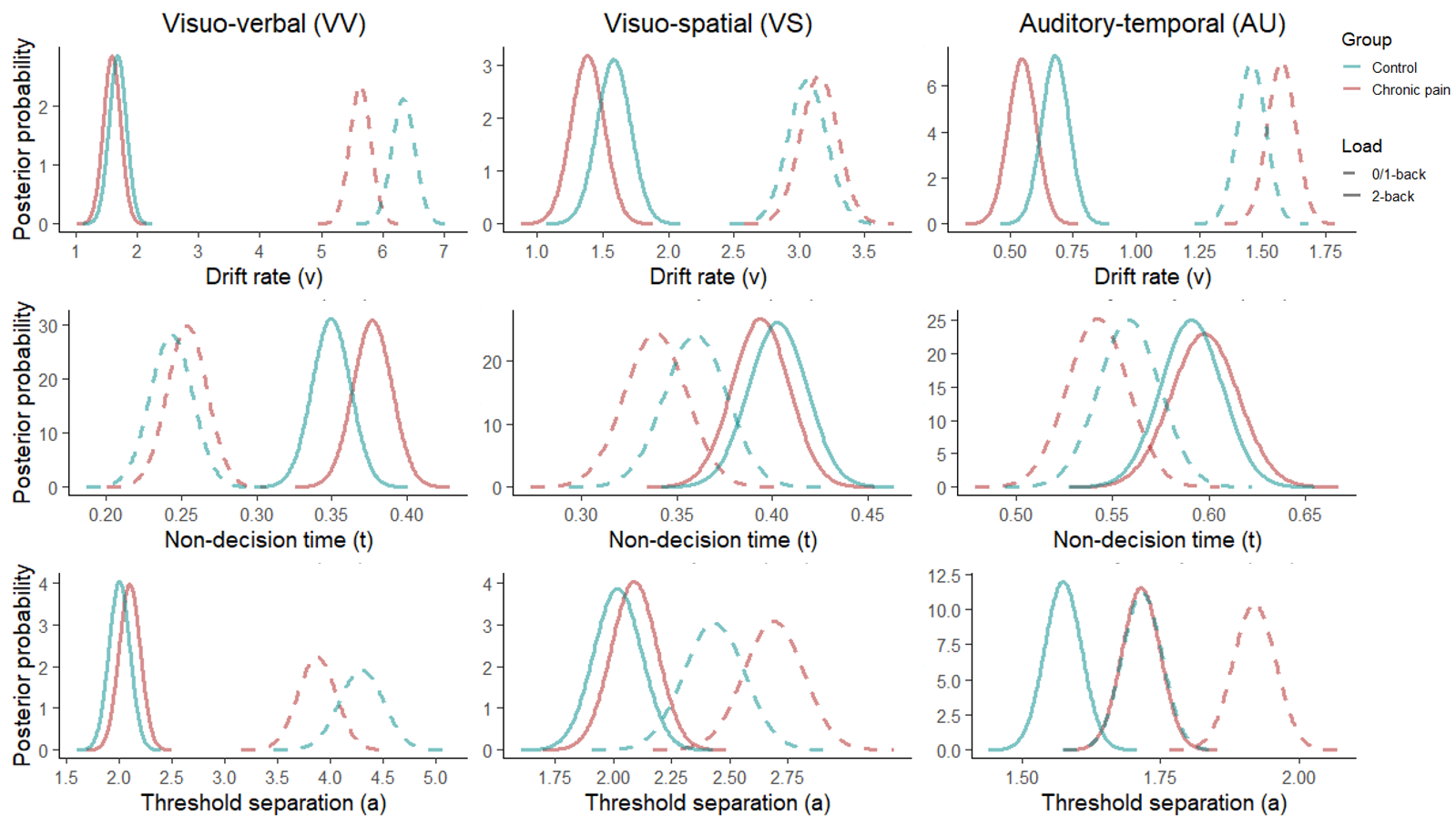
Figure S 18. Posterior probability distributions of group (control, chronic pain) and condition (0/1-back, 2-back) means of the HDDM parameters drift rate v (top row), non-decision time t (middle row), and threshold separation a (bottom row) in the visuo-verbal (VV; left column), visuo-spatial (VS; middle column), and auditory-temporal (AU; right column) tasks from Session 2. Note that the axis limits and intervals are specific to each task and parameter to best visualise the (non-)overlap between posterior distributions.
